# Development and validation of polygenic risk profiles of schizophrenia

**DOI:** 10.1101/2025.04.29.25326656

**Authors:** Ersoy Kocak, Joonas Naamanka, Tobias Gradinger, Fiona Klaassen, Johannes Nitsche, Philipp Grotehusmann, Kristina Adorjan, Linda A. Antonucci, Giuseppe Blasi, Monika Budde, Piergiuseppe Di Palo, Maria Heilbronner, Gianluca C. Kikidis, Alba Navarro-Flores, Mojtaba Oraki Kohshour, Sergi Papiol, Alessandra Raio, Antonio Rampino, Daniela Reich-Erkelenz, Eva C. Schulte, Fanny Senner, Leonardo Sportelli, Alessandro Bertolino, Peter Falkai, Urs Heilbronner, Giulio Pergola, Thomas G. Schulze, Andreas Meyer-Lindenberg, Fabian Streit, Emanuel Schwarz, FinnGen

## Abstract

Understanding the genetic architecture of schizophrenia (SCZ) is invaluable for the development of personalized treatment. In three independent cohorts, using an automated pipeline, we calculated 413 psychiatry-relevant polygenic scores. Using these scores, machine learning was applied to stratify SCZ patients into two genetic risk profiles (GRP) in FinnGen (n_SCZ_=7 486), and to validate these results in the PsyCourse Study (n_SCZ_=419) and Bari (n_SCZ_=531). While the two GRPs showed comparable polygenic SCZ risk, they were primarily distinguished by a greater predisposition for neuroticism, depression-related traits, and lower cognitive performance in GRP 1. The genetic prediction of GRP 1 was phenotypically characterized by an increased prevalence of SCZ and a more severe and complex clinical manifestation. This illustrates that the penetrance of genetic SCZ risk might partially depend on the predisposition for the aforementioned traits through pleiotropic variants. Our results provide novel, replicable insights into the genetic architecture of SCZ and might inform future personalized treatments.

## Introduction

Schizophrenia (SCZ) is a severe, usually chronic mental disorder with a lifetime prevalence estimate of up to 1% and an incidence peak in early adulthood[1]. Its severity is underscored by a life expectancy approximately 15-years shorter than the general population[2] and a mere 10-20% competitive employment rate[1, 3, 4].

SCZ is a phenotypically complex disease, and its individually variable manifestations span multiple domains like perception, affect, cognition, behavior, speech, motor skills, and autonomic regulation being affected to varying degrees among patients. Furthermore, the complexity and heterogeneity of SCZ extend to treatment response with approximately one-third of patients responding poorly[5, 6]. Flattening of affect, avolition and cognitive decline (collectively negative symptoms), are especially persistent[7, 8]. Formulations of sub-classification of SCZ have been abandoned in the latest editions of the commonly used diagnostic manuals (ICD-11 and DSM-V) due to a lack of reliability and clinical utility[9].

Evidence suggests that SCZ encompasses a multitude of overlapping neurobiological processes[10, 11]. Identifying more homogenous subgroups in terms of these processes would advance the etiological understanding of the disorder and personalized medicine approaches[12]. In family and twin studies, heritability for SCZ has been estimated at 60%-80% overall[13–15], among the highest for psychiatric disorders, and the phenotypic and biological heterogeneity of SCZ has been hypothesized to depend in part on genetic heterogeneity[16–18]. Therefore, the investigation of genetic variation is a promising basis for understanding the source of that heterogeneity through stratification[1, 19, 20]. Genome-wide association studies (GWAS) of SCZ indicate that it is a highly polygenic disorder with thousands of common variants (i.e., single-nucleotide polymorphisms (SNPs)) contributing to SCZ liability, each exerting a small effect on overall disorder risk[21, 22]. It is estimated that around 25% of liability to SCZ is attributable to the total contribution of common genetic variation (i.e., SNP-heritability). To estimate the genetic predisposition of an individual to a disease or a trait, the effects conferred by individual SNPs can be summed up into polygenic scores (PGS) (also called polygenic risk scores (PRS)) for diseases and a wide variety of traits. These indices of polygenic predisposition show robust predictive performance in independent target samples, but they currently fall short of explaining disease liability to the degree suggested by SNP-heritability estimates[19, 23].

Therefore, considered together with moderate-to-high genetic correlations with other psychiatric disorders (e.g., *r_g_*=0.7 with bipolar disorder (BIP) and *r_g_* =0.35 with depression)[15, 22] and related traits (e.g., *r_g_*=0.21 with Neuroticism)[23, 24], an approach based on a broad spectrum of PGS seems promising. Furthermore, SCZ-PGS are primarily designed to distinguish cases from controls, a limitation potentially remedied by a multi-PGS strategy in case-case discrimination. While the PGS of SCZ combined with additional psychiatric PGS with known associations with SCZ have been used for subtyping[25], and a data-driven multi-PGS approach has demonstrated superior performance in predicting SCZ case-control status over single-PGS models[26], the latter strategy has not been applied to characterize patients based on distinct genetic risk profiles (GRPs) before.

Building on this recent research, we aimed to: (i) identify distinct GRPs in SCZ by integrating large numbers of PGS of psychiatry-relevant phenotypes using robust, reproducible machine learning (ML) methods; (ii) validate these distinct GRPs in independent datasets; (iii) characterize the SCZ patients showing these GRPs in terms of clinical manifestations.

To achieve this, we developed *PRS*cope, a framework that generates PGS from a broad set of GWAS and then uses the resulting multi-PGS matrix to identify patient clusters through a data-driven approach and characterise them as GRPs based on their PGS compositions. Our analysis included data from a total of 8 436 patients with SCZ, and 28 325 healthy controls (HC) from three independent datasets, to identify and validate GRPs. In addition, we used data from 7 555 patients with BIP, and from 27 187 patients with Major Depressive Disorder (MDD), and from 37 539 HCs for a transdiagnostic analysis. Our method offers a novel approach to deconstructing the genetic heterogeneity of complex human disorders and may contribute to the personalization of medical care in SCZ and other disorders.

## Materials and methods

### Overview

The methods used can be separated into the following steps: I) Automated GWAS search, II) Quality control (QC) of the GWAS, III) QC of the genotype data in the target datasets, IV) Calculation of the corresponding PGS in the target datasets V) Clustering of patients based on multi-PGS similarities in the training dataset, VI) Predicting the cluster assignments in the validation datasets, VII) Phenotypic profiling, VIII) Genetic profiling. Steps I) - IV) were done using PRScope, and steps V) - VIII) were done by the ML framework implemented for phenotype or disease stratification (for a schematic overview of the Methods, see Figure 1).

**Figure 1.**
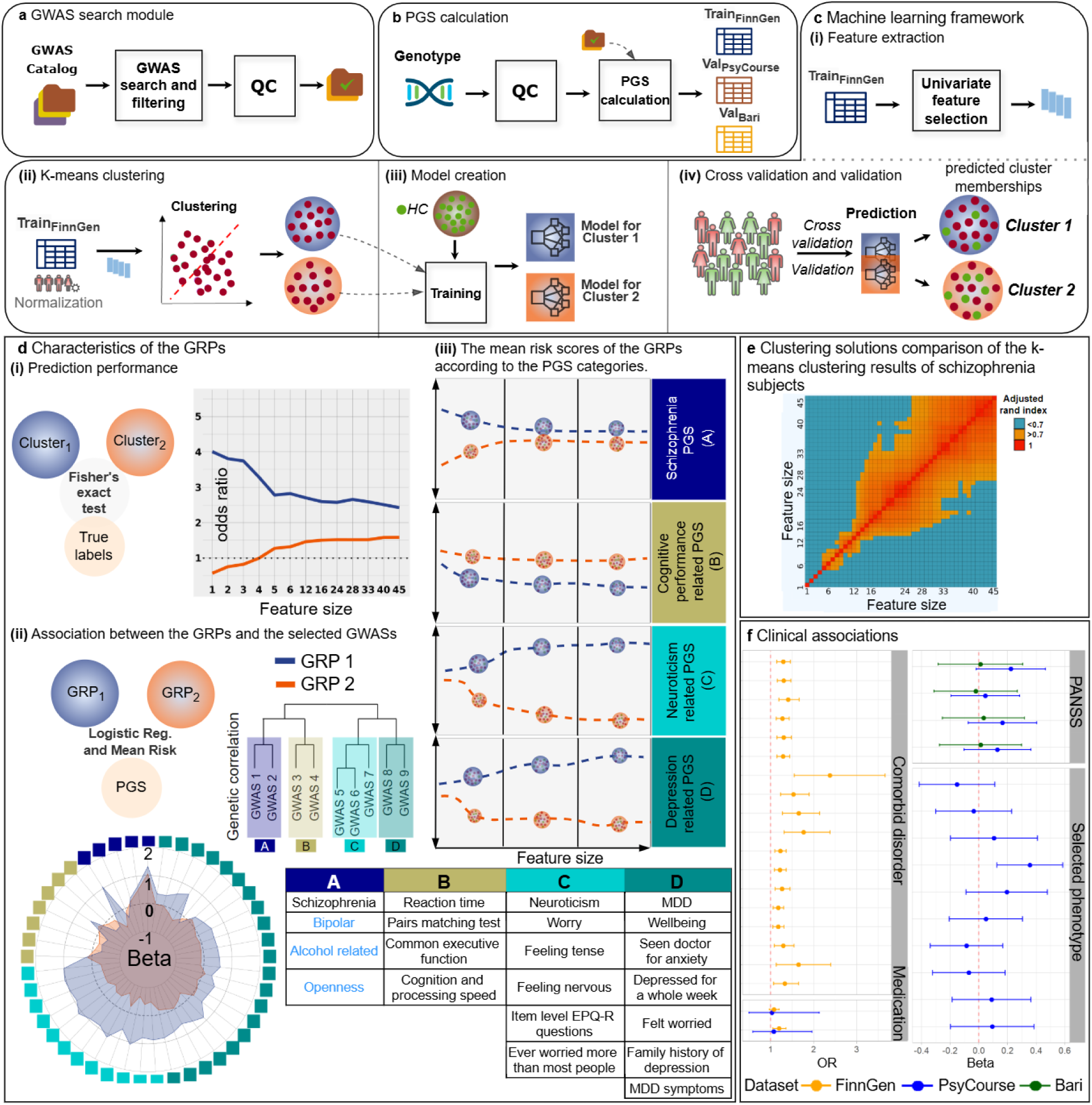
Overview of the PRScope framework and analysis pipeline. **a)** Psychiatry-related GWAS, annotated with relevant ontologies and fulfilling our filtering criteria, were downloaded from the GWAS Catalog and pre-processed for the subsequent PGS calculation. **b)** Genotype data from the three cohorts were pre-processed. PGS calculations for 413 pre-processed GWAS were performed for each GWAS using Bayesian-based PGS method LDPred2. **c)** ML was applied to identify two data-driven clusters with differing PGS profiles in large-scale training dataset. These GRPs were then validated in two independent datasets and through cross-validation in the discovery dataset. Definition of *Cluster*: Predicted cluster memberships derived from cluster-specific models include both HC and SCZ subjects. Definition of *GRP*: SCZ patients uniquely correctly predicted by these models. (i) Ranking of PGS with regard to their absolute correlation with SCZ. The top 45 PGS that showed the strongest correlation with SCZ were included in ML analysis (from 1 to 45, stepwise). To prevent data leakage during cross-validation in the training dataset, feature selection was performed separately for each cross-validation iteration. For validation, features were extracted using the entire training dataset. (ii) Clustering of SCZ patients based on a multi-PGS profile into two clusters via k-means clustering method using the selected features. The PGS of SCZ patients for a given GWAS were adjusted by subtracting the median of the HC and dividing by the standard deviation of the HC. (iii) Random forest ML was applied to train models for each cluster separately. (iv) Cluster memberships were predicted by the model in independent data and cross-validation test folds. **d)** The characterization of the multi-PGS signature of each GRP. (i) Associations between the predicted cluster memberships and diagnoses were quantified as odds ratios and significance was determined using two-sided Fisher’s exact test. (ii) The beta coefficients obtained from logistic regression represent the associations between each of the 45 most SCZ-relevant PGS and GRP relative to HC. The table shows the PGS categories identified based on genetic correlation among GWAS. SCZ category included only SCZ-PGS. Bipolar disorder, personality trait and alcohol related traits were not included in GRP characterization. (iii) PCs were computed separately for each GRP using all PGS within a PGS category, and the mean values of the first PCs were plotted for each GRP and feature size. **e)** The k-means clustering solution for a given feature size was compared with the clustering results of other feature sizes and visualized as a heatmap. **f)** To support the clinical validity of the identified GRPs, differences in clinical features were assessed across the investigated datasets.

### Cohorts

Data from FinnGen[27] release 12 (Train_FinnGen_) (7 486/27 288 cases/controls for SCZ, 7 555/11 198 for BIP, 27 187/26 993 for MDD. Controls for BIP and MDD are partly shared, but not with SCZ controls) were used for the identification of multi-PGS profiles, and to test associations. Data from the PsyCourse Study[28] (Val_PsyCourse_) (419/299 cases/controls for SCZ) and Bari cohort[21] (Val_Bari_) (531/738 cases/controls for SCZ) were used for GRP validation. All cohorts were independent of the original GWAS the PGS were derived from. Supplementary Tables 1-3 show the composition of the datasets after QC with respect to gender and diagnosis. Outliers resulting from systematic ancestry differences across all three datasets were removed using OGK estimator[29]. The first 20 genotype principal components (PCs) were used to detect the outliers and control for population stratification. Based on this method, the log distances above a threshold ( ≥ 4.5) were classified as outliers. (see Supplementary Methods 1.1, 1.2)

### PRScope

PRScope extracts GWAS from the GWAS Catalog[30] based on a predefined selection of psychiatry-relevant categories specified by a list of Experimental Factor Ontology[31] (EFO) ids, which were provided as input to the pipeline (see Supplementary Table 4). The harmonized GWAS fulfilling the filtering criteria and annotated with the provided EFOs were downloaded from GWAS Catalog and quality controlled, resulting in 348 GWAS (see Supplementary Table 5). Furthermore, 65 GWAS, not listed in the GWAS Catalog but accessible from Psychiatric Genomics Consortium (PGC), Complex Trait Genetics (CTG), and other public sources, were identified and included in the analysis, using the same harmonization pipeline[32] as applied by the GWAS Catalog. The corresponding PGS were then calculated for each of the subjects in the target genotype datasets using the Bayesian-based PGS method LDPred-2[33]. The application of PRScope resulted in quality-controlled PGS for 413 psychiatry-relevant phenotypes (see Supplementary Tables 6-10) that were computed in three cohorts (Train_FinnGen_, Val_PsyCourse_, Val_Bari_) (Figure 1a,1b) (see Supplementary Methods 1.3).

### Automated GWAS search module

A primary challenge for researchers conducting multi-PGS studies is selecting source GWAS from among 100 000+ available GWAS in the GWAS Catalog. It is crucial to include GWAS of traits that are of likely relevance to the target phenotype. To address this, we implemented a module to automate this selection process. This module uses the EFO ids to filter and collect relevant GWAS. First, the user performs a preliminary search in the EFO browser to select relevant ontologies for the target phenotype. Next, the user defines search criteria in the module configuration file, such as GWAS population, number of SNPs, sample size, and publication date (see Supplementary Methods 1.3.1.1). The module uses the R library *gwasrapidd* [34] to search the GWAS Catalog for studies annotated with the selected EFO ids, applying the user-defined filters. Finally, the output of the module is a list of GWAS Catalog summary statistic ids that meet the specified criteria (see Supplementary Figure 1).

### PGS calculation module

The PGS calculation module offers the advantage of quickly generating multiple PGS calculations using the LDPred2-auto script developed by Frei et al.[35]. This module requires minimal configuration, as it uses default parameters for calculation (see the original repository for details). The resulting multi-PGS matrices were adjusted for population stratification using the first 20 genotype PC. Resulting matrices were then transformed using the *scale* function of the R base package.

### Machine learning framework

ML was applied to identify and validate subgroups of patients with SCZ characterized by distinct multi-PGS profiles. The analytical pipeline involved: (a) ranking of PGS with regard to their absolute correlation with SCZ; (b) clustering of SCZ patients based on multi-PGS profile into two clusters via k-means clustering using the most relevant PGS; (c) training a separate random forest[36] model for each cluster to distinguish SCZ cases from HC; and (d) predicting cluster membership across cross- validation folds on Train_FinnGen_ and on validation datasets (models trained on Train_FinnGen_, and validated on Val_PsyCourse_ and Val_Bari_), resulting in one prediction from Cluster 1 model and one from Cluster 2 model for each dataset (Figure 1c).

These predictions were then used to evaluate the predictive accuracy of each model. SCZ patients predicted as patients by exactly one of the models were assigned to the corresponding group (hereafter referred to as GRP) and the GRPs were characterized based on their polygenic risk contributions, as well as psychopathological measures (Val_Bari_), comorbid disorders and medication information (Train_FinnGen_), and selected phenotypes (Val_PsyCourse_). The whole procedure was applied for each selected feature size separately, starting with the highest ranked PGS, adding PGS based on their ranking, resulting in a gradually increasing feature size (Figure 1d). For comparison, an alternative scheme where we group patients according to their predicted probabilities of having schizophrenia, as derived from prediction models trained on the same PGS as in our primary approach, was applied, hereafter referred to as *composite PGS*.

Genetic correlations among GWAS were used to categorize the 45 most SCZ-relevant PGS into meaningful PGS categories. Finally, this analysis was complemented by conjunctional False Discovery Rate analysis[37, 38] to investigate whether genetic susceptibility for such phenotypes was mediated by shared genetics between PGS categories. (see Supplementary Methods 1.4).

Given the limited validation sample sizes, we opted for a parsimonious two-cluster solution and assessed this decision in a post hoc analysis (see Supplementary Methods 1.4.1).

### Composite PGS analysis

To assess the merits of our clustering approach, we additionally trained models simply predicting SCZ in Train_FinnGen_ with the same PGS features as predictors as in the primary machine learning analyses (see above). Predictions were projected onto the test folds in a 10-fold CV scheme, and patients were then classified into two groups along the median of the predicted SCZ probabilities (i.e the fraction of trees in the models that classified a given individual as having SCZ). These classifications served as a comparison to the GRPs. Additionally, to assess whether the multi-PGS prediction model for SCZ outperforms SCZ-PGS alone, we conducted analyses aimed at optimal overall predictive performance rather than patient stratification (see Supplementary Methods 1.4.2).

### Feature selection

Feature selection aimed at identifying the PGS that showed the strongest association with diagnosis in order to improve the clustering performance. To achieve this, the PGS of 413 GWAS were ranked according to their relevance to SCZ. From that ranked list, the 45 most SCZ-relevant PGS (ranked 1–45) were included in the ML analysis. We selected 45 as the maximum feature size to evaluate GRP formation at larger feature sizes and to demonstrate that GRP assignments stabilize beyond a certain point. To prevent data leakage during cross-validation in the training dataset, feature selection was performed separately for each cross-validation iteration. For validation in the independent datasets, features were extracted from the entire training dataset (Train_FinnGen_). In addition to this univariate feature selection approach, we repeated the analyses using multivariate feature-selection methods (LASSO and sparse-group LASSO) as a sensitivity analysis to investigate whether selecting features in a univariate manner biased the resulting genetic profiles and downstream analyses. We evaluated the results of these analyses by calculating the ARI between the cluster solutions obtained using the multivariate approaches and those derived from our primary approach (see Supplementary Methods 1.5.1).

### Formation of the PGS categories

As many of the top 45 selected features were conceptually similar (e.g., different neuroticism items) we set out to investigate if they could be grouped empirically. The first step in forming the PGS categories was to calculate genetic correlations using LDSC software[39, 40] for the top 45 GWAS. The European samples reference panel from the 1000 Genomes project[41] was used for LD scoring. Then, hierarchical clustering was performed on the absolute values of the resulting (quasi-) correlation matrix. Upon visual inspection and theoretical consideration, a four-cluster solution was selected (see Supplementary Figures 2, 3). The first group includes SCZ itself, bipolar disorder (PGC 2019), openness (Big five personality traits 2024) and two alcohol traits. At smaller feature sizes (e.g., 12–16 top-ranked features), the grouping of GWAS based on genetic correlation remained unchanged (see Supplementary Figures 4-7).

### Characterizing the genetic risk profiles

#### Investigating PGS profiles for each PGS category

This step aims to characterize the GRPs by calculating their first PC for each PGS category using all PGS within a PGS category. All PGS of a PGS category for HCs and patients uniquely assigned to each GRP were combined together into a data table to calculate PCs. As SCZ was the primary phenotype of interest, for each GRP, the mean SCZ-PGS was presented to represent this group. To present the other three groups (cognition, depression, and neuroticism), for each GRP, the mean of the first PCs of the respective PGS category across feature sizes were plotted. The mean values of GRPs were adjusted such that HC were centered at zero (Figure 2). Patients predicted as SCZ by both models or by neither model were excluded from the primary analysis (see Results for the corresponding proportions). To assess potential selection due to these exclusions, we repeated the analysis by assigning all patients to GRPs according to the prediction model that yielded the higher probability of case status (see Supplementary Methods 1.5.2).

**Figure 2.**
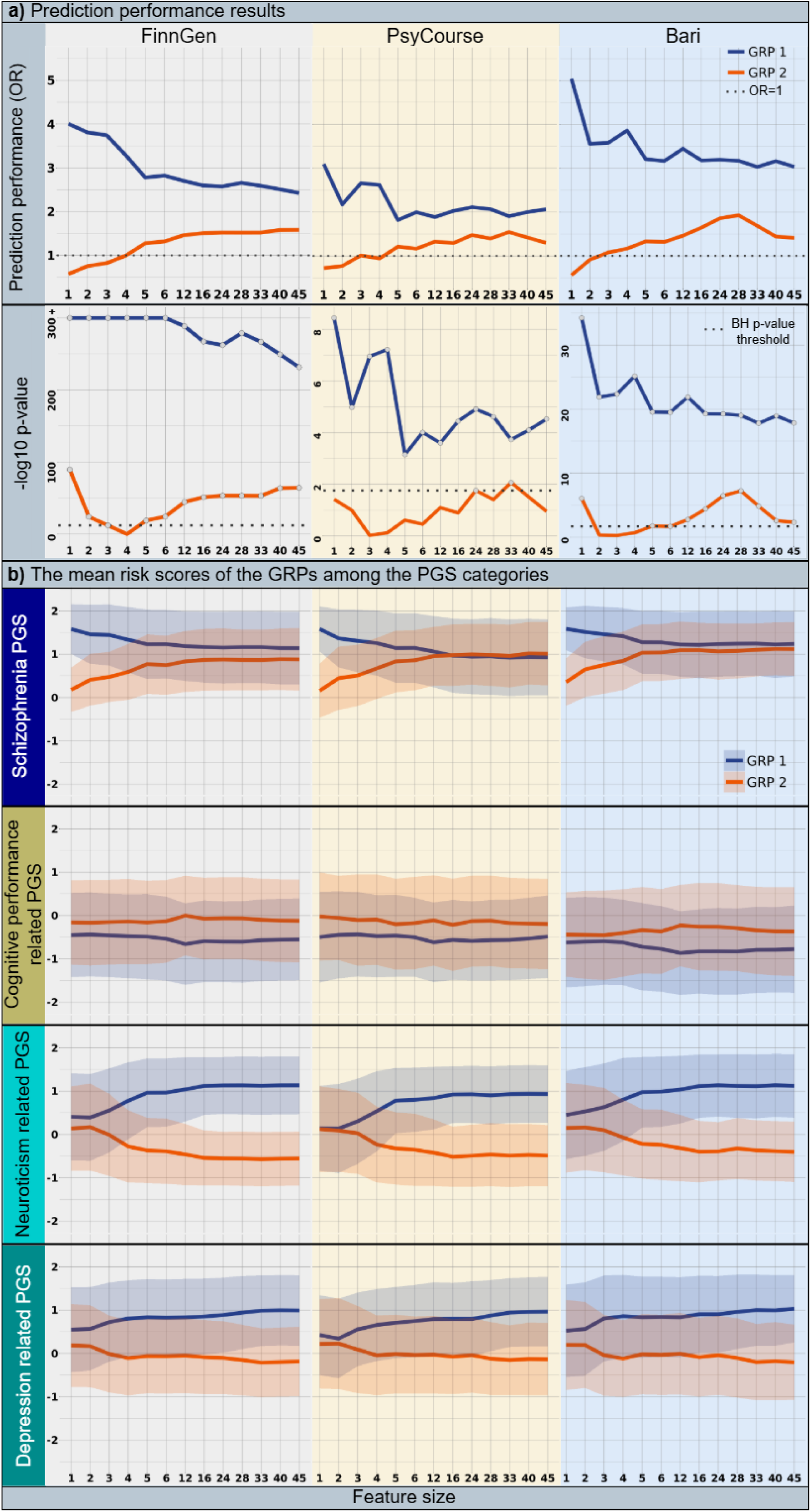
Results of GRP validation. **a)** Associations between the predicted cluster memberships and diagnoses were quantified as odds ratios and significance were determined using two-sided Fisher’s exact test where the input was a cross table of either GRP 1 or GRP 2 model and SCZ. The raw p-values were -log10 transformed to increase the readability of the plot. Feature sizes from 1 to 45 (x-axis) were plotted to demonstrate the formation with increasing feature size. An OR larger than 1 (dashed line) means that a prediction of having SCZ by a given model increases the odds of truly having SCZ. The dashed lines on the p-value plots show the cutoff for BH-corrected significance, with circle symbols showing traits that were significant after correction (number of tests = 26). **b)** The PGS compositions of the GRPs were illustrated for the three PGS categories (neuroticism, depression, and cognition-related PGS categories) and for SCZ-PGS. The mean of the first PC (to ensure comparability across units of analysis, all measures were scaled) of each PGS category was plotted across the feature sizes from 1 to 45 (x-axis) for GRP 1, GRP 2. The mean values were adjusted such that HC were centered at zero. Lines represent the mean values, and shaded ribbons indicate ±1 standard deviation.

#### Investigating PGS profiles for each PGS

This step aims to characterize the GRPs by estimating the associations between selected PGS and the GRPs using logistic regression. HCs and patients uniquely assigned to each GRP were sampled to create dependent variables of logistic regression. Logistic regression was then applied separately for each PGS of the top 45 GWAS to associate the GRPs as displayed in the formulas (1, 2).

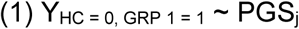

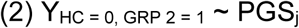

PGS_j_ is the best PGS of j^th^ GWAS of the top 45.

The beta values calculated by the logistic regression represent the association between the PGS_j_ and the GRP assignments (Figure 3).

**Figure 3.**
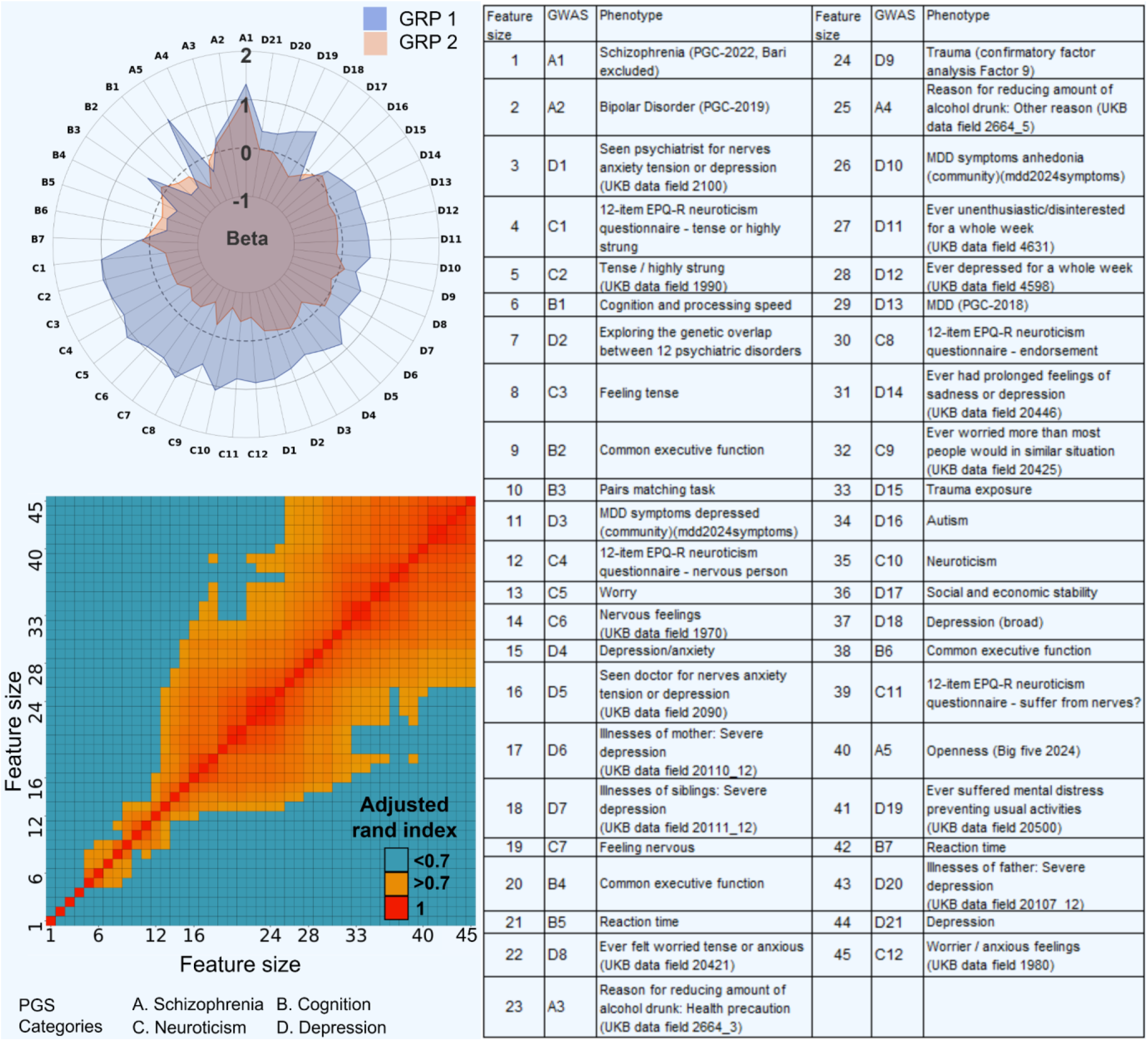
PGS composition of the GRPs and stability across the feature sizes. The polar plot was generated using the FinnGen cohort’s association results for the feature size 45. To enable comparison across PGS categories, the PGS categories were plotted sequentially. The legend (in a form of a table) of the polar plot shows the ranked order of the 45 most SCZ-relevant PGS. The beta coefficients obtained from logistic regression represent the associations between each PGS and GRP relative to HC. Adjusted rand index (ARI) comparisons were generated using the FinnGen cohort’s k-means clustering results and plotted as heatmap. ARI was calculated directly using the whole sample prediction results. To enable comparison across features, 0.7 is selected as the threshold for high similarity. Detailed information about the 45 GWAS listed in the table can be found in Supplementary Table 27. The values used to create the polar plot can be found in Supplementary Table 28. The ARI values used to generate the heatmap can be found in Supplementary Table 29.

#### Investigating cluster stability

The k-means clustering solution for a given feature size was compared with the clustering results of all other feature sizes and visualized as a heatmap (Figure 3). Cluster similarities were quantified using the adjusted Rand index [42]. To address potential bias introduced by correlated PGS in the k-means clustering, we performed a sensitivity analysis in which the original PGS were replaced by principal components derived within each PGS category, thereby reducing feature collinearity (see Supplementary Methods 1.5.3).

#### Investigating phenotypic profiles

To compare the phenotypic profiles of the two GRPs, we used a general linear model (GLM) regressing the phenotypic variable j on the GRP (n = either 1 or 2), with the SCZ-PGS and available demographic data used as covariates using (3):

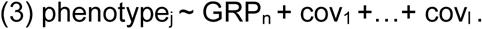

Continuous phenotypes were initially z-standardized 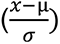. R version 4.4.1 was used for all statistical analyses. The glm function from the standard stats package was used to apply the GLM as displayed in (3). In the case of binary phenotypic variables, a logistic regression with the same covariates was performed and odds ratios are reported. As the two groups most representative of each GRP are the SCZ patients uniquely correctly identified by either of the prediction models, we used these groups for the primary comparisons, thereby excluding SCZ patients for whom both models agreed. As a sensitivity analysis, we performed the same comparison analyses with each SCZ patient assigned to one of the GRPs based on which of the two models predicted case status with the higher probability.

#### Comparing the predictive performance of pleiotropic and SCZ-specific SNPs

To gain insight into why one GRP model predicted SCZ better than the other, despite similar overall polygenic risk for SCZ, we further decomposed the polygenic risk [43, 44], by comparing the contribution of genomic loci shared between the SCZ GWAS and the other three PGS categories versus SCZ-specific loci. We calculated a corresponding subset SCZ-PGS using the SNPs with conjunction FDR<0.1 to ensure a sufficient number of SNPs (see Supplementary Table 11) for each of the PGS categories, and weighting the SNPs by their association in the SCZ GWAS. To achieve this, we first conducted the conjunctional FDR[37, 38] approach between SCZ GWAS and all the other GWAS among the top 45. If a SNP had a conjFDR of below 0.1 with any of the GWAS within a PGS category, it was included in the calculation of the subset SCZ-PGS of that PGS category. For example, for the PGS category “Cognition” a corresponding subset PGS was constructed, capturing the genetic variation relating to both SCZ and cognitive traits (i.e. pleiotropic effects). For each of the three SNP subsets 100 p-value matched SNP sets (as indicated in the SCZ GWAS) of equal size were sampled from the SNPs outside of the corresponding pleiotropic SNP set. The matching was performed with the R function *matchit* (distance = “glm”) from the package *MatchIt*[45]. Different matched subsets were achieved by adding slight random error to the p-values against which the matching was done.

After defining the SNP sets, PGS were calculated for each of them using PRSice-2[46]. Their predictive performance was assessed with logistic regressions where SCZ was the outcome and a given subset PGS the predictor. Before the analyses, all the PGSs were adjusted for the first 20 ancestral PCs and then standardized. Finally, Cochran’s Q test (function *rma* with method = “EE”, from the R package *metafor* [47]) was used to determine a p-value for the difference in effect size between a subset PGS of a given PGS category and the best corresponding comparative PGS.

Additionally, to assess whether the difference in predictive performance was driven by the slight difference in their mean SCZ-PGS, we repeated the analyses using patient training clusters matched for SCZ-PGS (see Supplementary Methods 1.5.4).

## Results

The PRScope procedure was applied to data from the Train_FinnGen_ to identify two data-driven clusters of SCZ. This *a priori* choice of a two-cluster solution was supported by the observation that the largest reduction in within-cluster variation occurred when increasing the number of clusters from one to two (see Supplementary Figure 8). Separate ML models were then trained to characterize each of these clusters as GRPs and enable their prediction in independent test data. The predictive value of these cluster models, and their dependence on the number of included PGS, was first tested in Train_FinnGen_ using a cross-validation approach (Figure 2a, for uncorrected p-values see Supplementary Table 12). Subsequently, these models were applied to the independent test data, illustrating that GRPs could be successfully validated at the selected feature sizes (Figure 2a).

Consistent with the Train_FinnGen_ cross-validation results, GRP 1 was significantly positively associated with SCZ in Val_Bari_ at all evaluated feature sizes, whereas GRP 2 was significant from feature size 5 onward. In Val_PsyCourse_, GRP 1 was also significant at all evaluated feature sizes, whereas GRP 2 met the Benjamini–Hochberg (BH)-corrected significance criterion only at feature sizes 24 and 33 (see Supplementary Tables 13 and 14 for the raw p-values). The proportion of SCZ cases predicted by both models in Train_FinnGen_ ranged from 0.63% to 6.24% across the feature sizes. It was generally lower for smaller feature sizes and higher for larger feature sizes. The proportion of SCZ cases not predicted by either model ranged from 16% to 18% in Train_FinnGen_ and remained relatively uniform across the feature sizes. Consistent results were observed in the validation datasets, although the exact proportions varied across datasets (see Supplementary Tables 15-17). The GRPs did not differ significantly in terms of age or sex in any of the three datasets.

With increasing feature size, the composition of the GRPs changed (Figure 2b), and we observed a substantial reallocation of patients, which started leveling off with approximately feature size 12 (ARI heatmap in Figure 3). Thus, in order to test clinical associations with the GRPs, we decided to predict the GRPs at feature sizes (1-6, 12, 24, 33, 45) that represent the points of a high rate of reallocation into two validation datasets (see Supplementary Methods 1.4.3).

The sensitivity analyses using multivariate feature-selection approaches resulted in unstable feature sets across cross-validation folds. The traits selected by these approaches showed only partial overlap with the primary univariate approach (see Supplementary Tables 18, 19). However, the traits that reflect the essence of the PGS categories and characterize the GRPs were also selected by these methods and the predicted GRPs showed substantial similarity to the primary GRPs (LASSO; ARI: 0.62, OR_GRP 1_: 3.21, OR_GRP 2_: 1.69, feature size: 36. Sparse-group LASSO; ARI: 0.64, OR_GRP 1_: 3.12, OR_GRP 2_: 1.82, feature size: 41). The PCA-based clustering approach resulted in GRPs that were nearly identical to the primary solution (ARI: 0.87, OR_GRP 1_: 3.41, OR_GRP 2_: 1.28, feature size: 24). The details are presented in Supplementary Table 20. PGS compositions can be found in Supplementary Figures 9-11.

### The contribution of neuroticism, depression and cognitive performance-related predisposition to SCZ susceptibility is mediated by shared genetic variation

The top 45 selected features were grouped using pairwise genetic correlation analysis of the corresponding GWAS, resulting in 4 categories of PGS which can be characterized as relating to SCZ, cognition, depression and neuroticism. Figure 2 illustrates that the two GRPs were (at higher feature sizes) defined by a divergence in these PGS categories while their overall SCZ risk was very similar. Specifically, GRP 1 showed markedly elevated neuroticism and depression related PGS and relatively low cognition related PGS, whereas GRP 2 displayed an opposite pattern for neuroticism and depression related PGS and intermediate levels (between GRP 1 and HC) for the cognition related PGS.

The composite PGS approach shows a somewhat similar pattern of genetic risk, with the predicted high risk group showing higher levels of SCZ, depression and neuroticism, and lower cognition PGS. However, the differences are markedly smaller except for SCZ-PGS, for which it is much larger (see Supplementary Figure 12). The multi-PGS model showed a higher AUC than the SCZ-PGS only model in all 100 train/test splits (mean paired difference = 0.0106, 95% CI: 0.0103-0.0110, p < 10^-15^, see Supplementary Figure 13).

Since SCZ has well-established genetic correlations with neuroticism, depression, and cognition[21, 48], we investigated whether these aspects were collectively reflected as disproportional risk contributions of pleiotropic SNPs. For this, we constructed a subset PGS based on SNPs that were associated with SCZ and each of the PGS categories, using conjunctional FDR[37, 38] (conjFDR). All SNP weights were taken from the SCZ summary statistic. We could show that all three of these subset PGS predicted SCZ better than PGS constructed of p-value matched (as indicated in the SCZ GWAS) SNPs outside their respective pleiotropy SNP group (Figure 4). Supplementary Tables 21-23 show the pleiotropic variants as identified by conjFDR.

**Figure 4.**
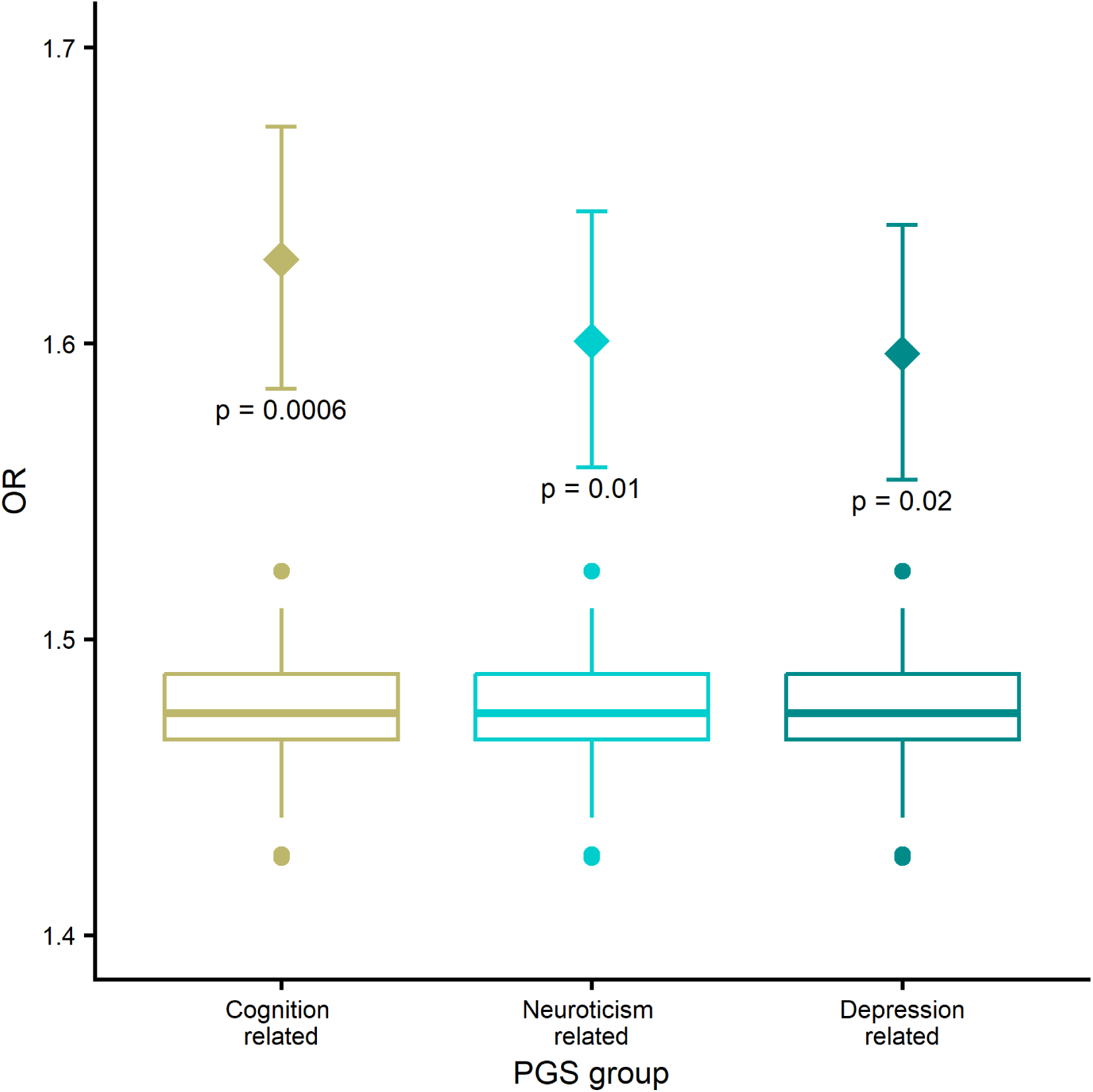
Predictive performance of the conjFDR-based PGS in Train_FinnGen_. The diamonds indicate the odds ratios for a PGS constructed with conjunctional FDR with a given PGS category (see Methods) in a logistic regression where true SCZ status was the dependent variable. The boxplots represent estimates from similar models with different PGSs serving as predictors. Each PGS was constructed on a p-value-matched random sample of SCZ risk SNPs. The p-values are derived from Cochran’s Q test of heterogeneity of effects, where the maximum estimate and the corresponding standard error from the 100 respective comparative PGSs are contrasted with the conjFDR PGS.

Even after eliminating differences in SCZ-PGS mean values by matching the clusters for SCZ-PGS, GRP 1 still showed better predictive performance than GRP 2 and the composition of the GRPs remained unchanged (see Supplementary Figures 14, 15 and Supplementary Table 24).

#### Genetic risk profiles have transdiagnostic relevance and differ in clinical measures

To clinically characterize the identified GRPs, differences in clinical features were assessed across the investigated datasets. All phenotypic results are shown for feature size 24 only for simplicity of illustration (for other feature sizes see Supplementary Figures 16-30). In Train_FinnGen_, GRP 1 was associated with a more severe and complex clinical manifestation in patients with SCZ as indicated by higher rates of antidepressant prescriptions and comorbid psychiatric diagnoses; including depression, neurotic (F40-F48 in ICD10), personality, and eating disorders (Figure 5). Importantly, the composite PGS comparison showed no significant differences in comorbidity rates (Supplementary Figures 31-41, and Supplementary Table 25 for group sizes).

**Figure 5.**
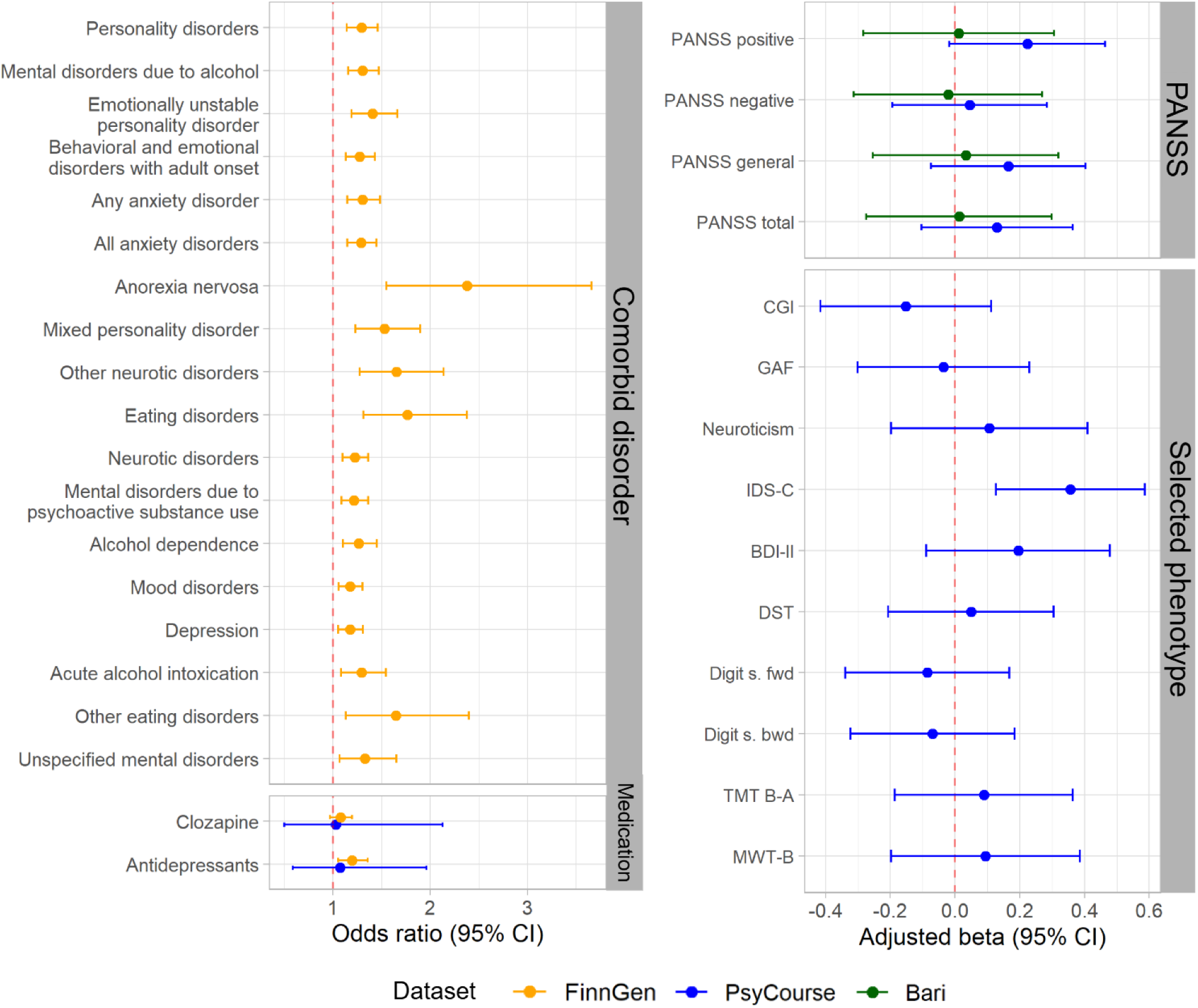
Phenotypic profiling of GRP 1 and GRP 2. Feature size = 24. Baseline: GRP 2. All comparisons are adjusted for the SCZ-PGS, sex, and age. In Val_PsyCourse_ and Val_Bari_ additionally adjusted for center/study; all results with the 95% CI (displayed) not overlapping the null effect are also significant with an FDR of 0.05 applied within each category (for comorbid disorders only the significant ones are shown); analyzed case numbers of GRP 1 and GRP 2 are listed in brackets (n_GRP 2_ / n_GRP 1_): Clozapine prescription (105/88), Antidepressants prescription (105/88), PANSS = Positive and Negative Syndrome Scale, positive, negative, general and total score (Val_Ps_yCourse 108/108, Val_Bari_ 89/95). CGI = Clinical global impression (109/108), GAF = Global Assessment of Functioning (108/108), Neuroticism = Dimension from Personality (87/76), IDS-C_30_ = Inventory of depressive symptomatology (109/103), BDI-II = Beck Depression Inventory-II (98/93), DST = Digit- Symbol-Test (105/103), Digit s. fwd = Verbal digit span forward (108/105), Digit s. bwd = Verbal digit span backward (108/105), TMT B-A = Trail making test part B - part A (105/97), MWT-B = Multiple- Choice Vocabulary Intelligence Test (74/74). For all Train_FinnGen_ analyses, the corresponding group sizes are (2696/3077). PsyCourse variable names can be found in Supplementary Table 30. The exact ICD codes and ATC drug codes of Train_FinnGen_ can be found in Supplementary Table 31. The phenotype codes for comorbid disorders in the FinnGen dataset can be found in Supplementary Table 32.

Both in Val_PsyCourse_ and Val_Bari_ the Positive and Negative Syndrome Scale (PANSS) showed no significant differences in any of the scales (Figure 5). In Val_PsyCourse_ we further investigated a collection of phenotypic variables representing the major risk components of the GRPs as represented by the PGS categories defining them. We found a significantly (FDR corrected p-values < 0.05) higher score for GRP 1 in the Inventory of depressive symptomatology (IDS-C_30_) (Figure 5). As statistical power varied depending on the feature size and number of missings in the validation datasets, we displayed the results of the post-hoc power analyses for all variables in Supplementary Table 26. In all datasets, the sensitivity analysis using a group assignment method that assigns all cases, i.e. each individual to the GRP with the highest probability, did not yield significantly different results from those of the primary analysis, demonstrating the robustness of the primary method (see Supplementary Figures 27-30 and 42,43). In Train_FinnGen_, GRP 1 model predictions were also associated with BIP (n cases = 7 555; n controls = 11 198) and MDD (n cases = 27 187; n controls = 26 993) in a gradient-dependent manner, with the highest OR observed for SCZ, intermediate for BIP, and the lowest for MDD. The corresponding associations for GRP 2 model predictions followed a slightly different pattern, where the associations with both BIP and DEP were negative, with no consistent difference in magnitude across feature sizes (see Supplementary Figure 44).

## Discussion

Here, we aimed to capture distinct GRPs within the SCZ patients through the simultaneous investigation of patients’ susceptibility to psychiatry-relevant phenotypes using 413 publicly available GWAS, leveraging a massive training dataset (Train_FinnGen_). We asked whether finding two genetically distinctive clusters of SCZ patients might help to identify phenotypically distinct and clinically relevant subgroups and lead to a better understanding of the genetic risk architecture of SCZ. This resulted in two distinct GRPs with similar SCZ risk; one GRP was characterized by high PGS of phenotypes relating to neuroticism and depression and low PGS for cognitive traits (GRP 1), whereas the other GRP (GRP 2) had a profile similar to HC in those dimensions. While the GRPs exhibited approximately the same elevated levels of genetic liability for SCZ, a random forest ML model based on GRP 1 clearly outperformed a model based on GRP 2 in predicting SCZ in Train_FinnGen_ and in independent validation datasets (Val_PsyCourse_ and Val_Bari_). Furthermore, patients with a GRP 1 tended to have clinical manifestations that indicate higher rates of disease complexity (psychiatric comorbidity), a clinical profile which does not show up when grouping the patients along their SCZ-PGS alone. Interestingly, the two groups did not differ with regards to phenotypic measures which are commonly used to stratify SCZ patients (e.g., PANSS score / subscores), indicating their potential value beyond a phenotypical or clinical assessment of patients. While previous studies have demonstrated that disease prediction can be improved with a multi-PGS approach[26], this is the first time it has been applied to data-driven clustering of patients. The observation that the model based on GRP 1 is a considerably better predictor of SCZ than the one based on GRP 2, despite the similar SCZ risk, has several potential explanations that are not mutually exclusive. First, the genetic risk contributions for neuroticism, depression-related traits, and low cognitive performance may reflect biological mechanisms facilitating the actualization of disease risk. Second, the corresponding genetic risk may be only partially captured by SCZ-PGS. Third, SCZ genetic risk estimation may also be more accurate in GRP 1 than in GRP 2 patients. In this scenario, the higher levels of polygenic risk of non-SCZ traits/disorders in GRP 1 would suggest a higher prevalence of alleles that confer risk to both SCZ and other related traits. These pleiotropic SNPs could be more often truly causal for SCZ than the ones only associated with SCZ risk, as the additional associations can be considered as an increase in the strength of statistical evidence. Our finding that the PGS based on conjFDR SNPs are slightly better predictors than PGS based on comparable subsets of SCZ risk SNPs lends support for this. Finally, phenotypic mediation, such as neuroticism being associated with higher stress levels, which in turn increase the risk of developing SCZ, could play a role[49, 50]. Another concern regarding the better predictive performance of GRP1 was that it could have been driven by the slightly higher SCZ-PGS in GRP 1 compared to GRP 2. However, this performance difference persisted in the sensitivity analysis using clusters matched for SCZ-PGS.

The current study has several limitations. First, a methodological limitation of the univariate feature selection was that it assigned high rank to multiple correlated traits providing redundant information. To address this limitation, we evaluated multivariate feature-selection methods, which resulted in unstable trait sets when predictors were correlated and had small effect sizes. Furthermore, correlated traits with potential biological relevance to schizophrenia might be disproportionately penalized by multivariate methods and therefore not selected because of their small effect sizes. By contrast, univariate feature selection evaluated each trait independently and therefore yielded a stable, albeit partially redundant, trait set in our large-scale discovery cohort. Importantly, reducing redundancy among correlated PGS using a PCA-based approach produced similar results, suggesting that the primary findings were not driven by this redundancy. In addition, retaining correlated traits may accommodate cohort-specific differences, as PGS derived from different GWAS of similar traits may vary in their predictive value across validation datasets. Taken together, these observations support univariate feature selection as a pragmatic strategy for clustering in highly heterogeneous disorders such as schizophrenia, as it allows the analysis to focus on the most disorder-relevant traits and identify more genetically homogeneous groups. Nevertheless, the sensitivity analyses suggested that this methodological limitation did not bias the estimation of the effects associated with each GRP in the downstream analyses. Second, as one of the main goals of this study was reproducibility, and thus considering the sample size limitations of the validation datasets, we restricted the analysis to classify patients into two GRPs (a choice for k = 2 supported by the elbow method, see Methods). This two-cluster solution will likely not capture the entire complexity of SCZ risk mediated by common variants. It should also be noted that the observed divergence in GRPs can arise from the correlation structure of the input variables to the clustering algorithm, even without the existence of truly separate clusters. Third, our method only considered genetic predisposition to psychiatry-related traits. It is possible that the inclusion of genetic risk in somatic domains would improve the classification and provide more insight into underlying biological mechanisms[51–53]. Fourth, the heterogeneity between the datasets we used in terms of the availability of phenotypic data hampered us from conducting more harmonized analyses in that regard, drawing less definite conclusions about generalizability. On the other hand, the fact that our procedure works across such different datasets can be seen as an indication of good portability.

Finally, as only samples of European descent and GWAS from that ancestry were used, the findings presented in this paper may not generalize to individuals with non-European ancestry[54, 55].

In summary, our data-driven ML scheme successfully leveraged a large number of psychiatry-related PGS, for the first time, to derive two GRPs of SCZ patients. Potential next steps to further characterize the identified signal include relating the GRPs to additional biological and clinical measures, for example structural MRI measures, and outcomes in longitudinal datasets, such as psychosis onset, diagnostic transition, treatment response, and relapse. As the GRPs were not associated with common state-dependent clinical measures (e.g., PANSS scores), they may provide complementary information beyond current symptom presentation. This could ultimately support treatment personalization, a goal which stratification based solely on clinical measures so far failed to accomplish [56, 57]. A prediction model based on GRP 1, characterized by a high liability for neuroticism, depression related traits and low cognitive functioning, is a promising step toward identifying SCZ patients with a poor prognosis at an early stage. One potential application is the combination of GRP models with established clinical predictors in multi-modal prediction models.

Randomized controlled trials could be used to evaluate whether GRP-informed risk stratification models improve personalized preventive interventions among individuals with psychosis risk syndrome [58, 59]. Despite the potential usefulness of GRP 1 in future clinical research, we note that it should not be currently considered as a clinical subtype of SCZ.

Taken together, this work establishes a framework for integrating multi-phenotype PGS with ML approaches for the genetic stratification of complex disorders. While demonstrated here in SCZ, the framework can be applied to other psychiatric and somatic conditions.

## Code Availability

The code used to generate PGS and identify GRPs is implemented in PRScope and is publicly available at https://github.com/transbioZI/PRScope.

## Data Availability

For FinnGen, researchers can apply for health data from the Finnish Data Authority Findata (https://findata.fi/en/permits/) and individual-level genotype data available through the Fingenious portal (https://site.fingenious.fi/en/). These resources are hosted by the Finnish Biobank Cooperative FINBB (https://finbb.fi/en/). Access can only be provided for research projects within the scope of the Finnish Biobank Act, which includes health promotion, understanding disease mechanisms or developing medical products or treatment practices.
PsyCourse data is available to bona fide reseachers upon a submission and approval of a research proposal.
Individual genotype and clinical data from the University of Bari Aldo Moro with demographic and behavioral characteristics cannot be shared at individual level in raw format because of ethic restrictions.

http://www.psycourse.de/openscience-de.html

https://docs.finngen.fi/

## Acknowledgments

We thank all research participants and all researchers and clinicians who collected, generated, or processed the data used in this study.

