## Supplementary materials for "Development and validation of polygenic risk profiles of schizophrenia"

8, Lieber Institute for Brain Development, Johns Hopkins Medical Campus, Baltimore, MD, USA
9, International Max Planck Research School for Translational Psychiatry (IMPRS-TP), Max-Planck-Institute of Psychiatry, Munich, Germany
10, Department of Immunology, Faculty of Medicine, Ahvaz, Iran
11, Department of Psychiatry and Psychotherapy, LMU University Hospital, Munich, Germany
12, Department Clinical Translation, Max Planck Institute of Psychiatry, Munich, Germany, Munich, Germany
13, Department of Psychiatry and Psychotherapy, Faculty of Medicine and University Hospital Bonn, Bonn, Germany
14, Institute of Human Genetics, Faculty of Medicine and University Hospital Bonn, Bonn, Germany
15, German Center for Mental Health (DZPG), partner site Munich/Augsburg, Germany
16, Centres for Psychiatry Suedwuerttemberg, Centres for Psychiatry Suedwuerttemberg, Ravensburg, Germany
17, Department of Psychiatry and Behavioral Sciences, Johns Hopkins University School of Medicine, Baltimore, MD, USA
18, Department of Genetic Medicine, Johns Hopkins University School of Medicine, Baltimore, MD, USA
19, Department of Psychiatry and Behavioral Sciences, SUNY Upstate Medical University, NY, USA
20, German Center for Mental Health (DZPG), Partner Site Mannheim - Heidelberg - Ulm, Germany

* Contributed equally
### Joint senior authors

Corresponding author
Prof. Emanuel Schwarz, PhD

Address: Hector Institute for Artificial Intelligence in Psychiatry (HITKIP), M7 68161 Mannheim, Germany

### 1. Supplementary Methods

#### 1.1. Detailed cohort descriptions

##### 1.1.1. The PsyCourse Study

PsyCourse is a multi-center observational longitudinal deep-phenotyping study of the affective-to-psychotic spectrum, including healthy controls. All participants provided written informed consent, and the study was approved by all relevant Ethics Committees (see [1]). Demographic covariates used for the phenotypic profiling in $Val_{PsyCourse}$ were sex, age, and center of recruitment (in addition to the SCZ-PGS on all p-value threshold levels used in all datasets). Due to the large number of recruitment sites (n=20), and the unequal distribution of patients recruited across these sites, we focused on sites that recruited at least 20 patients (resulting in a selection of n = 419 SCZ patients and n=299 HC after post-QC ). Due to the longitudinal study design, some of the phenotypic measures are taken at up to four time points (spread over 18 months) for each subject. If this was the case, we used the maximum value over the time points. Due to the sparse pattern of missing values in the PsyCourse dataset, the number of available data varies depending on the phenotypic measure. Based on the results from $Train_{FinnGen}$, we compared the number of patients taking clozapine or antidepressants between the GRPs (this analysis was run without controlling for center, due to the low number of cases from each center). The information about the clozapine and antidepressant treatment was derived from the medication dataset, a subset of the PsyCourse data set (see PsyCourse codebook v5.0). In addition to this we analyzed phenotypic measures based on PGS defining the GRPs which could be summarized as SCZ (Positive and Negative Syndrome Scale (PANSS): positive, negative and general symptoms subscale and total score) and three additional major components, which genetic predispositions mainly drive the difference between the GRPs: Cognitive function (verbal digit span forward, verbal digit span backward, Digit-Symbol-Test (DST), Multiple-Choice Vocabulary Intelligence Test (MWT-B), TMT B-A = Trail Making Test part B - part A), depression-related traits (Inventory of depressive symptomatology (IDS-$C_{30}$), Beck Depression Inventory II (BDI-II)) and neuroticism (Neuroticism = Dimension from BFI-10 questionnaire [2]). On top of these, we also added measures of overall disease severity (Clinical global impression (CGI), Global Assessment of Functioning (GAF)). Variable names as used in the PsyCourse codebook: https://data.ub.unimuenchen.de/251/1/210908_PsyCourse_v5.0.html are mapped in Supplementary Table 30. The resulting p-values were separately FDR-corrected for each analysis block (medication, PANSS scales, additional phenotypic variables).

##### 1.1.2. Bari

Demographic covariates used for the phenotypic profiling in $Val_{Bari}$ were sex, age and the original study in which the data was acquired (in addition to the SCZ-PGS on all p-value threshold levels used in all datasets). The Bari dataset consists of data from two studies. Subjects with European ancestry, comprising both healthy controls and individuals diagnosed with SCZ or schizoaffective disorder, were recruited from the Apulia region in Italy. The recruitment process followed the ethical standards outlined in the World Medical Association’s Declaration of Helsinki and was approved by the local ethics committee (“Comitato Etico Indipendente Locale - Azienda Ospedaliero-Universitaria Consorziale Policlinico di Bari”). Healthy controls were screened using the Non-Patient Structured Clinical Interview for DSM-IV to ensure they were unaffected by any psychiatric condition. Diagnosis of SCZ was made using the Structured Clinical Interview for the DSM-IV, Axis 1 disorders by board-certified psychiatrists. Participants were excluded if they had a significant history of drug or alcohol abuse, active substance use within the past year, a history of head trauma with loss of consciousness, or any other serious medical condition. Clinical phenotypic data in this dataset was the PANSS negative sum, PANSS positive sum, PANSS general sum, and PANSS total sum, which was available for 249 of the SCZ patients. Other phenotypic data was also available but for only a very small number of SCZ patients. Analogous to the approach in the PsyCourse dataset we used the max value whenever values for more than one time point were available. The resulting p-values were separately FDR-corrected for the PANSS scales.

##### 1.1.3. FinnGen

FinnGen is a public-private partnership project between Finnish research institutes and biobanks and private pharmaceutical companies. It leverages comprehensive Finnish digital health care registers combined with genotypic data from 500k individuals. The diagnoses in the registers are based on ICD-10, ICD-9 and ICD-8 criteria. To examine GRP differences in comorbidity of psychiatric phenotypes each phenotype was analyzed with a separate logistic regression with GRP as a dichotomous predictor and

SCZ-PGS, age, and sex as covariates. Endpoints with less than 100 cases among the SCZ patients were excluded. Benjamini-Hochberg multiple-testing correction was applied within a given feature size at a false discovery rate of 0.05. The same procedure was applied for the medication analyses (see chapter 1.1.3.1 below for how medication history was determined).

###### 1.1.3.1. Phenotype definitions in FinnGen

SCZ, BIP, and MDD cases were selected using the specialized inpatient and outpatient, cause of death, and drug purchase registers (coded in the data as INPAT, OUTPAT, DEATH and PURCH, respectively). For each disorder, their respective ICD codes served as case criteria. Additionally, individuals who had ever purchased the drug clozapine were determined as having SCZ, as that drug is virtually never prescribed for other disorders. Lithium for BD or antidepressants for depression were not utilized to that end as they are more commonly prescribed for other disorders and diseases. The disorders were regarded as hierarchical in the way that SCZ superseded BIP which in turn superseded MDD.

For defining controls, more stringent criteria were applied. In the case of SCZ, all psychotic disorders lead to exclusion, as well as Olanzapine, Risperidone, Amisulpride and Haloperidol purchases. The aforementioned drugs indicate a psychotic disorder or episode, but not necessarily confirmed SCZ. For BIP, the same criteria applied with the addition of not having any mood disorders. Additional registers of primary health care (PRIM_OUT) and drug reimbursement (REIMB) were used for control exclusion to add more certainty that control subjects are truly not cases. The exact ICD codes and ATC drug codes can be found in Supplementary Table 31.

From the remaining eligible controls a sex and age-matched sample was taken (*matchit* function in the R-package *MatchIt[3]*) to simplify downstream analyses and to limit the number of individuals for whom PGSs needed to be calculated. The control group sizes were four times the number of cases for SCZ and BIP and of equal size for MDD. Finally, the full sample (cases of all disorder phenotypes and all control groups) was restricted to unrelated individuals so that the maximum allowed degree of relatedness was 3 (e.g. first cousins). 10,000 random selections of removal with a preference for keeping cases were performed and the one that resulted in the least subjects getting excluded was applied.

###### 1.1.3.2. Phenotypes for comorbidity and medication analyses

All psychiatry-related (category F5) FinnGen core endpoints served as phenotypes in the comorbidity analyses without further modifications. Core endpoints are the ones defined by the FinnGen core analysis team and their descriptions can be browsed at https://risteys.finngen.fi/. For the medication analyses, clozapine use was identified with the ATC code N05AH02, and antidepressants with the ATC category N06A as recorded in the drug purchase register.

#### 1.2. Genotype data imputation and pre-processing

##### 1.2.1. The PsyCourse Study genotype data

The pre-imputation genotype data provided by the PsyCourse study included 428,904 genome-wide SNPs (GSA chip) from 1,600 individuals diagnosed with BD, SCZ, schizoaffective disorder, schizophreniform disorder and HC. From this dataset, 26 related individuals were excluded, and SCZ patients (ICD-10 Schizophrenia and Schizophrenia according to DSM-IV-TR) and HC born in European countries (PsyCourse codebook variable : v1_cntr_brth) were selected, resulting in a final dataset of 755 individuals.

In the first QC-cycle of the imputation, Ricopili's pre-imputation module with default parameters was executed. Thus, 500 SNPs and one individual failing to meet the criteria were excluded from the dataset. In the next step, Ricopili's PCA module using default parameters was executed to identify the outlier subjects. 31 outlier subjects were identified and excluded that exceeded ±0.1 of the first two PCs. In the second QC-cycle, Ricopili's pre-imputation module with default parameters was executed again and 84 SNPs failing to meet the criteria were excluded from the dataset. The final pre-imputation dataset included 723 subjects and 428,320 SNPs.

As the final data met quality control criteria (lambda1000 = 1.014 and no genome-wide significant SNPs were found), imputation was performed using the 1000 Genomes European population as reference. Once the imputation was completed, SNPs with info scores above 0.8 (n = 4,883,969) were selected for post-imputation QC using our in-house genotype data pre-processing module.

The imputed and quality-controlled dataset consisted of 3,470,953 SNPs and 718 individuals.

##### 1.2.2. Bari genotype data

Genotype data from Bari was collected over four years: 2015, 2016, 2017, and 2019. Genotyping for all participants was conducted using the Illumina HumanOmni2.5-8 v1 BeadChip platform. From the pre-imputation genotype data, subjects with SCZ and HC were selected from all batches. SNP overlaps between batches were then compared, and those with the highest overlap were combined, and imputed together. Accordingly, the batches from 2015 (557 individuals, 1,258,881 SNPs) and 2019 (154 individuals, 1,454,515 SNPs) were combined and imputed together. Similarly, the batches from 2016 (139 individuals, 1,424,791 SNPs) and 2017 (501 individuals, 1,431,583 SNPs) were merged and imputed together.

###### 1.2.2.1. Dataset 1 (2015 together with 2019)

Pre-imputation data consisted of 1,197,491 overlapping SNPs between two batches and 711 individuals from Italy diagnosed with SCZ and HC.

In the first QC-cycle, Ricopili's pre-imputation module with default parameters (except minor allele frequency (MAF) > 0.02, because the lambda1000 value was high due to the genetic inflation of SNPs with MAF values lower than this.) was executed. Thus, 59,089 SNPs failing to meet the criteria were excluded from the dataset. In the next step, Ricopili's PCA module using default parameters was executed to identify the related and outlier individuals. In total, 34 subjects were excluded from the data which were identified as related (pi-hat > 0.2). The final pre-imputation dataset included 677 subjects and 1,138,402 SNPs.

As the final data met quality control criteria (lambda1000 = 1.022 and one genome-wide significant SNPs was removed), imputation was performed using the 1000 Genomes European population as reference. Once the imputation was completed, SNPs with info scores above 0.8 (n = 7,892,583) were selected for merging with the Dataset 2.

###### 1.2.2.2. Dataset 2 (2016 together with 2017)

Pre-imputation data consisted of 1,357,782 overlapping SNPs between two batches and 640 individuals from Italy diagnosed with SCZ and HC.

In the first QC-cycle, Ricopili's pre-imputation module with default parameters (except MAF > 0.02, because the lambda1000 value was high due to the genetic inflation of SNPs with MAF values lower than this.) was executed. Thus, 57,412 SNPs failing to meet the criteria were excluded from the dataset. In the next step, Ricopili's PCA module using default parameters was executed to identify the related and outlier individuals. In total, 21 subjects were excluded from the data which were identified as related (pi-hat > 0.2). The final pre-imputation dataset included 619 subjects and 1,300,370 SNPs.

As the final data met quality control criteria (lambda1000 = 0.979 and one genome-wide significant SNPs was removed), imputation was performed using the 1000 Genomes European population as reference. Once the imputation was completed, SNPs with info scores above 0.8 (n = 8,147,504) were selected for merging with the Dataset 1.

###### 1.2.2.3. Merging Dataset 1 and Dataset 2

Merging Dataset 1 and Dataset 2 resulted in a final dataset with 7,458,687 overlapping SNPs and 1,296 subjects. Post-imputation QC was performed using our in-house genotype data pre-processing module.

After completing the pre-processing module, the imputed and quality-controlled dataset consisted of 5,432,607 SNPs and 1273 individuals. That is, 23 individuals and 2,026,080 SNPs, which did not meet the criteria specified in the genotype pre-processing module, were removed from the post-imputation dataset. Four subjects whose deviations exceeded four times the standard deviation based on PCs 1 and 2 were excluded, resulting in a final sample of 1,269 subjects for analysis.

##### 1.2.3. FinnGen genotype data

Samples were genotyped using Affymetrix (ThermoFisher Scientific) and Illumina arrays.  Imputation was performed using a reference panel specific to the Finnish population (SISu v.4) based on 8,554 whole-genome-sequenced individuals resulting in 21 million imputed variants for each participant. Sample QC involved the removal of participants whose genetic sex did not match the ones provided in the registers, with a variant missingness of >2%, heterozygosity of more than 4 SD from the mean in common variant (MAF > 0.05) or excess relatedness to other samples (more than 500 relatives with PI_HAT > 0.1). Genetic variants were pruned out if their allele frequencies significantly differed from the reference panel (p < 5e-8) adjusted for the first 10 principal components, if they had HWE p < 1e-10 or 15% of the batches had a missing rate > 0.04. Additionally, variants were removed within a batch if HWE p-value was below 1e-6 or the missing rate was above 0.02. Each genotyping batch consisted of ~5000 participants.

#### 1.3. PRScope in detail

##### 1.3.1. Automated GWAS search module

In the present study, our strategy for ontology selection aimed at including a broad spectrum of phenotypes relevant to SCZ. For this, we focused on psychiatry-related ontologies from different domains such as biological, behavioral, brain development, psychiatric traits, and psychiatric disorders. In total, we selected 23 ontological categories (see Supplementary Table 4) and performed our GWAS search module to search the GWAS Catalog for GWAS summary statistics annotated with these ontological categories or their sub-categories. The resulting list included 348 GWAS summary statistics that met our criteria and had a harmonized version of the respective GWAS summary statistics. Using these harmonized versions of GWAS summary statistics led to two favorable outcomes: First, the GWAS summary statistics were transformed into a uniform format, enabling effective calculation of PGSs. Second, variants were already mapped to the latest genome assembly and alleles were orientated to the forward strand. Additionally, we found 1,462 relevant GWAS summary statistics that met our filtering criteria but did not have a harmonized version (see Supplementary Table 5). Furthermore, 65 GWAS summary statistics not listed in the GWAS Catalog but accessible from PGC, CTG, and other public sources (see Supplementary Tables 6-10), were prepared and included in our analysis using the same harmonization pipeline as applied by the GWAS Catalog. In total, the resulting list contains 413 GWAS.

###### 1.3.1.1. GWAS search criteria

The following filtering was used to include summary statistics in the analyses:

- European ancestry samples
- Total sample size > 10,000
- Summary statistics containing at least 250,000 SNPs
- The publication date is newer than January 1, 2018
- Harmonized version

##### 1.3.2. GWAS summary statistics QC module

Duplicate, ambiguous, and multi-allelic SNPs, as well as SNPs that lacked both beta value and odds ratio, were removed from the GWAS summary statistics. The p-value was set to 1 for SNPs without a p-value.

##### 1.3.3. Genotype QC module

First, SNPs and subjects were filtered based on a relaxed threshold (missing call rate of > 20%). Then a filter with a more stringent threshold was applied. Subjects with missing call rates of > 2% were removed. Next, SNPs with missing call rates of > 2%, minor allele frequency < 1%, and deviation from the Hardy-Weinberg equilibrium ($p \leq 1 \times 10^{-6}$) were removed. Additionally, non-autosomal, duplicate, and ambiguous SNPs were excluded. The R-package used to estimate relatedness was carried out as implemented in plinkQC v0.3.4[4], by estimating the relatedness between pairs of individuals based on identity-by-state similarity (PI_HAT > 0.1875) and retaining the subject with the lowest genotype missingness in pairs or larger groups of related subjects. Marees et al.[5] and more recently Choi et al.[6] have published detailed protocols for the calculation of PGS. These protocols include detailed instructions for performing quality control on genotype data and GWAS summary statistics, as well as standards for interpreting PGS. Our approach followed the guidelines presented by Marees, et al.[5] and Choi, et al.[6]. Detailed information about the imputation and pre-imputation can be found in the section *Genotype data imputation and pre-processing*.

PLINK v1.9 was used to assess the impact of population stratification via PC analysis as described in RICOPILI's PCA module[7]. In case the sample was bigger than 1,000 subjects, a subset of approximately 1,000 subjects was randomly selected for the pruning of the SNP set prior to the PCA. Similarly, if more than 1 million SNPs were present in the dataset, a random SNP set of ~1 million SNPs was selected for pruning. For the PCA calculation, we excluded SNPs with more than 2% missing call rates, a minor allele frequency below 5%, or a Hardy-Weinberg equilibrium deviation ($p \leq 1 \times 10^{-3}$), and SNPs in the long-range linkage disequilibrium (LD) regions of the Major Histocompatibility Complex region (chr 6, 25-35 MB) and the chromosome 8 inversion region (chr 8, 7-13 MB). The remaining SNPs were pruned using an r-squared threshold of 0.2, a window size of 200 kb, and a step size of 100 to minimize LD, followed by a second round of pruning with the same parameters. Finally, PLINK was used to calculate PCs for ancestry estimation for all subjects using the SNPs generated in the previous steps.

#### 1.4. Machine learning framework in detail

The goal of the applied ML analyses was to identify (using $Train_{FinnGen}$) and validate (using the $Val_{PsyCourse}$ and $Val_{Bari}$ data) clusters of patients in SCZ with differing multi-PGS profiles (i.e. 'GRPs'). Using the multi-PGS matrix we first determined the respective z-score of each PGS for patients with SCZ by subtracting the median of the controls and dividing by the standard deviation of the controls. To tune clustering to SCZ-relevant features, we then selected the PGS that showed the strongest association with the diagnosis (see Methods section *Feature selection*). After feature selection, clustering was performed on different numbers of selected PGSs (from top 1 to top 45) using k-means clustering (k=2, performed using the *kcca* function of the *flexclust* library)[8], to identify two clusters of patients. The maximum feature size of 45 was used, as at higher feature sizes no meaningful changes in cluster assignment were observed. The reason to use k=2 for the k-means clustering was to take into account the limited statistical power, due to sample size restrictions, especially in the deeply phenotyped validation datasets. While it is possible that k=2 does not produce an exhaustive result, the focus of our approach was on robustness and reproducibility, which in our opinion is a major weak point of more complex and exhaustive clustering approaches in the context of the limitation of data size in genomic psychiatric research. Clustering was repeated 100 times, the co-occurrence of subjects within a given cluster was recorded for each subject pair, and a consensus was identified by clustering the resulting matrix. Random forest ML[9] was applied to characterize the multi-PGS signature of each GRP and to predict this signature in independent data.

To characterize the PGS signature associated with each GRP, 51-times repeated random forest ML (using the ranger v0.17.0 R library with 1000 trees and default parameters)[10] was applied using under-sampling. Under-sampling was used to balance the unequal group sizes of patients within a given GRP, and the control group. To evaluate the reproducibility of this approach in the training data, the entire procedure was applied within a 10-fold cross-validation. For instance, to train the model for Cluster 1, the training folds comprised an equal number of HCs and SCZ patients. After training on the training folds, the Cluster 1 membership was predicted by the model in the test fold and associations between the predicted cluster memberships and diagnosis were quantified as the odds ratio (OR). The significance was determined using a two-sided Fisher's exact test. The same approach was applied to Cluster 2. The resulting p-values were used to find the rejection threshold of the BH procedure for controlling false discovery rate in multiple testing (number of tests: 26). The raw p-values and BH thresholds were -log10 transformed and plotted (Figure 2a). To prevent data leakage, the z-scoring and cluster identification were performed separately for each CV-iteration. To obtain comparable cluster assignments (as the cluster numbering is arbitrary) across CV folds, these were aligned to the first CV-iteration based on linear correlation obtained from predictions in overlapping training subjects. Predicted labels derived from cluster-specific models include both HC and SCZ subjects. In contrast, GRPs consist exclusively of SCZ patients uniquely identified by these models; therefore, the term GRP was adopted for characterization analysis.

##### 1.4.1. Determining the number of clusters (k in k-means)

Many methods exist to determine the k in k-means, with none of them being "the right one" [11]. As the focus of our approach was on robustness and reproducibility, we aimed to detect the smallest number of clusters that is still appropriate for our data. Thus, the number of clusters for k-means clustering was determined by plotting the within-cluster sums of squares (WSS) for different values of *k* at different feature sizes in the full FinnGen sample. The point at which the decrease in WSS slows down as *k* increases was interpreted as the optimal number of clusters. The k-means was performed with the base R function kmeans().

##### 1.4.2. Multi-PGS and SCZ-PGS only predictive performance

To assess whether the multi-PGS prediction model for SCZ outperforms SCZ-PGS alone, we conducted analyses aimed at optimal overall predictive performance rather than patient stratification. First, the FinnGen data was randomly split to tuning, training and testing sets with 3/5/2 ratios. In tuning sets (30% of the data), logistic LASSO regression with an inner 5-fold cross-validation was trained to tune the lambda parameter. The best lambda was then used to train another LASSO in the training set. Finally, the resulting model was used to predict SCZ in the test set. For the SCZ-PGS only comparison, a univariate logistic regression was trained in the same training set with SCZ-PGS as the predictor and SCZ diagnosis as the outcome. The test set performance was evaluated with AUC. The whole procedure was repeated 100 times with different tuning/training/test partitions of the data.

##### 1.4.3. Determining feature sizes for the full FinnGen data training

The number of source GWAS used for ML is denoted as *feature size*. To limit the number of models to be tested in the validation data sets ($Val_{PsyCourse}$ and $Val_{Bari}$), a few feature sizes were selected. This was achieved by first converting the cluster assignment table across all feature sizes to a distance matrix (function *dist* with the method option “binary”, from the R-package *stats [12]*) and then performing hierarchical clustering (function *hclust* from the R-package *stats [12]*) to determine feature sizes which give out similar cluster assignments. Upon visual inspection of the resulting dendrograms, 5 clusters (see Supplementary Figure 2) were deemed appropriate, and for each of the clusters, the lowest feature size was selected for the full data training. Additionally, feature sizes 1 to 5 were included so that the development of the profiles of the clusters could be demonstrated when more and more features were added to the clustering/prediction procedure.

#### 1.5. Sensitivity analyses

##### 1.5.1. Alternative feature selection

The purpose of the primary feature selection method, a stepwise feature addition based on univariate correlation, was to use a simple and robust method to investigate how genetic profiles evolve as increasing numbers of schizophrenia-relevant features are added into the models.
To address potential problems of this method, resulting mainly from unaddressed correlatedness of the selected features, we performed multivariate feature selection methods (see chapter 1.5.1.1 below) and a PCA based method (see chapter 1.5.3 below).

###### 1.5.1.1. LASSO and sparse-group LASSO

First, the lambda was tuned in 5-fold cross-validation using the function cv.glmnet(family = binomial, alpha = 1) from the R-package *glmnet[13]*. The lambda with the lowest out-of-fold deviance (lambda.min) was then used to train the model with the function glmnet(family = binomial, alpha = 1, lambda = lambda.min). This procedure was repeated 200 times in random 80% subsets of the data. The features whose effects were never regularized to 0 over the 200 random iterations were considered as the selected features. Because this approach might select against strong features that have a high correlation with another feature, we also performed feature selection using grouped sparse LASSO, where regularization is first applied to variable groups and only then within groups. The variable groups were formed so that all pairs of variables with an absolute correlation of 0.5 or higher belonged to the same group. Therefore, while not all variables have high correlations with all the other variables in the same group, all of them do have an absolute correlation of 0.5 or higher with at least one other variable in the group. The grouped sparse LASSO feature selection followed the same exact procedure with the same exact subsets of the data as the LASSO version. As *glmnet* does not include grouped sparse LASSO implementation, package *sparsegl[14]* was used with functions cv.sparsegl() and sparsegl() replacing cv.glmnet() and glmnet(), respectively, and the parameters remaining the same.
Applying these methods to our data resulted in unstable feature sets across cross-validation folds and repeated runs. This instability is likely due to the high degree of correlation among features and their relatively small individual effect sizes. As a consequence, biologically related traits were frequently

penalized and excluded by these methods, despite their potential relevance to SCZ, making downstream analyses and interpretations challenging.

##### 1.5.2. Alternative cluster assignment strategy

To address the possibility that the selected cluster assignment strategy introduced bias by leaving some patients unassigned, we implemented an alternative, forced-choice assignment strategy. First predicted probability was defined as the proportion of individual classification trees (51 forests × 1,000 trees = 51,000 trees total) in the random forest ensemble that classified a given individual as a case. Using these predicted probabilities, every patient was assigned to the cluster for which they had the highest value, regardless of the magnitude of that probability or its difference from the alternative. For example, a participant with a higher predicted probability of an SCZ diagnosis under the GRP1 model than under the GRP2 model was assigned to GRP1, irrespective of how close the two probabilities were. No ties occurred, so no patients required exclusion or arbitrary tie-breaking.

##### 1.5.3. Clustering based on the first principal components of each PGS category

We performed PCA within the GWAS groups we defined previously based on hierarchical clustering of their genetic correlation matrix (see Methods section *Formation of the PGS categories*). This was done at feature size 24, as that is the feature size for which we report the main results. The first PC from each GWAS group, alongside SCZ-PGS, was then used as a feature in the k-means clustering. As using this method on fold in a cross validation scheme when training the prediction models produced very unstable results, we used the full feature set when training the prediction models.

Applying this method to our data resulted in major challenges when trying to consequently use it for the full ML framework and overall incomplete leveraging of the available variance.

##### 1.5.4. Clusters Matched for SCZ-PGS

This analysis was conducted to determine whether the difference in predictive performance between the GRPs could be explained by differences in their SCZ-PGS levels. Though the SCZ-PGS means in patients assigned to GRP 1 and 2 were similar, they were not exactly the same. The slightly higher mean in GRP 1 could therefore explain the better case-control predictive performance. To see if that difference could indeed be driven by the SCZ-PGS difference alone, we performed the analyses with training patients groups matched for SCZ-PGS. This was achieved by matching a sample of equal size from the bigger cluster from the k-means part to the SCZ-PGS of the smaller cluster. Random subsamples of gradually decreasing size of the smaller cluster were taken until the matching could be achieved so that the difference in SCZ-PGS mean was no greater than 0.02 standard deviations. This was done for each feature size and performed utilizing the function matchit() from the R-package *MatchIt* [3]. After the patients groups matched for SCZ-PGS were derived, the analysis proceeded the same way as in the primary machine learning analyses.

#### 1.6. Research Ethics and Informed Consent

All participants provided written informed consent (see below). Procedures were approved by Ethics Committee II of the University of Heidelberg Medical Faculty Mannheim (protocol no. 2024-845) and the respective local ethical committees.

Study subjects in FinnGen provided informed consent for biobank research, based on the Finnish Biobank Act. Alternatively, separate research cohorts, collected prior the Finnish Biobank Act came into effect (in September 2013) and start of FinnGen (August 2017), were collected based on study-specific consents and later transferred to the Finnish biobanks after approval by Fimea (Finnish Medicines Agency), the National Supervisory Authority for Welfare and Health. Recruitment protocols followed the biobank protocols approved by Fimea. The Coordinating Ethics Committee of the Hospital District of Helsinki and Uusimaa (HUS) statement number for the FinnGen study is Nr HUS/990/2017.

The FinnGen study is approved by Finnish Institute for Health and Welfare (permit numbers: THL/2031/6.02.00/2017, THL/1101/5.05.00/2017, THL/341/6.02.00/2018, THL/2222/6.02.00/2018, THL/283/6.02.00/2019, THL/1721/5.05.00/2019 and THL/1524/5.05.00/2020), Digital and population data service agency (permit numbers: VRK43431/2017-3, VRK/6909/2018-3, VRK/4415/2019-3), the Social Insurance Institution (permit numbers: KELA 58/522/2017, KELA 131/522/2018, KELA 70/522/2019, KELA 98/522/2019, KELA 134/522/2019, KELA 138/522/2019, KELA 2/522/2020, KELA 16/522/2020), Findata permit numbers THL/2364/14.02/2020, THL/4055/14.06.00/2020, THL/3433/14.06.00/2020, THL/4432/14.06/2020, THL/5189/14.06/2020, THL/5894/14.06.00/2020, THL/6619/14.06.00/2020, THL/209/14.06.00/2021, THL/688/14.06.00/2021, THL/1284/14.06.00/2021, THL/1965/14.06.00/2021, THL/5546/14.02.00/2020, THL/2658/14.06.00/2021, THL/4235/14.06.00/2021, Statistics Finland (permit numbers: TK-53-1041-17 and TK/143/07.03.00/2020 (earlier TK-53-90-20) TK/1735/07.03.00/2021, TK/3112/07.03.00/2021) and Finnish Registry for Kidney Diseases permission/extract from the meeting minutes on 4th July 2019.

The Biobank Access Decisions for FinnGen samples and data utilized in FinnGen Data Freeze 12 include: THL Biobank BB2017_55, BB2017_111, BB2018_19, BB_2018_34, BB_2018_67, BB2018_71, BB2019_7, BB2019_8, BB2019_26, BB2020_1, BB2021_65, Finnish Red Cross Blood Service Biobank

7.12.2017, Helsinki Biobank HUS/359/2017, HUS/248/2020, HUS/430/2021 §28, §29, HUS/150/2022 §12, §13, §14, §15, §16, §17, §18, §23, §58, §59, HUS/128/2023 §18, Auria Biobank AB17-5154 and amendment #1 (August 17 2020) and amendments BB_2021-0140, BB_2021-0156 (August 26 2021, Feb 2 2022), BB_2021-0169, BB_2021-0179, BB_2021-0161, AB20-5926 and amendment #1 (April 23 2020) and it´s modifications (Sep 22 2021), BB_2022-0262, BB_2022-0256, Biobank Borealis of Northern Finland_2017_1013, 2021_5010, 2021_5010 Amendment, 2021_5018, 2021_5018 Amendment, 2021_5015, 2021_5015 Amendment, 2021_5015 Amendment_2, 2021_5023, 2021_5023 Amendment, 2021_5023 Amendment_2, 2021_5017, 2021_5017 Amendment, 2022_6001, 2022_6001 Amendment, 2022_6006 Amendment, 2022_6006 Amendment, 2022_6006 Amendment_2, BB22-0067, 2022_0262, 2022_0262 Amendment, Biobank of Eastern Finland 1186/2018 and amendment 22§/2020, 53§/2021, 13§/2022, 14§/2022, 15§/2022, 27§/2022, 28§/2022, 29§/2022, 33§/2022, 35§/2022, 36§/2022, 37§/2022, 39§/2022, 7§/2023, 32§/2023, 33§/2023, 34§/2023, 35§/2023, 36§/2023, 37§/2023, 38§/2023, 39§/2023, 40§/2023, 41§/2023, Finnish Clinical Biobank Tampere MH0004 and amendments (21.02.2020 & 06.10.2020), BB2021-0140 8§/2021, 9§/2021, §9/2022, §10/2022, §12/2022, 13§/2022, §20/2022, §21/2022, §22/2022, §23/2022, 28§/2022, 29§/2022, 30§/2022, 31§/2022, 32§/2022, 38§/2022, 40§/2022, 42§/2022, 1§/2023, Central Finland Biobank 1-2017, BB_2021-0161, BB_2021-0169, BB_2021-0179, BB_2021-0170, BB_2022-0256, BB_2022-0262, BB22-0067, Decision allowing to continue data processing until 31st Aug 2024 for projects: BB_2021-0179, BB22-0067,BB_2022-0262, BB_2021-0170, BB_2021-0164, BB_2021-0161, and BB_2021-0169, and Terveystalo Biobank STB 2018001 and amendment 25th Aug 2020, Finnish Hematological Registry and Clinical Biobank decision 18th June 2021, Arctic biobank P0844: ARC_2021_1001.

#### 1.7. Funding

This work was supported by the Hector foundation II, the German Federal Ministry of Education and Research (BEST project, grant 01EK2101B), the German Research Foundation (DFG) (TRR 265/2 TP A06), and was endorsed by German Center for Mental Health (DZPG). U.H. was supported by European Union’s Horizon 2020 Research and Innovation Programme (PSY-PGx, grant agreement No 945151) and the Deutsche Forschungsgemeinschaft (DFG, German Research Foundation, project number 514201724). T.G.S. was supported by the European Union Horizon 2020 Research and Innovation

Program (PSY-PGx, grant agreement No 945151), and also by the Deutsche Forschungsgemeinschaft within the framework of the projects www.kfo241.de and www.PsyCourse.de [SCHU 1603/4–1, 5–1, 7–1]. T.G.S. was further supported by the Dr Lisa Oehler Foundation (Kassel, Germany), the Bundesministerium für Bildung und Forschung (BMBF, Federal Ministry of Education and Research; projects: IntegraMent [01ZX1614K], BipoLife [01EE1404H], e:Med Program [01ZX1614K]) and European Union’s Horizon 2020 Research and Innovation Programme (ERA-NET Neuron Projects GEPI-BIOPSY [BMBF No 01EW2005] and MulioBio [BMBF No 01EW2009]). G.C.K. 's PhD scholarship is financially supported by Exprivia S.p.A. under the Italian ministerial decree D.M. 351. Furthermore, this study was supported by FAIR - Future AI Research (PE00000013), spoke 6 – Symbiotic AI, under the NRRP MUR program funded by the NextGenerationEU, awarded to L.A.A. and G.P.; Project CUP : H97G22000210007, and the Apulian regional government for the project: Early Identification of Psychosis Risk, awarded to A.B.

#### Supplementary Figure 1. Automated GWAS search module workflow

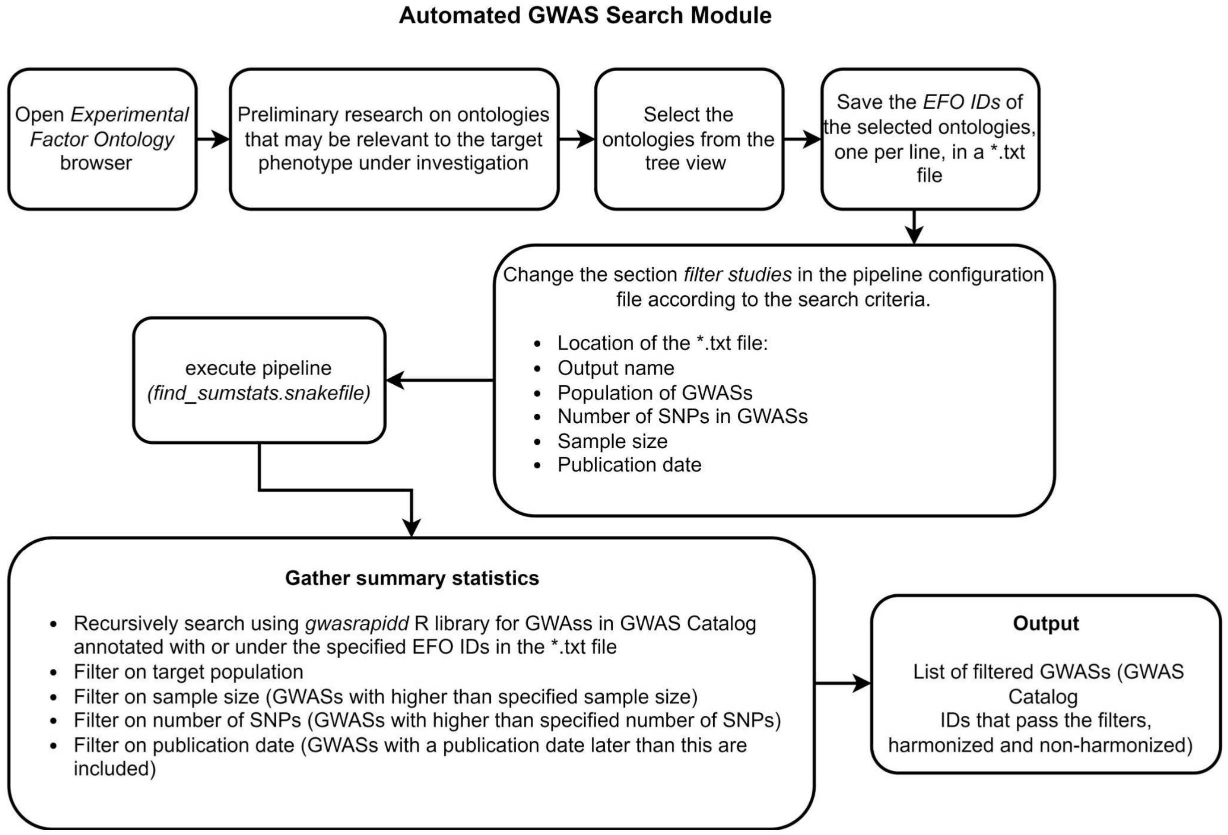

**Supplementary Figure 2. Hierarchical clustering of GWAS based on genetic correlations at feature size 45**

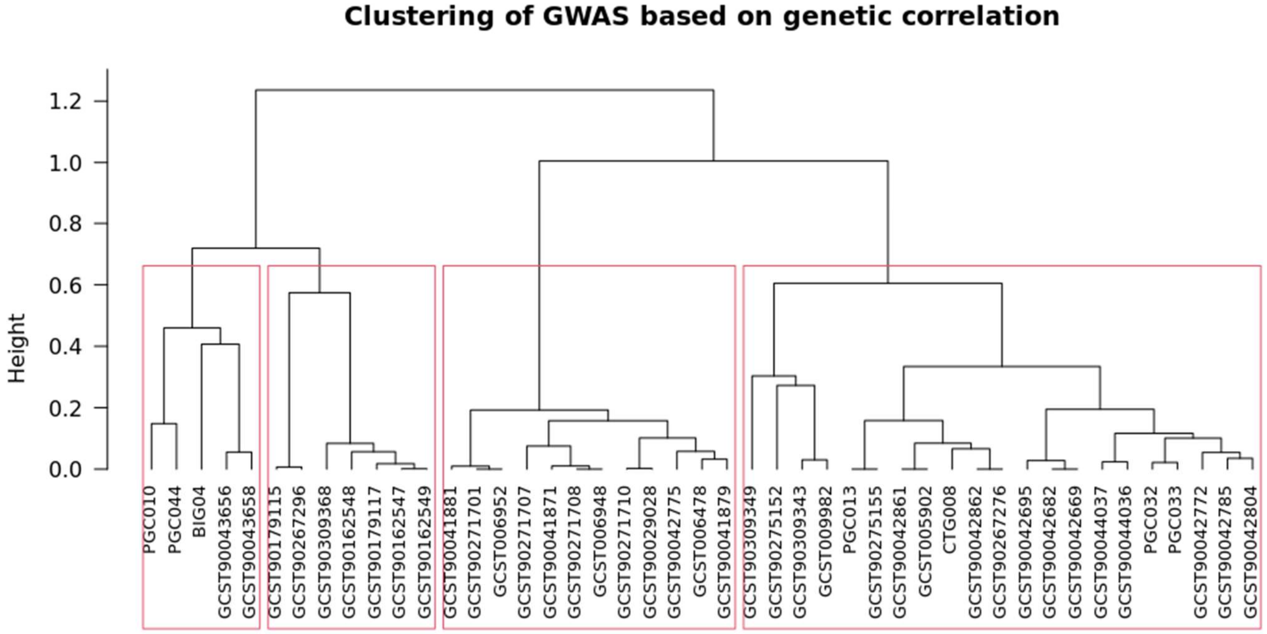


Red boxes indicate the four identified GWAS categories. From left to right, the clusters represent schizophrenia-related, cognition-related, neuroticism-related, and depression-related GWAS. The group first on the left contains SCZ itself, a gene-environment interaction GWAS and two measures related to alcohol use. For clearer interpretation of selected downstream analyses characterizing the contribution of the different PGS categories, we investigated SCZ as a single PGS.

**Supplementary Figure 3. Genetic correlations between the top 45 source GWAS**

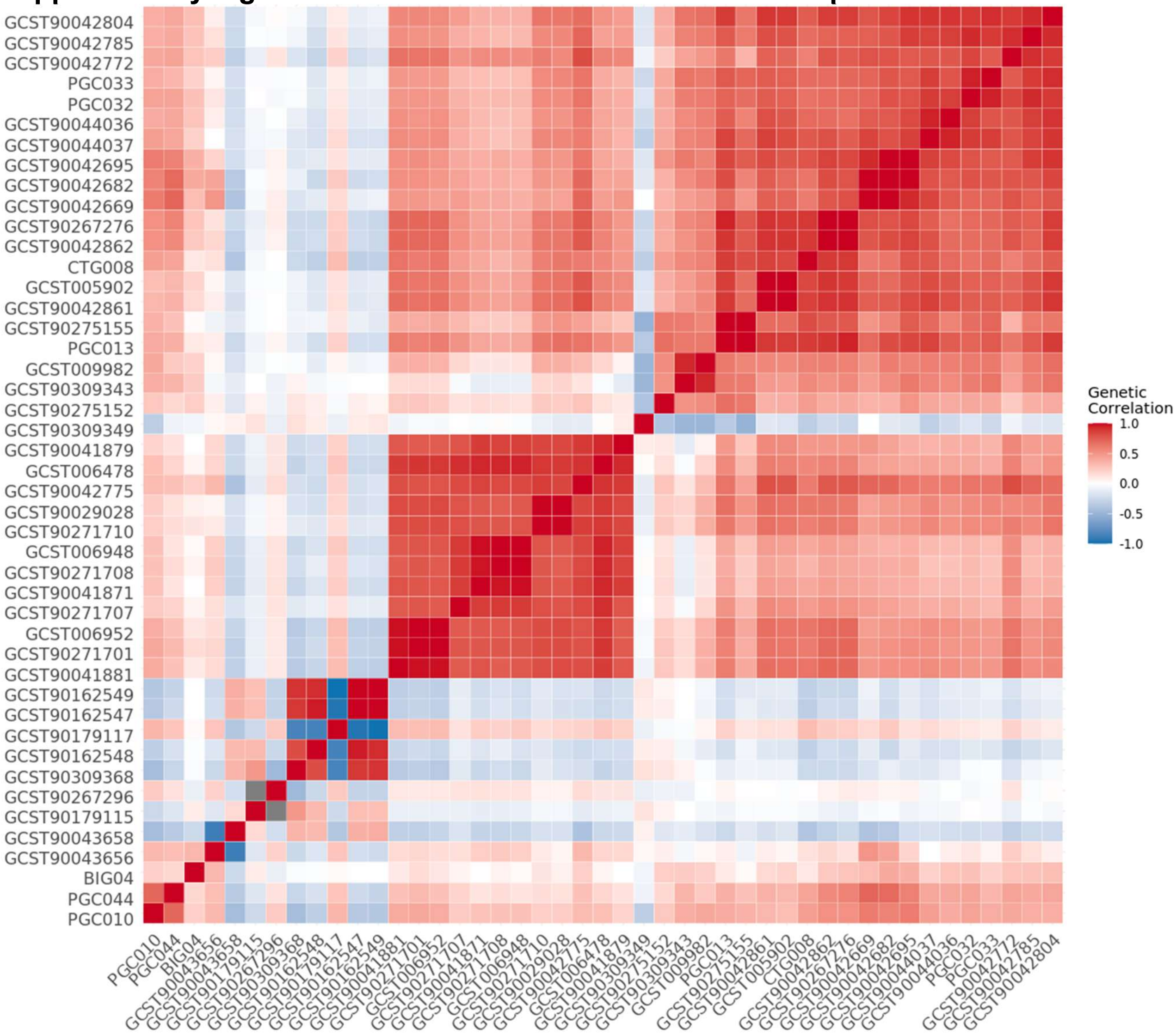


The GWAS on the x- and y-axes were ranked according to their relevance to SCZ.

**Supplementary Figure 4. Hierarchical clustering of GWAS based on genetic correlations at feature size 12**

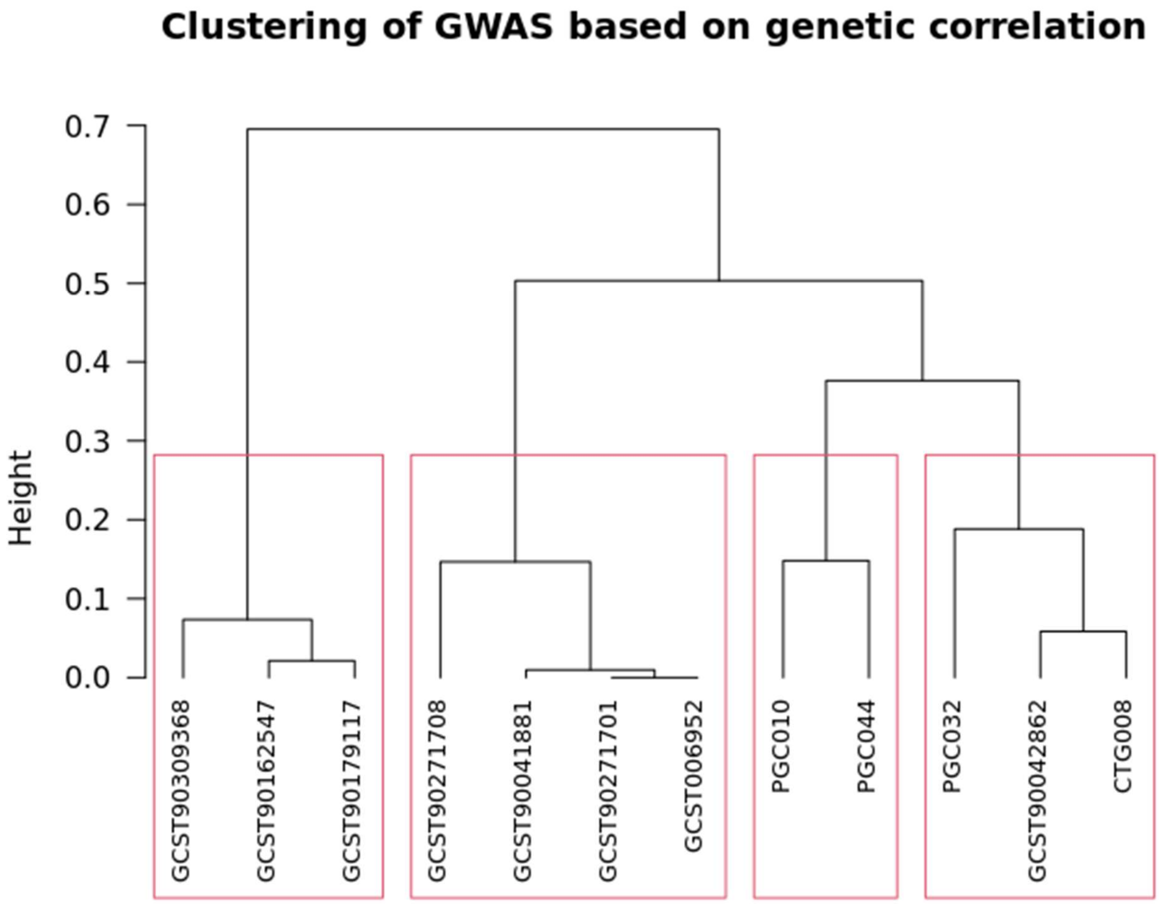


Red boxes indicate the four identified GWAS categories. From left to right, the clusters represent cognition-related, neuroticism-related, schizophrenia-related, and depression-related GWAS.

**Supplementary Figure 5. Hierarchical clustering of GWAS based on genetic correlations at feature size 16**

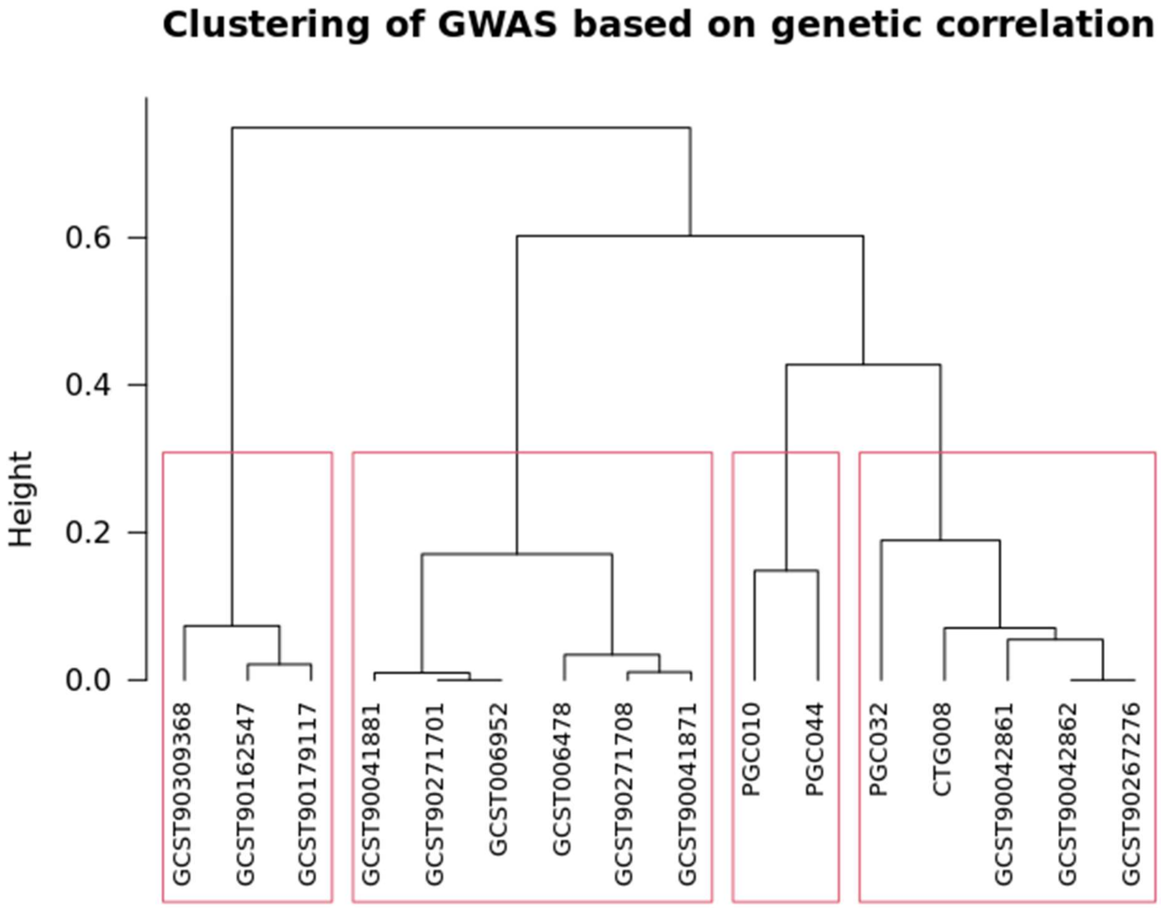


Red boxes indicate the four identified GWAS categories. From left to right, the clusters represent cognition-related, neuroticism-related, schizophrenia-related, and depression-related GWAS.

**Supplementary Figure 6. Genetic correlations between the top 12 source GWAS**

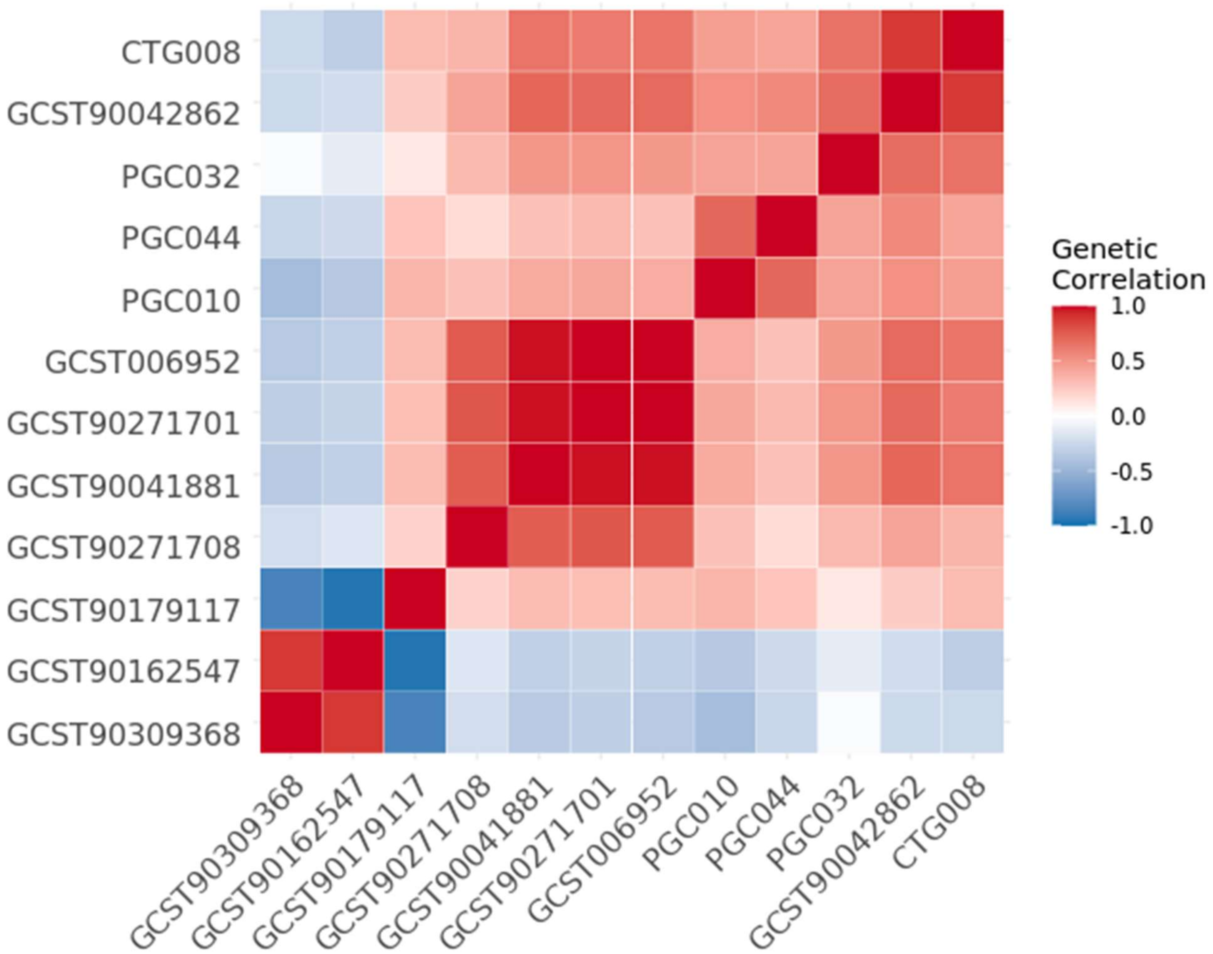


From left to right, the clusters represent cognition-related, neuroticism-related, schizophrenia-related, and depression-related GWAS.

**Supplementary Figure 7. Genetic correlations between the top 16 source GWAS**

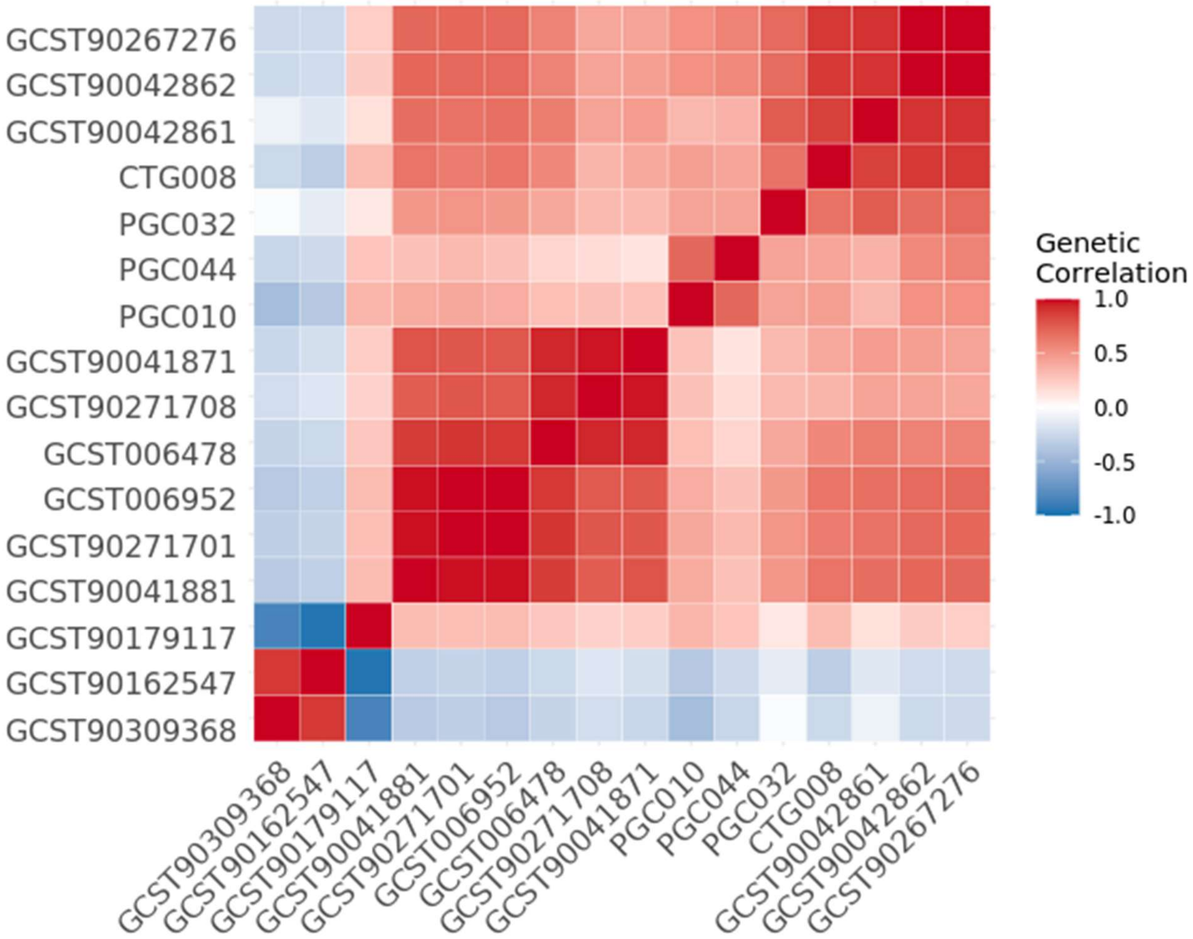


From left to right, the clusters represent cognition-related, neuroticism-related, schizophrenia-related, and depression-related GWAS.

**Supplementary Figure 8. Determination of the optimal number of clusters using the first-difference criterion**

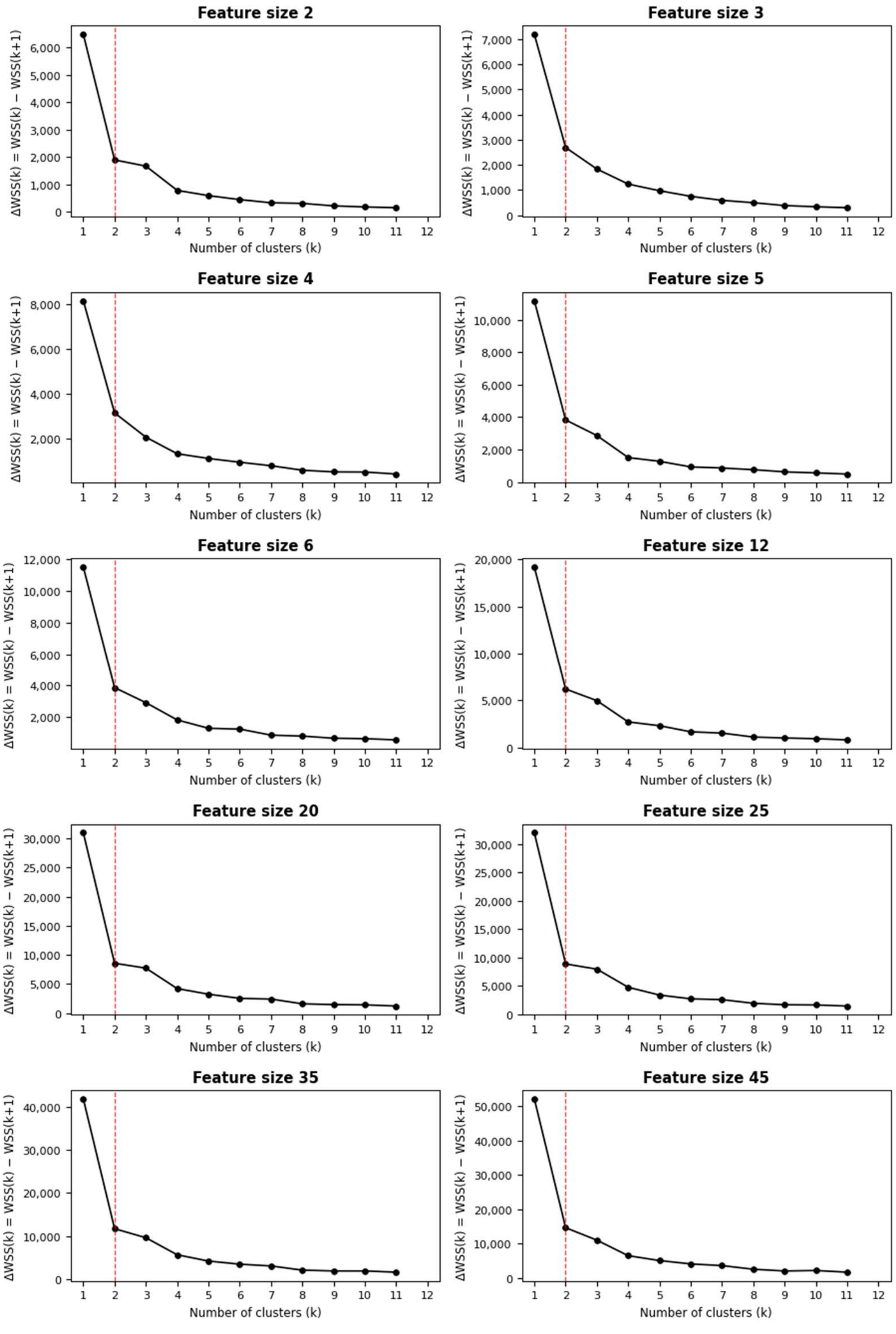


First-difference criterion based on the reduction in within-cluster sum of squares (ΔWSS) for determining the number of clusters. ΔWSS represents the improvement in within-cluster homogeneity obtained by increasing the number of clusters from (k-1) to (k). Across all evaluated feature sizes, the largest reduction in WSS occurred when increasing the number of clusters from one to two, indicating that the primary partition of the data is captured by a two-cluster solution. WSS values were evaluated for (k=1)–12.

**Supplementary Figure 9. Projection of LASSO-derived GRPs onto univariately selected features at feature size 36**

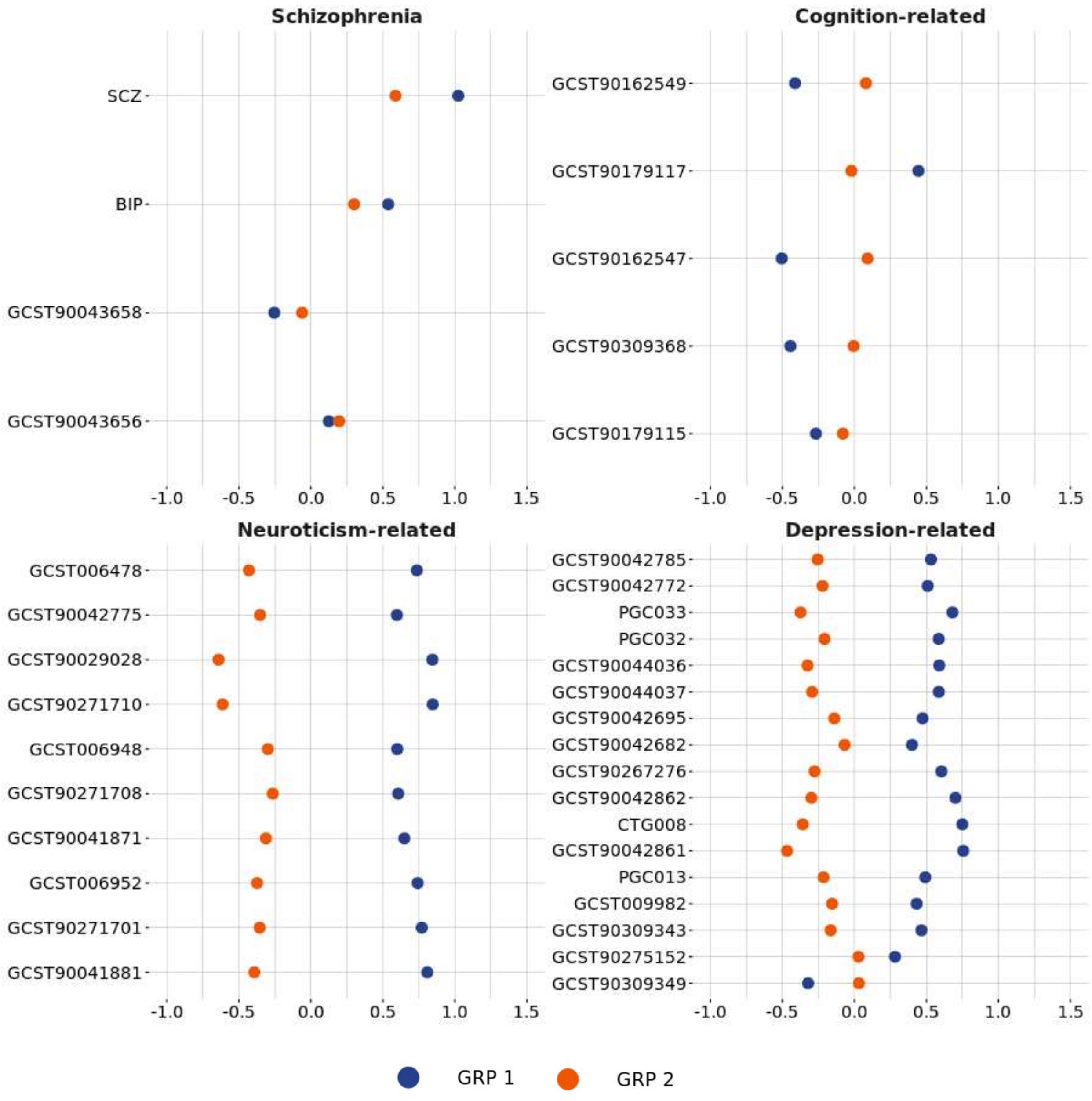


Points represent the mean standardized value of each polygenic risk score among individuals assigned to GRP 1 (blue) or GRP 2 (orange). The scores were grouped into schizophrenia-, cognition-, neuroticism-, and depression-related categories.

**Supplementary Figure 10. Projection of sparse-group LASSO-derived GRPs onto univariately selected features at feature size 41**

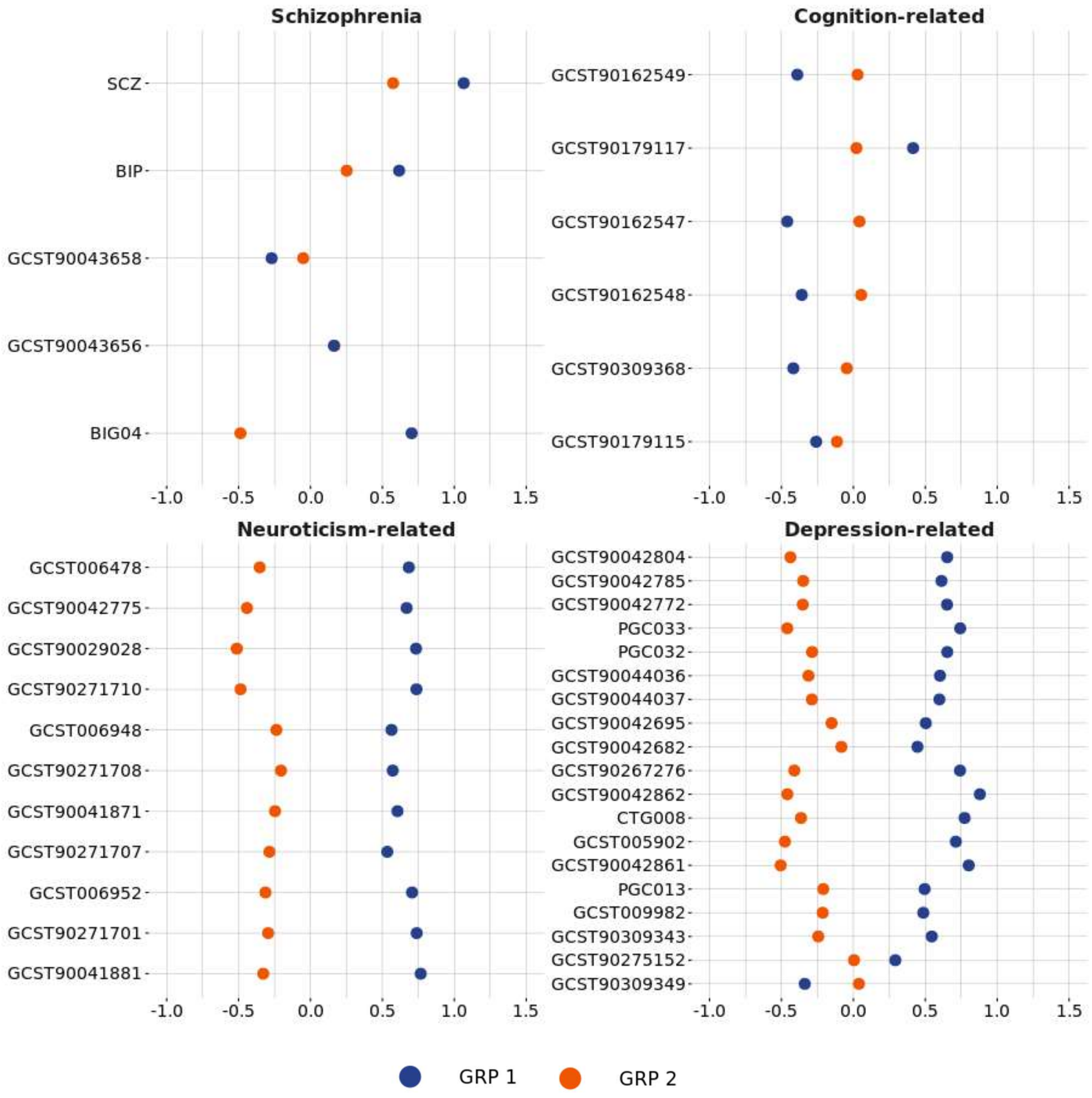


Points represent the mean standardized value of each polygenic risk score among individuals assigned to GRP 1 (blue) or GRP 2 (orange). The scores were grouped into schizophrenia-, cognition-, neuroticism-, and depression-related categories.

**Supplementary Figure 11. GRPs uniquely predicted by models trained on clusters grouped based on the first principal component of each PGS category at feature size 24**

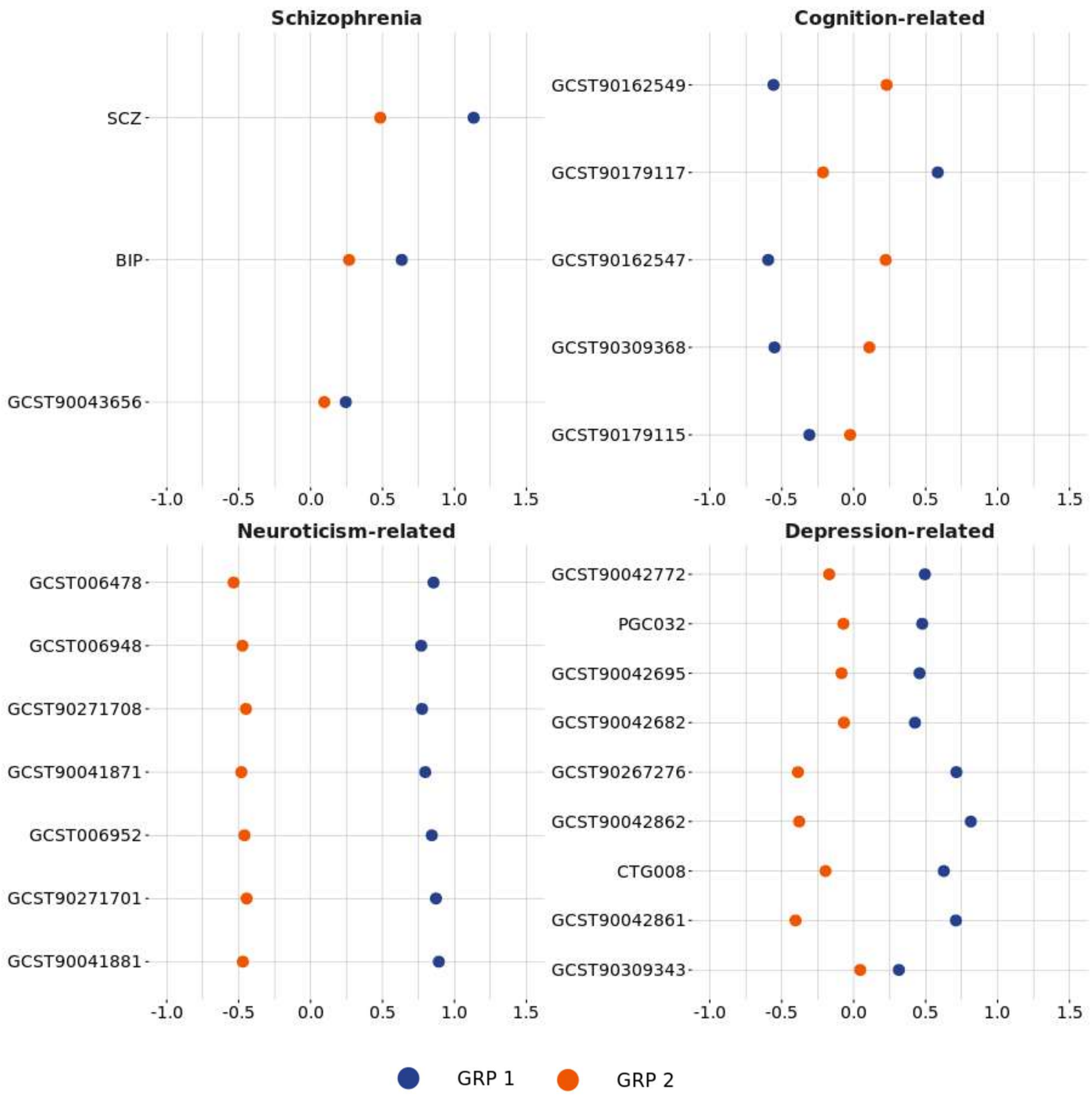


Points represent the mean standardized value of each polygenic risk score among individuals assigned to GRP 1 (blue) or GRP 2 (orange). The scores were grouped into schizophrenia-, cognition-, neuroticism-, and depression-related categories.

**Supplementary Figure 12. The genetic profiles of the composite PGS groups across feature sizes**

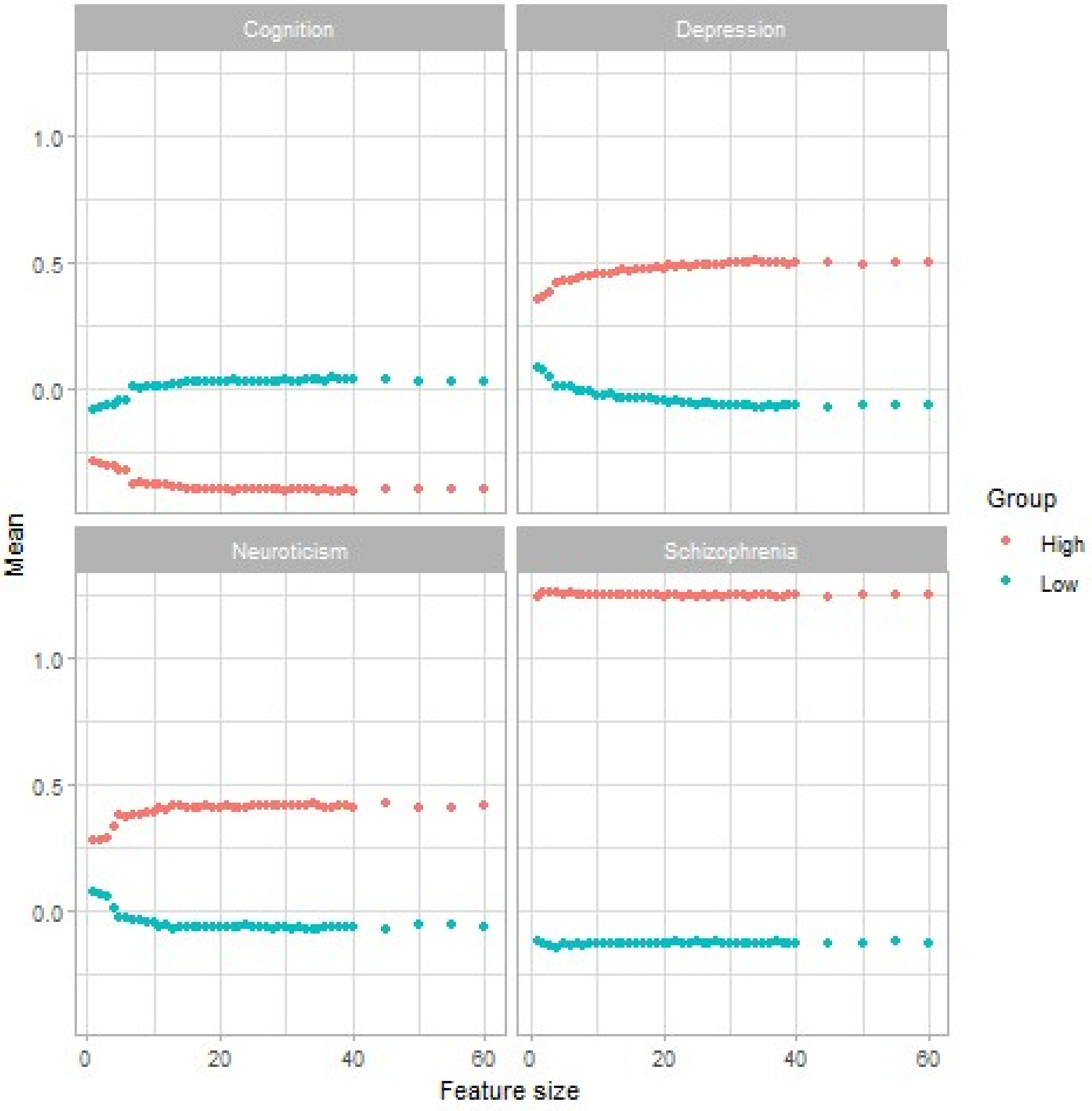


The values on the y-axis indicate the means of the first PC of PGS belonging to the cognition, depression and neuroticism GWAS groups. Color marks the composite PGS group, high being the high genetic risk group.

**Supplementary Figure 13. Comparison of predictive performance between the multi-PGS and SCZ-PGS only models**

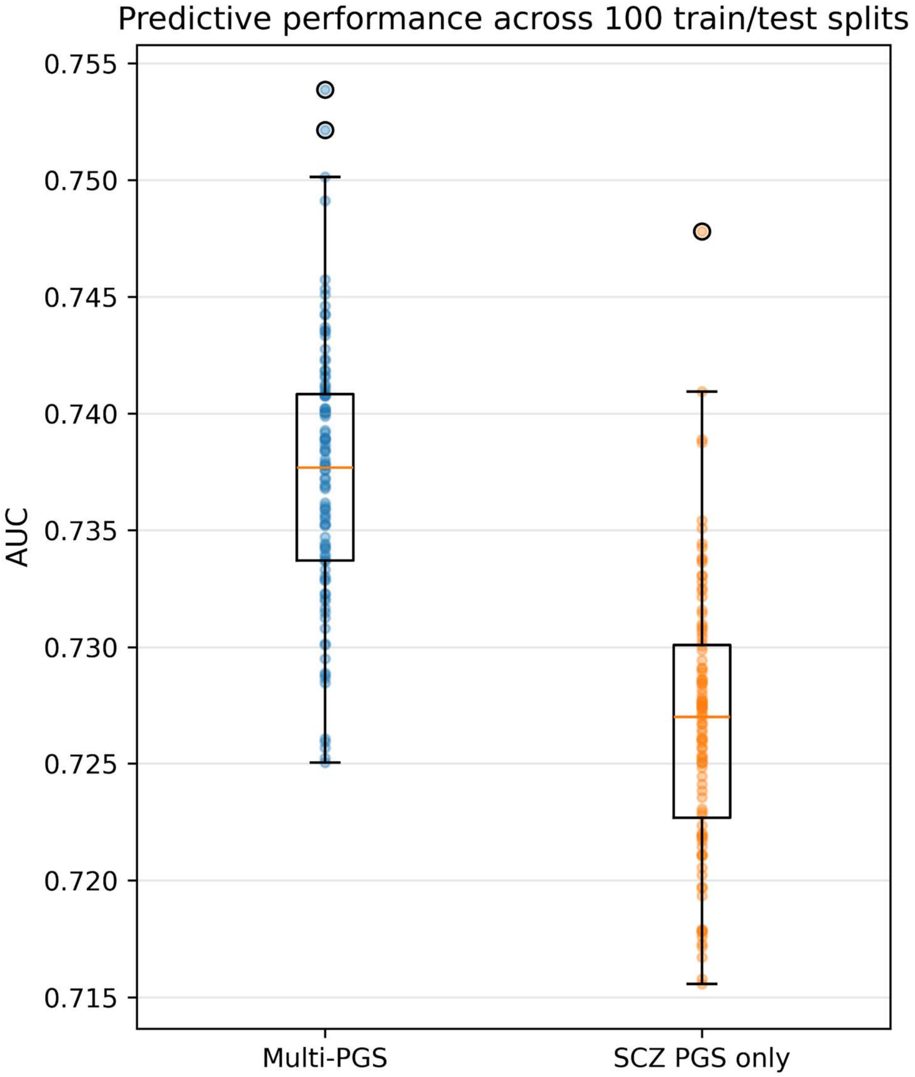

**Supplementary Figure 14. GRPs uniquely predicted by models trained on clusters grouped based on matched for SCZ-PGS at feature size 45**

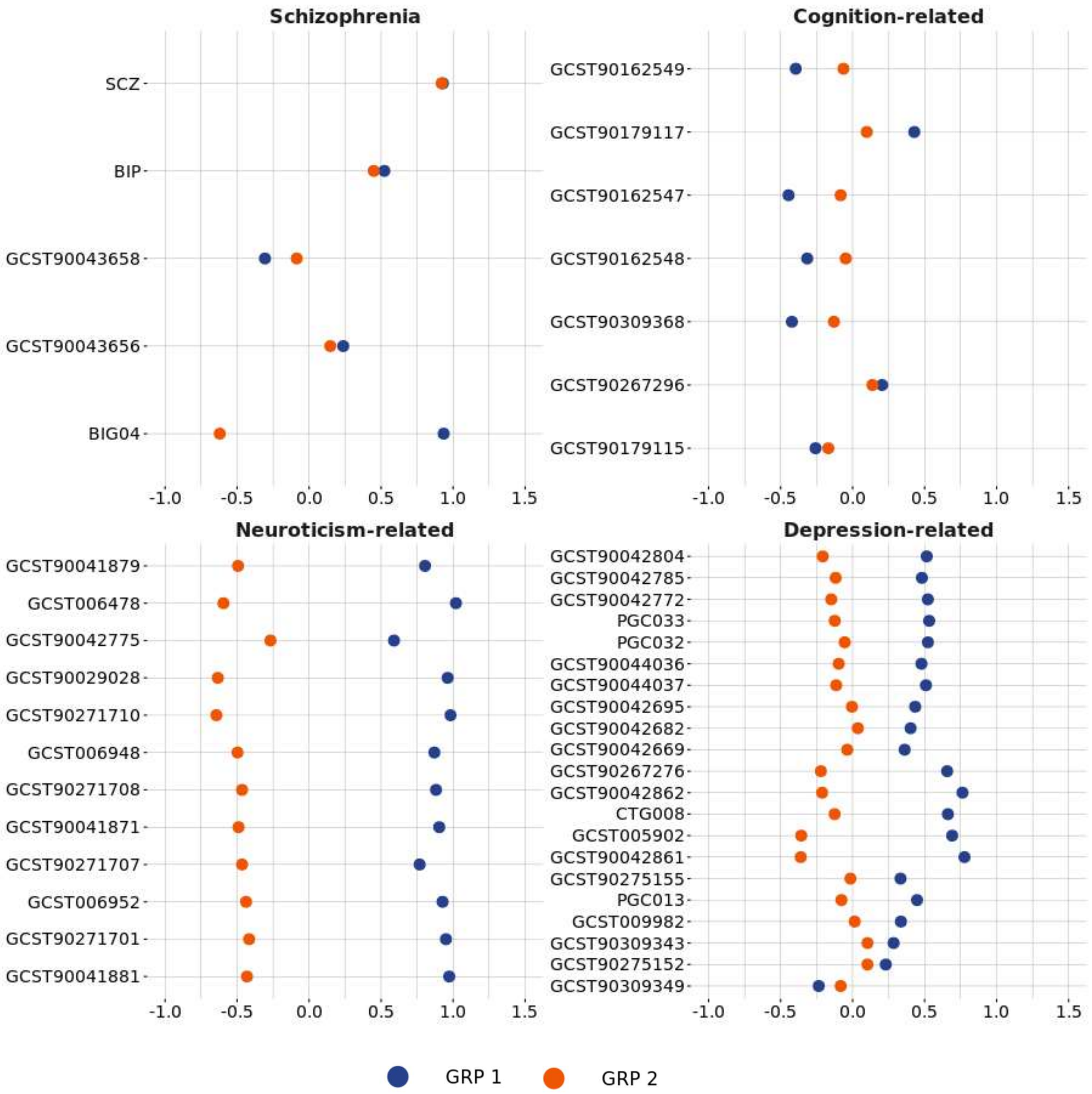


Points represent the mean standardized value of each polygenic risk score among individuals assigned to GRP 1 (blue) or GRP 2 (orange). The scores were grouped into schizophrenia-, cognition-, neuroticism-, and depression-related categories.

**Supplementary Figure 15. Predictive performance of models trained on the clusters grouped based on the matched for SCZ-PGS**

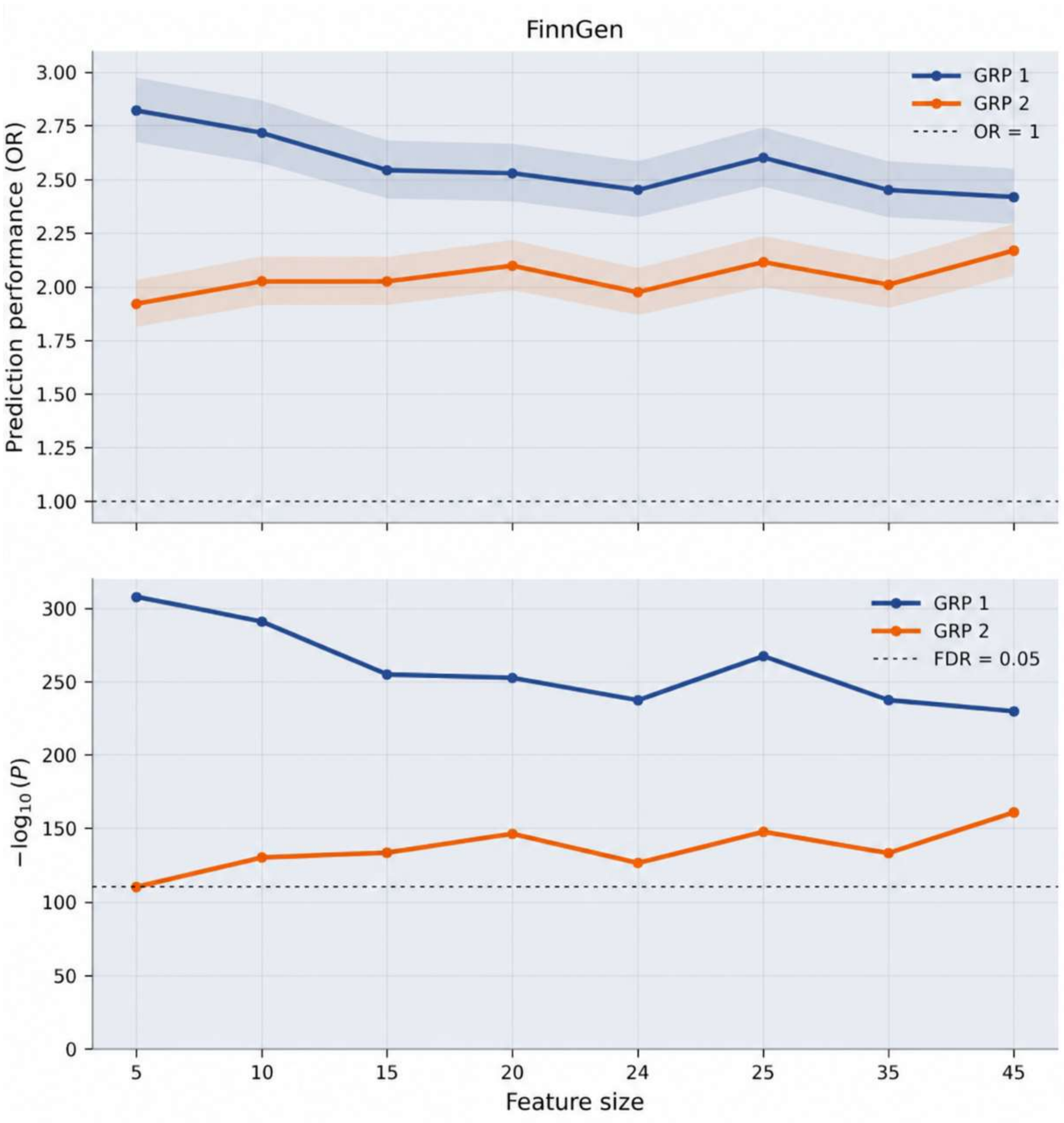

**Supplementary Figure 16. Feature size 1: Differences in comorbidity rates of mental disorders and disorder categories between the GRPs in $\text{Train}_{\text{FinnGen}}$.**

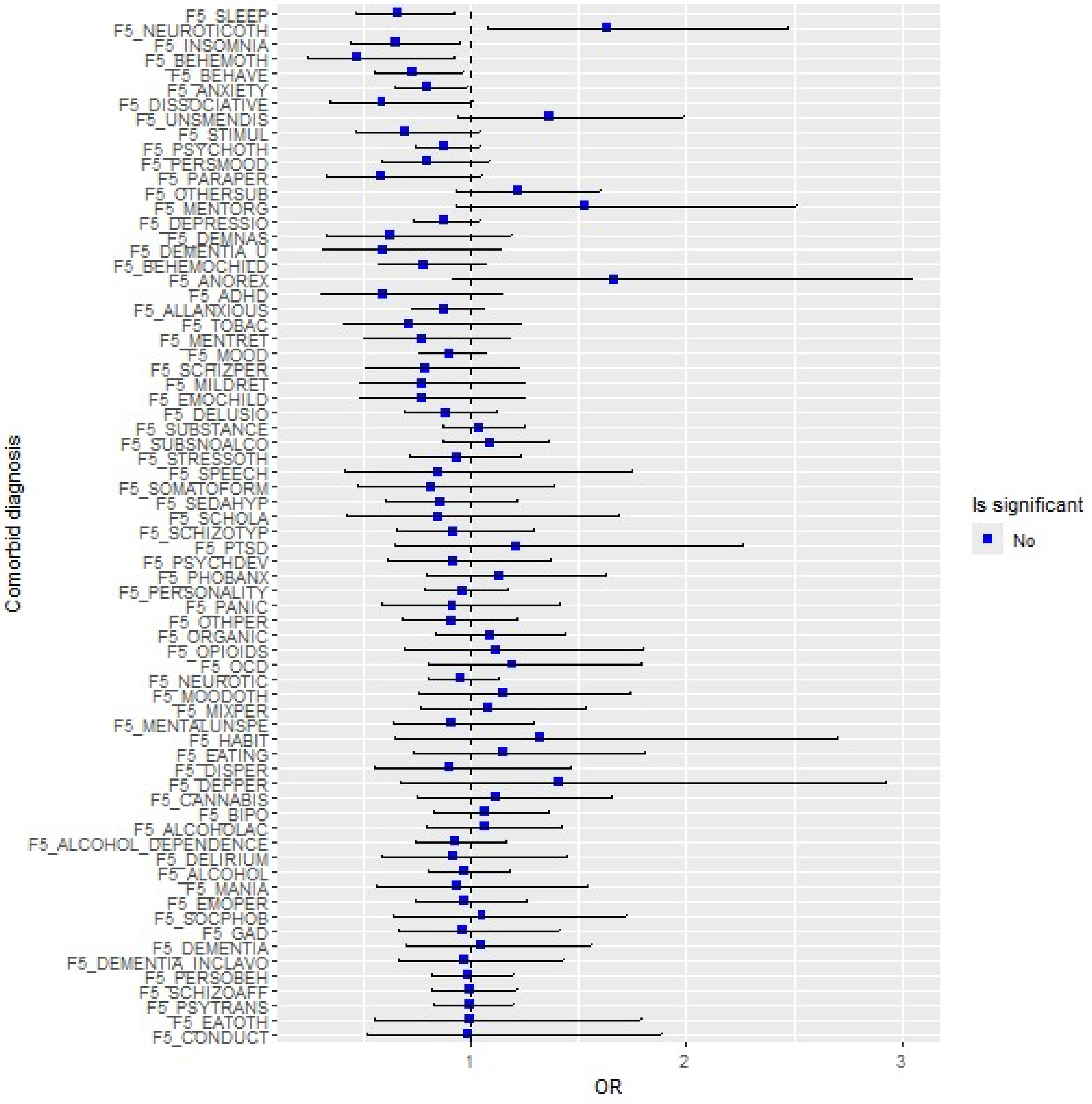


Red color indicates statistical significance at FDR of 0.05. The error bars show 95 % confidence intervals. GRP 2 was used as the reference group; therefore, ORs < 1 indicate higher rates in GRP 2, whereas ORs > 1 indicate higher rates in GRP 1. The definitions of the phenotype codes for comorbid diagnosis can be found in Supplementary Table 32.

**Supplementary Figure 17. Feature size 2: Differences in comorbidity rates of mental disorders and disorder categories between the GRPs in Train$_{FinnGen}$.**

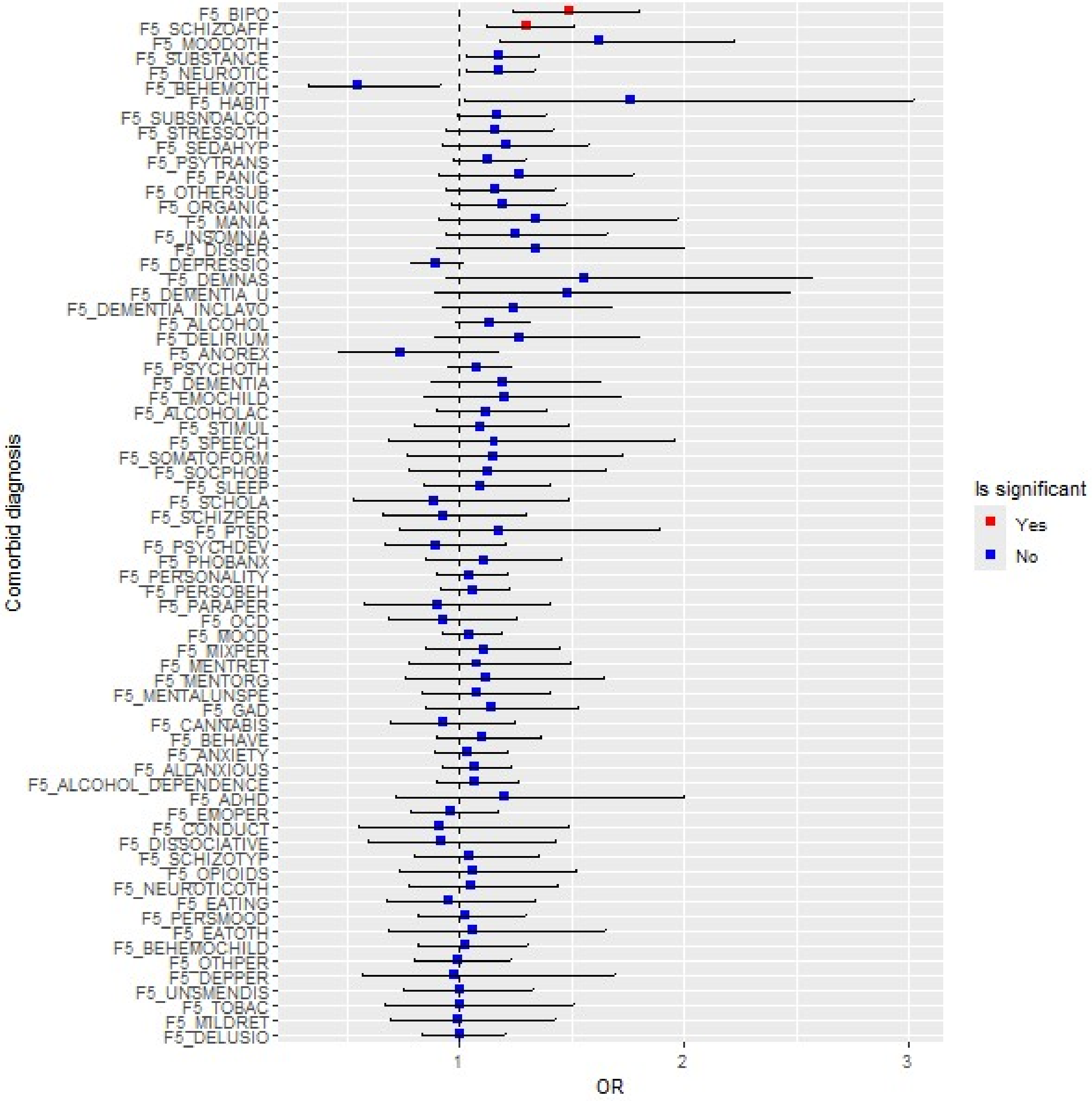


Red color indicates statistical significance at FDR of 0.05. The error bars show 95 % confidence intervals. GRP 2 was used as the reference group; therefore, ORs < 1 indicate higher rates in GRP 2, whereas ORs > 1 indicate higher rates in GRP 1. The definitions of the phenotype codes for comorbid diagnosis can be found in Supplementary Table 32.

**Supplementary Figure 18. Feature size 3: Differences in comorbidity rates of mental disorders and disorder categories between the GRPs in Train$_{FinnGen}$.**

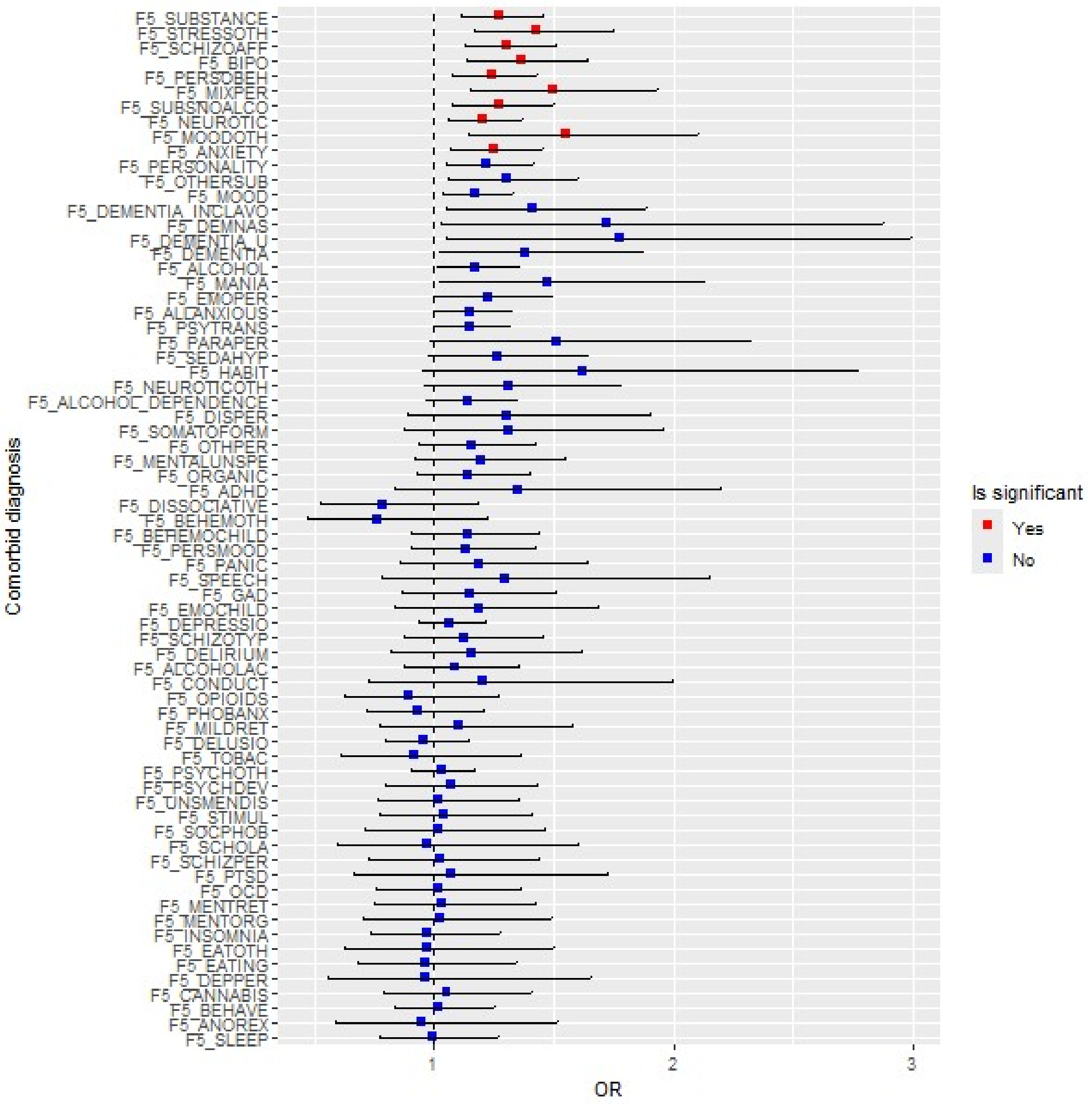


Red color indicates statistical significance at FDR of 0.05. The error bars show 95 % confidence intervals. GRP 2 was used as the reference group; therefore, ORs < 1 indicate higher rates in GRP 2, whereas ORs > 1 indicate higher rates in GRP 1. The definitions of the phenotype codes for comorbid diagnosis can be found in Supplementary Table 32.

**Supplementary Figure 19. Feature size 4: Differences in comorbidity rates of mental disorders and disorder categories between the GRPs in $\text{Train}_{\text{FinnGen}}$.**

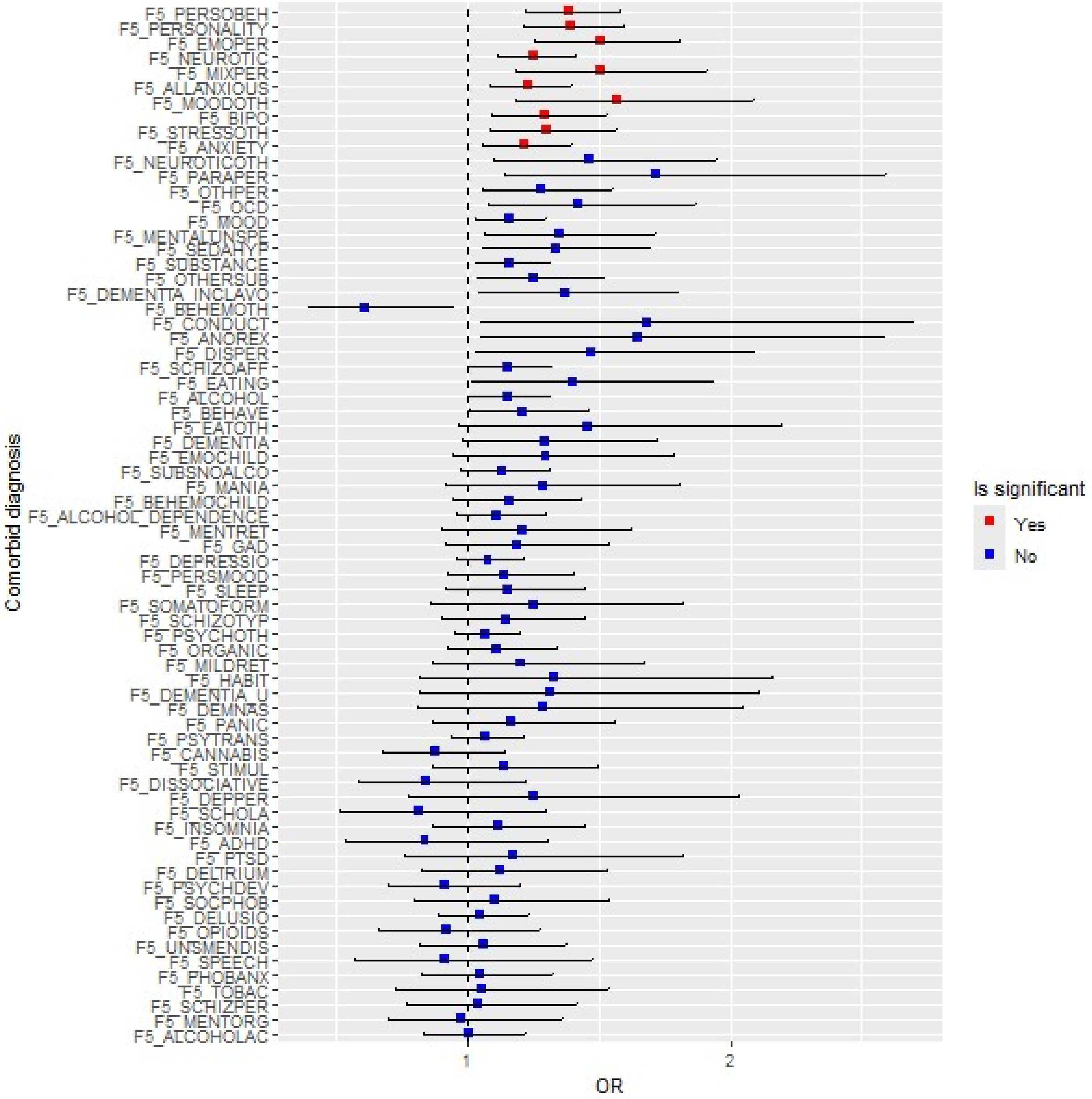


Red color indicates statistical significance at FDR of 0.05. The error bars show 95 % confidence intervals. GRP 2 was used as the reference group; therefore, ORs < 1 indicate higher rates in GRP 2, whereas ORs > 1 indicate higher rates in GRP 1. The definitions of the phenotype codes for comorbid diagnosis can be found in Supplementary Table 32.

**Supplementary Figure 20. Feature size 5: Differences in comorbidity rates of mental disorders and disorder categories between the GRPs in $Train_{FinnGen}$.**

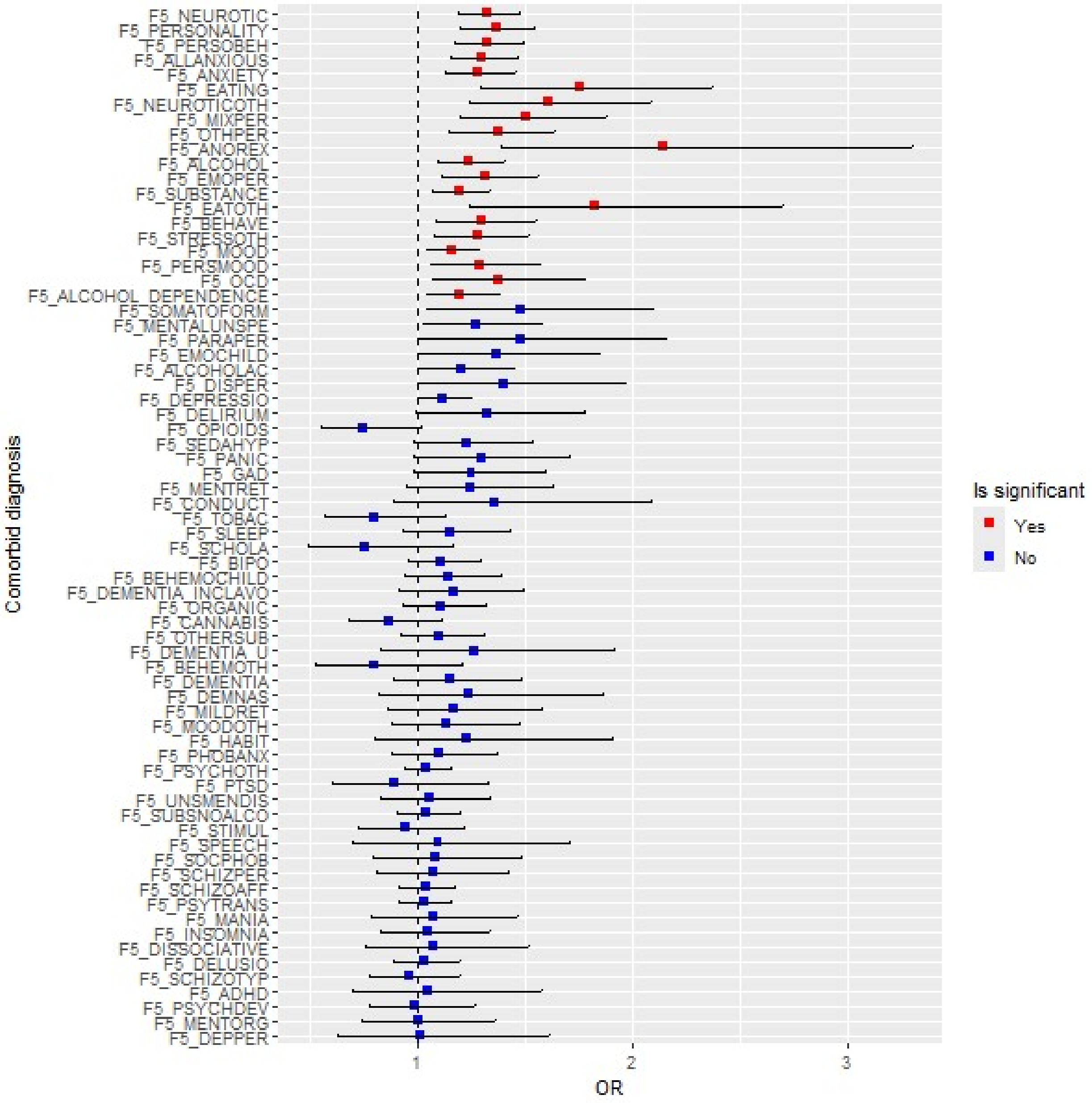


Red color indicates statistical significance at FDR of 0.05. The error bars show 95 % confidence intervals. GRP 2 was used as the reference group; therefore, ORs < 1 indicate higher rates in GRP 2, whereas ORs > 1 indicate higher rates in GRP 1. The definitions of the phenotype codes for comorbid diagnosis can be found in Supplementary Table 32.

**Supplementary Figure 21. Feature size 6: Differences in comorbidity rates of mental disorders and disorder categories between the GRPs in $Train_{FinnGen}$.**

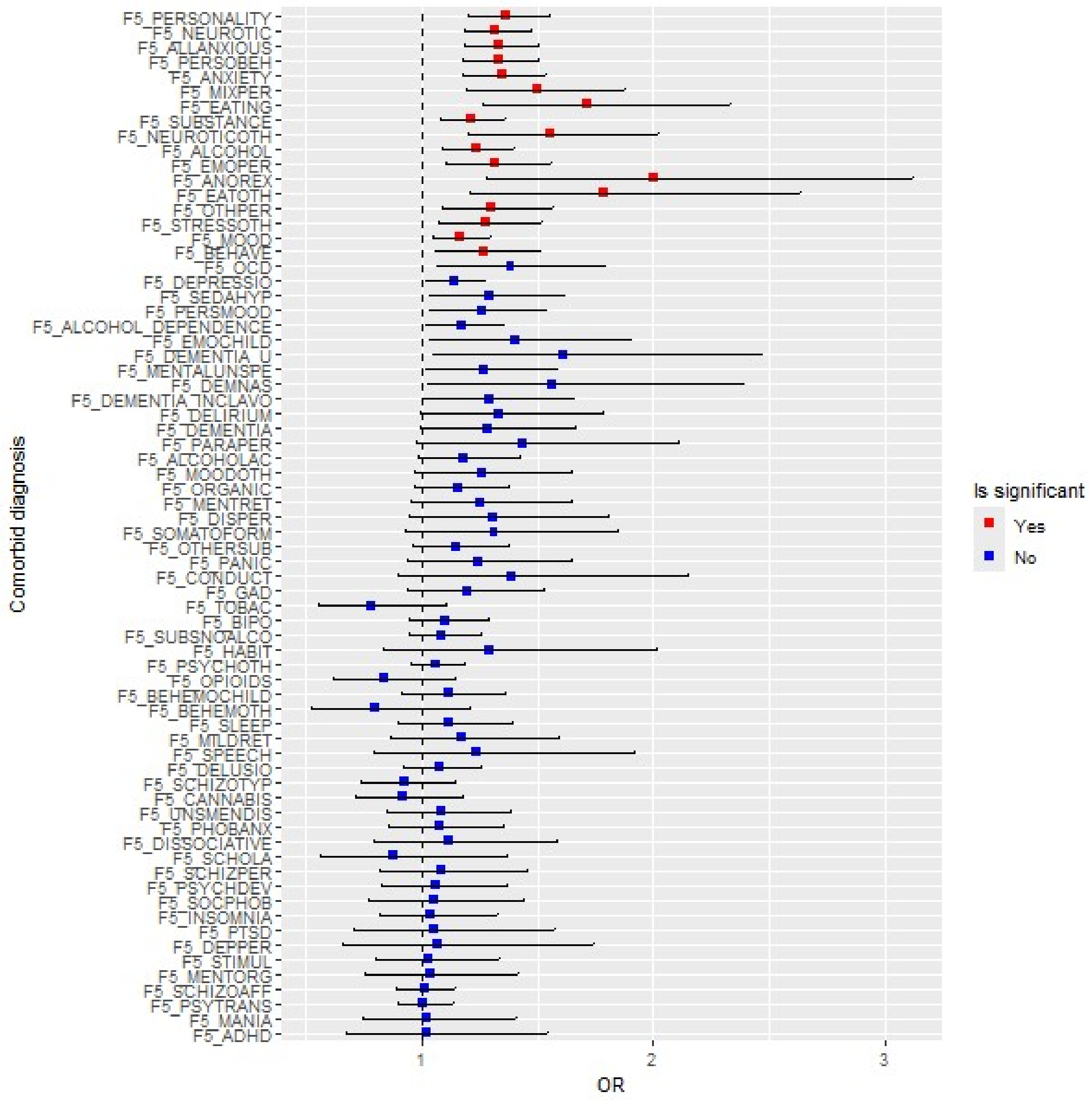


Red color indicates statistical significance at FDR of 0.05. The error bars show 95 % confidence intervals. GRP 2 was used as the reference group; therefore, ORs < 1 indicate higher rates in GRP 2, whereas ORs > 1 indicate higher rates in GRP 1. The definitions of the phenotype codes for comorbid diagnosis can be found in Supplementary Table 32.

**Supplementary Figure 22. Feature size 12: Differences in comorbidity rates of mental disorders and disorder categories between the GRPs in Train$_{\text{FinnGen}}$.**

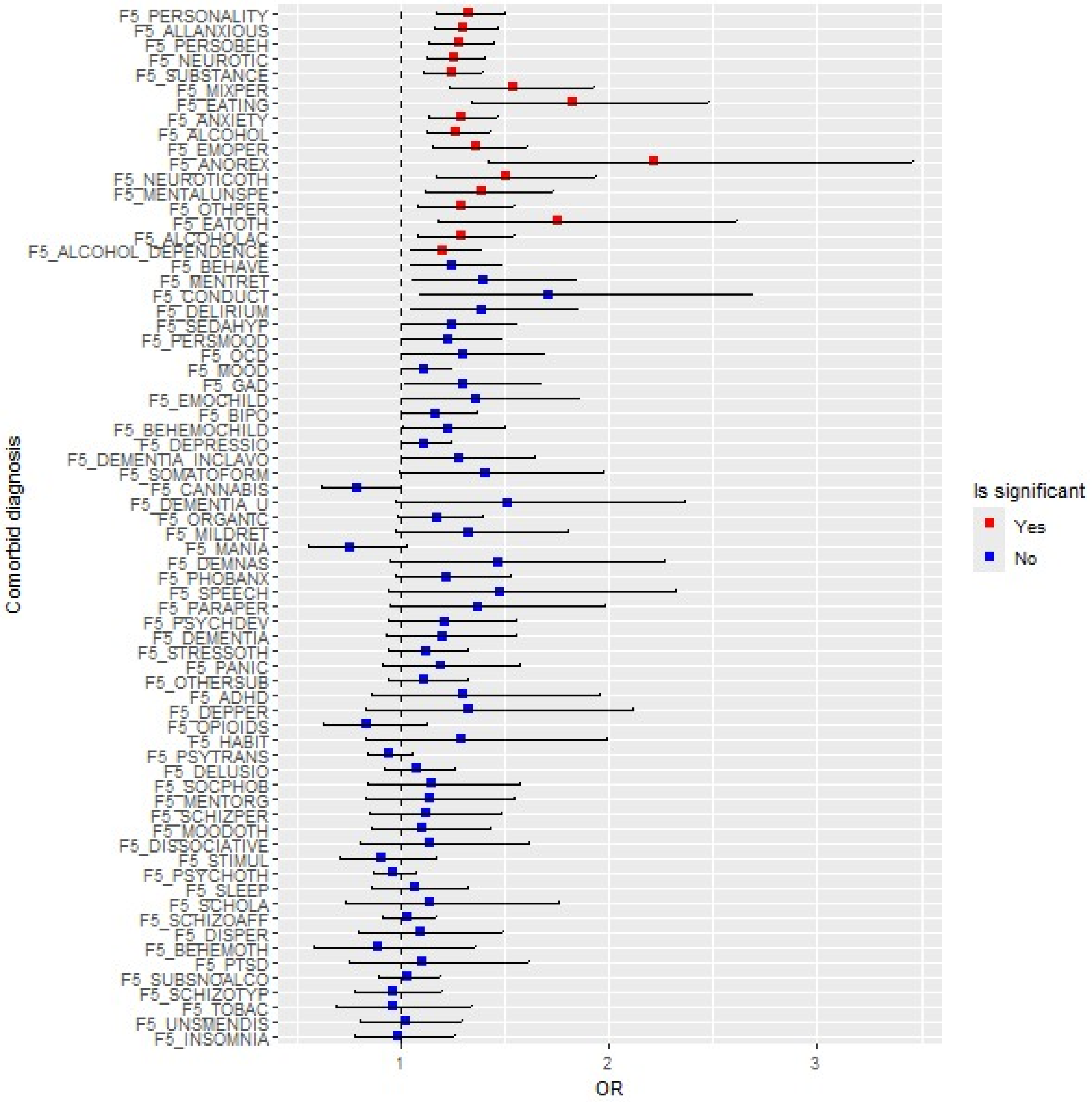


Red color indicates statistical significance at FDR of 0.05. The error bars show 95 % confidence intervals. GRP 2 was used as the reference group; therefore, ORs < 1 indicate higher rates in GRP 2, whereas ORs > 1 indicate higher rates in GRP 1. The definitions of the phenotype codes for comorbid diagnosis can be found in Supplementary Table 32.

**Supplementary Figure 23. Feature size 24: Differences in comorbidity rates of mental disorders and disorder categories between the GRPs in Train$_{FinnGen}$.**

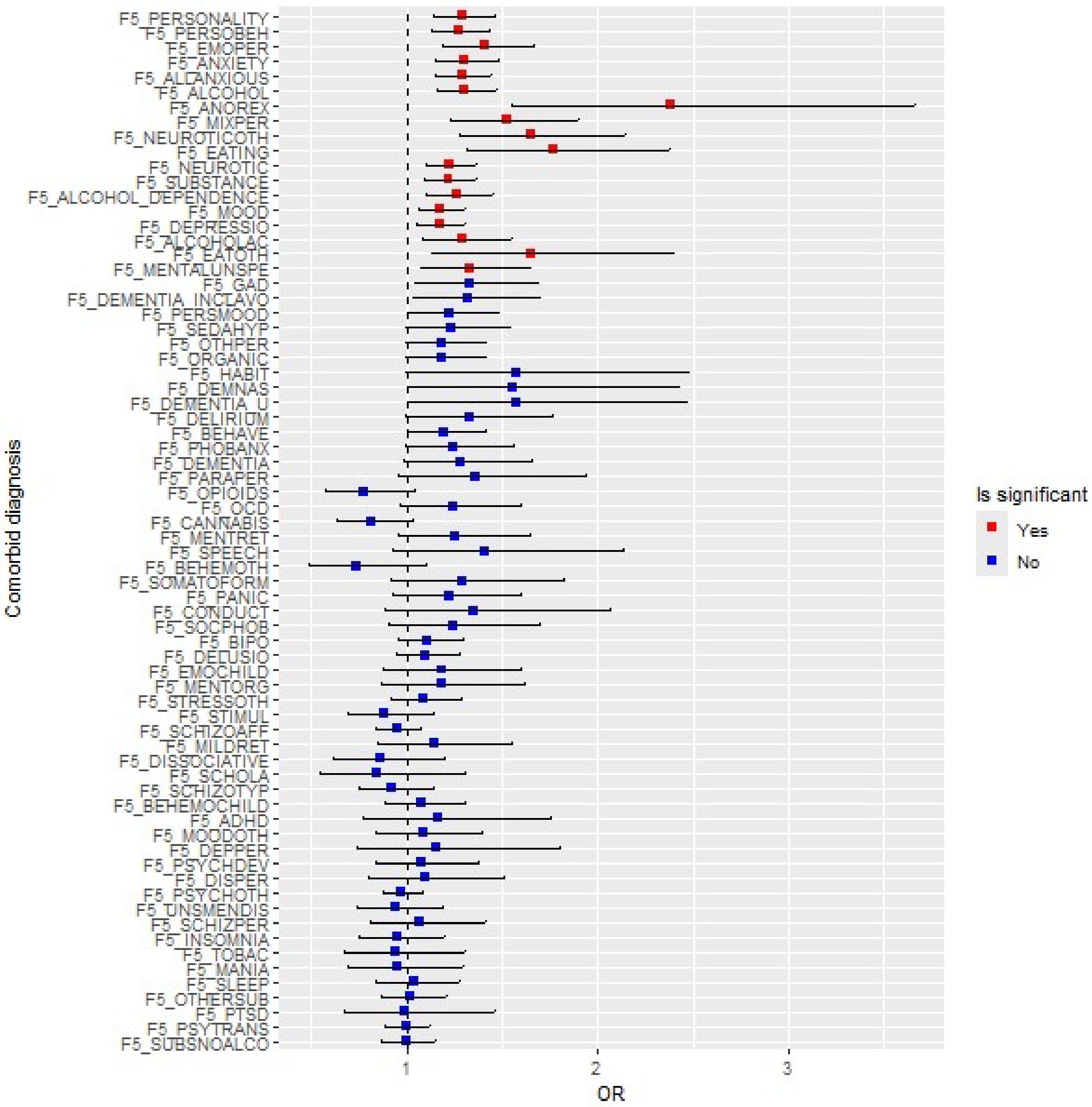


Red color indicates statistical significance at FDR of 0.05. The error bars show 95 % confidence intervals. GRP 2 was used as the reference group; therefore, ORs < 1 indicate higher rates in GRP 2, whereas ORs > 1 indicate higher rates in GRP 1. The definitions of the phenotype codes for comorbid diagnosis can be found in Supplementary Table 32.

**Supplementary Figure 24. Feature size 33: Differences in comorbidity rates of mental disorders and disorder categories between the GRPs in Train$_{FinnGen}$.**

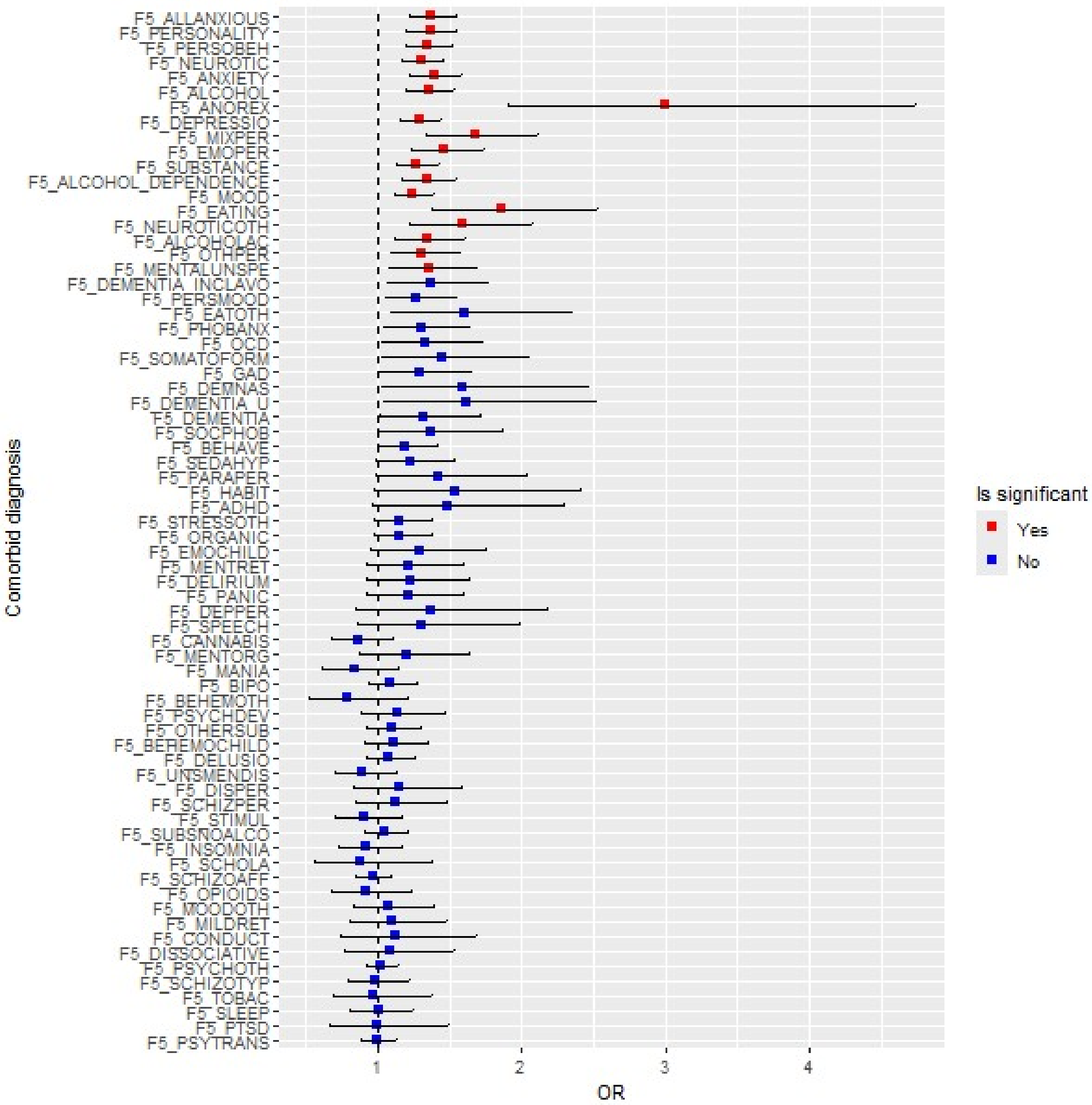


Red color indicates statistical significance at FDR of 0.05. The error bars show 95 % confidence intervals. GRP 2 was used as the reference group; therefore, ORs < 1 indicate higher rates in GRP 2, whereas ORs > 1 indicate higher rates in GRP 1. The definitions of the phenotype codes for comorbid diagnosis can be found in Supplementary Table 32.

**Supplementary Figure 25. Feature size 45: Differences in comorbidity rates of mental disorders and disorder categories between the GRPs in Train$_{FinnGen}$.**

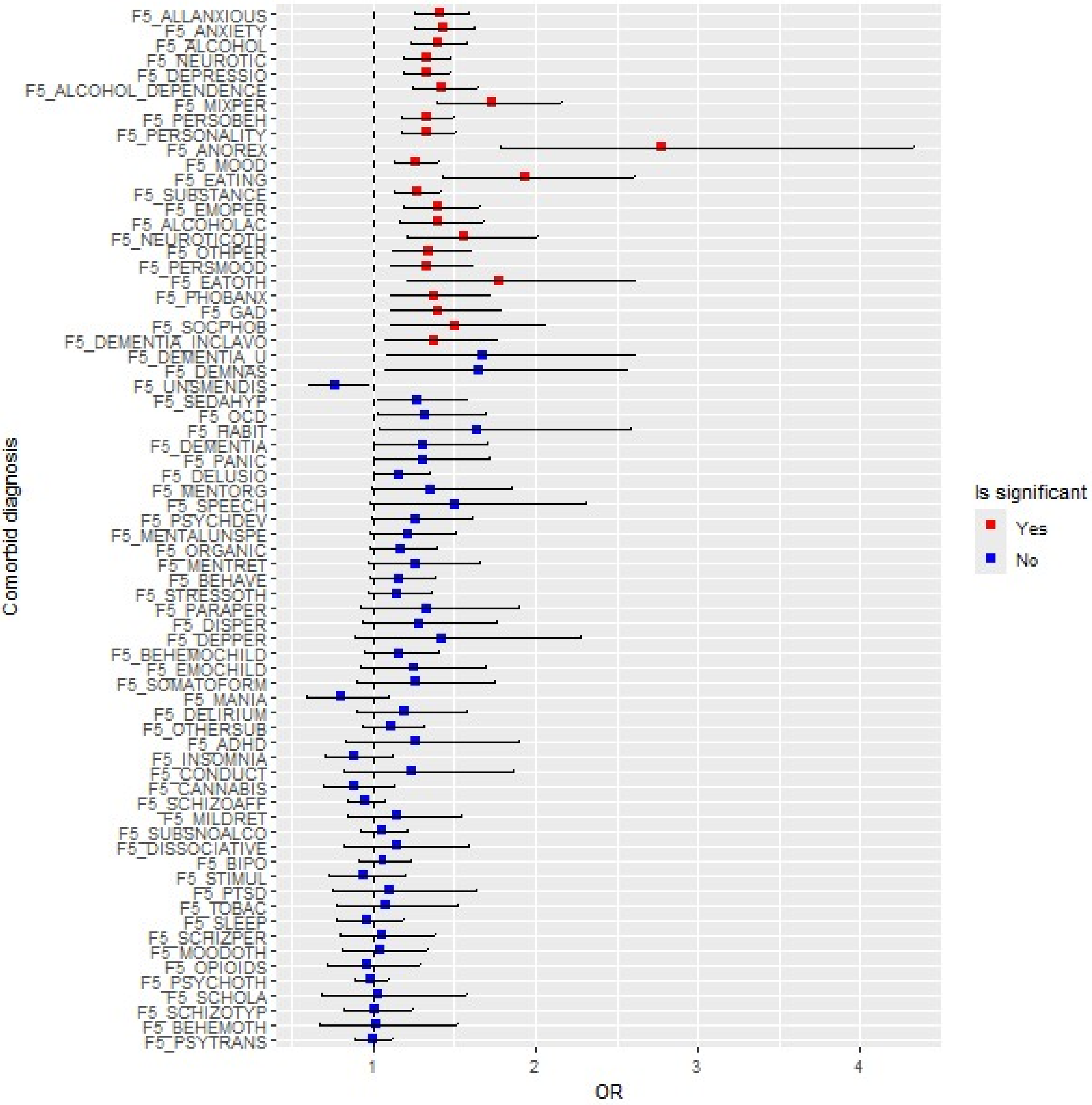


Red color indicates statistical significance at FDR of 0.05. The error bars show 95 % confidence intervals. GRP 2 was used as the reference group; therefore, ORs < 1 indicate higher rates in GRP 2, whereas ORs > 1 indicate higher rates in GRP 1. The definitions of the phenotype codes for comorbid diagnosis can be found in Supplementary Table 32.

**Supplementary Figure 26. Difference in Clozapine and antidepressants prescription rates between the GRPs across feature sizes in Train$_{FinnGen}$.**

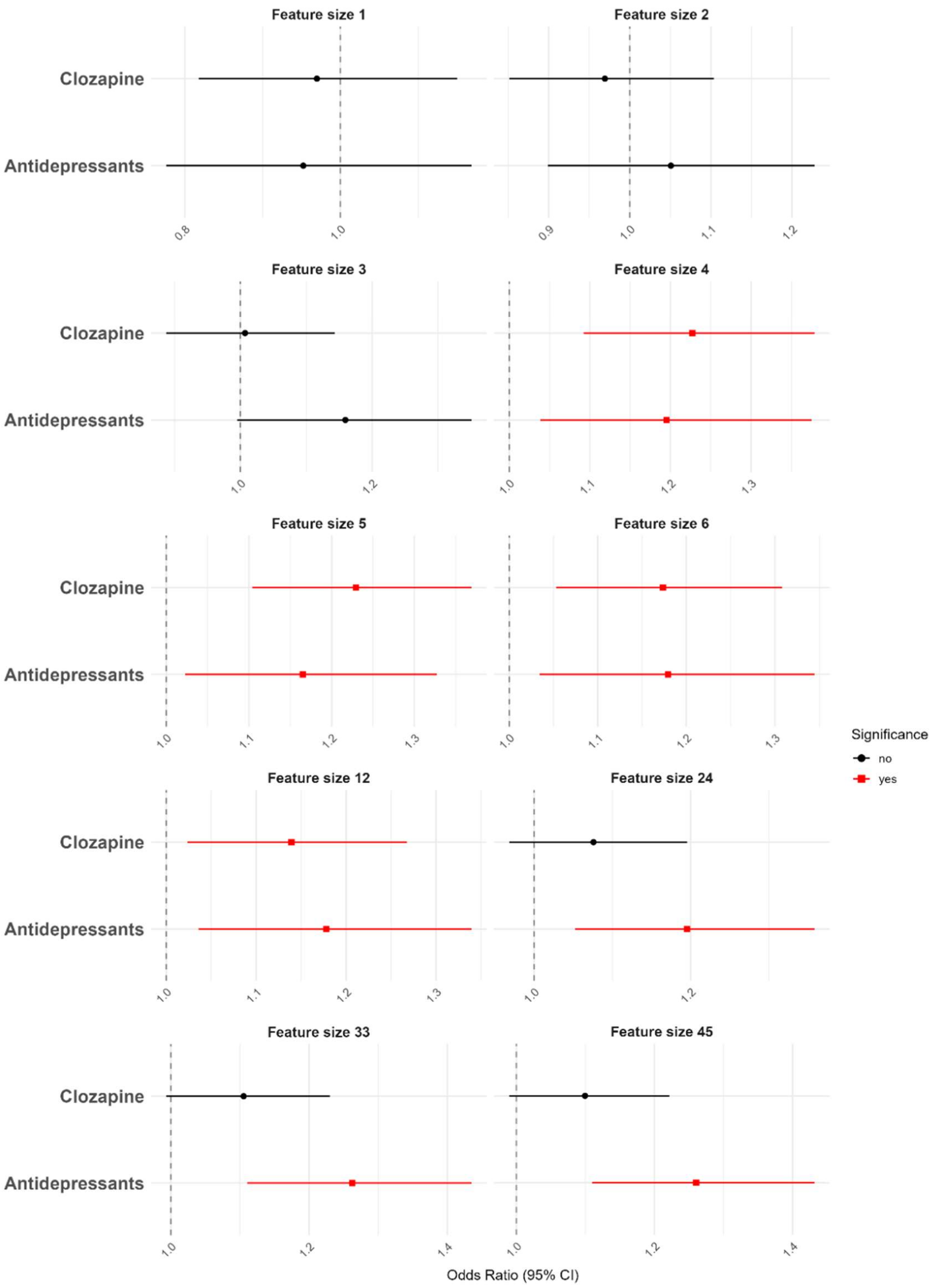


Red color indicates statistical significance at FDR of 0.05. The error bars show 95 % confidence intervals. GRP 2 was used as the reference group; therefore, ORs < 1 indicate higher prescription rates in GRP 2, whereas ORs > 1 indicate higher prescription rates in GRP 1.

**Supplementary Figure 27. Associations of clozapine and antidepressant prescriptions with GRPs across all feature sizes in $\text{Val}_{\text{PsyCourse}}$ using the primary assignment method and the sensitivity-analysis method.**

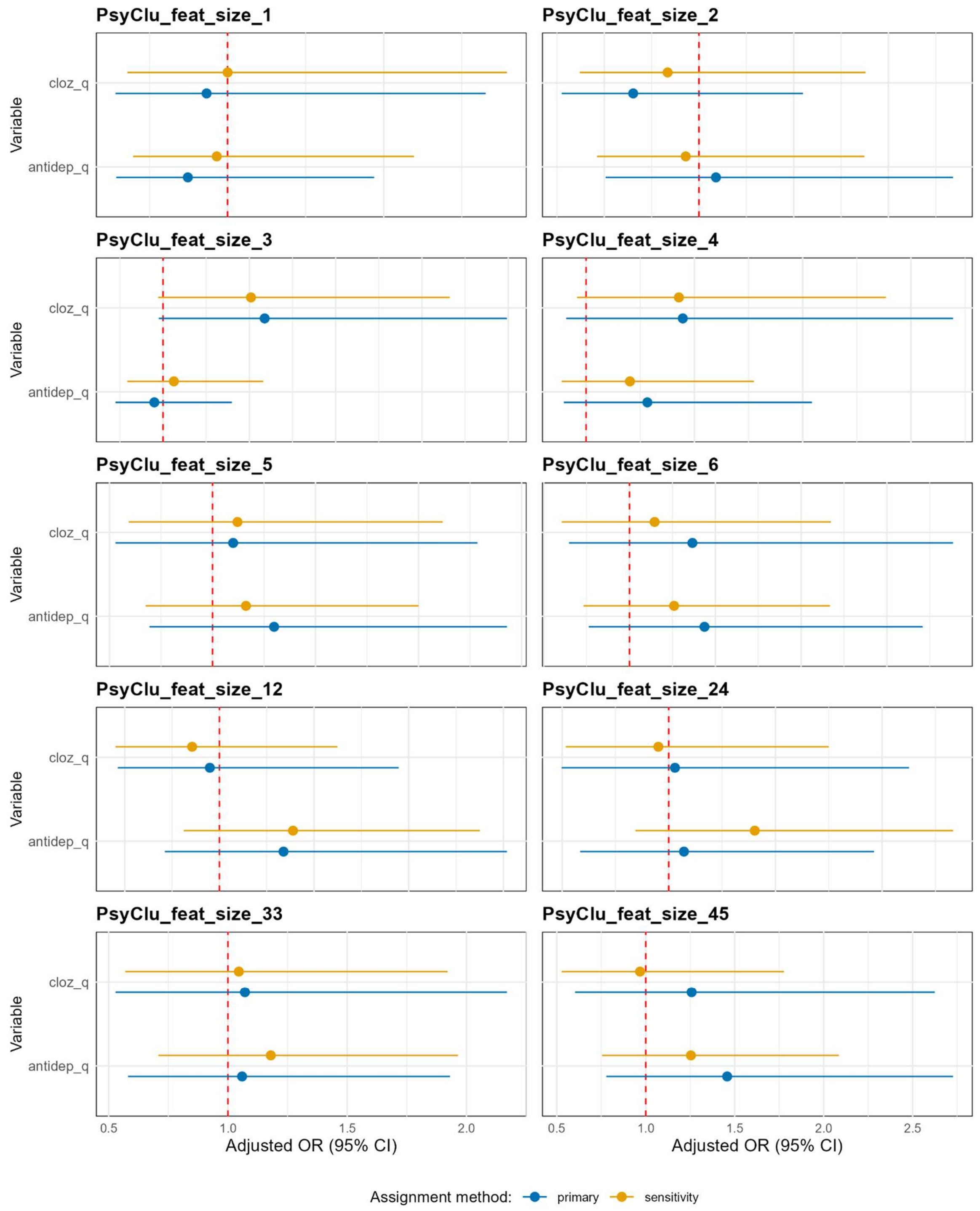


The error bars show 95 % confidence intervals. Orange indicates the method in which each SCZ patient was assigned to the GRP corresponding to the model that predicted case status with the higher probability. Blue indicates the method in which SCZ patients were assigned to the corresponding GRP only if they were correctly identified as cases by one of the two prediction models. GRP 2 was used as the reference group; therefore, ORs < 1 indicate higher prescription rates in GRP 2, whereas ORs > 1 indicate higher prescription rates in GRP 1. cloz_q : Clozapine prescription, antidep_q : Antidepressant prescription

**Supplementary Figure 28. Associations of PANSS variables with GRPs across all feature sizes in $\text{Val}_{\text{PsyCourse}}$ using the primary assignment method and the sensitivity-analysis method.**

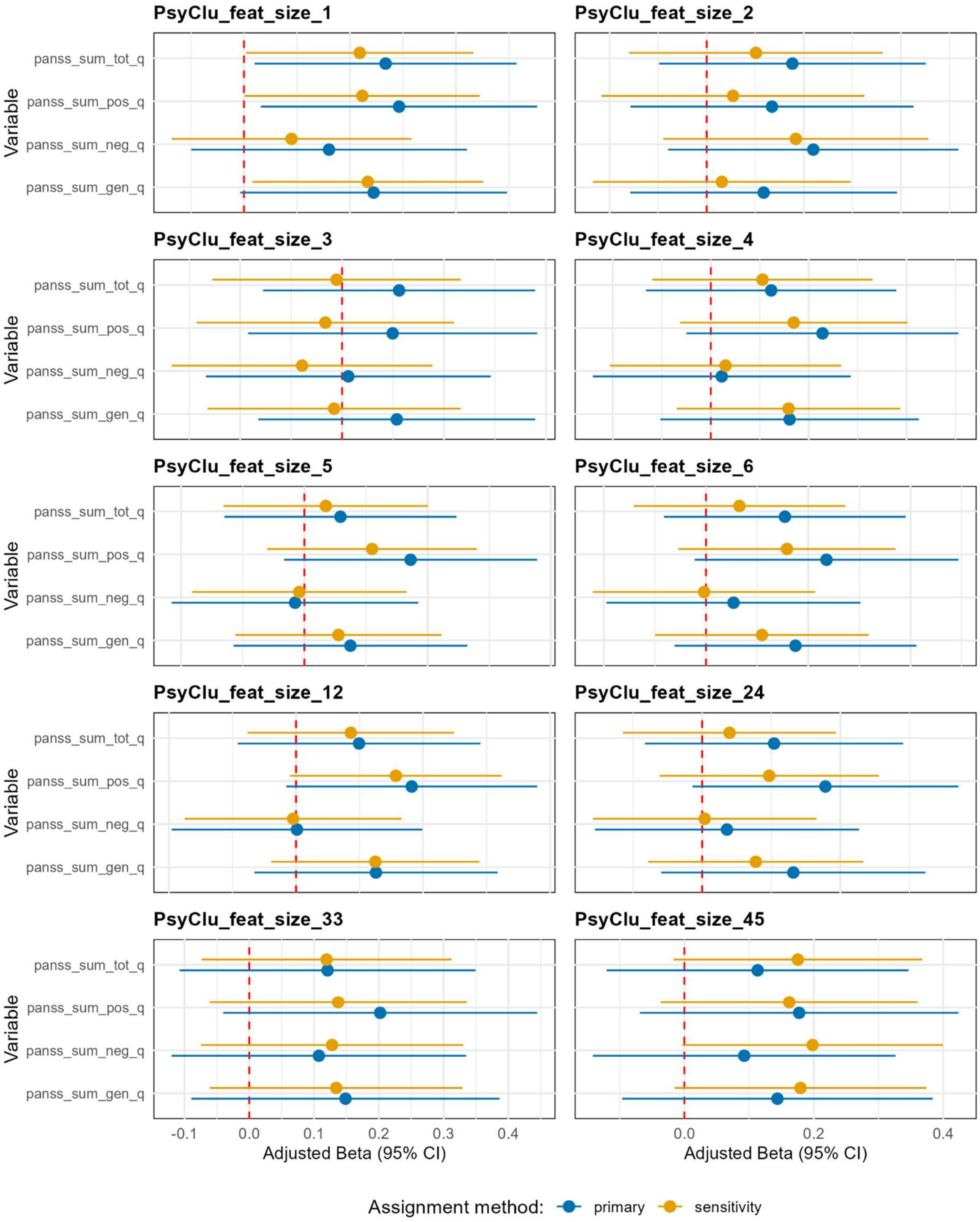

The error bars show 95 % confidence intervals. Orange indicates the method in which each SCZ patient was assigned to the GRP corresponding to the model that predicted case status with the higher probability. Blue indicates the method in which SCZ patients were assigned to the corresponding GRP only if they were correctly identified as cases by one of the two prediction models. GRP 2 was used as the reference group; therefore, Beta $< 0$ indicates higher scores in GRP 2, whereas Beta $> 0$ indicates higher higher scores in GRP 1. PsyCourse variable names can be found in Supplementary Table 30.

**Supplementary Figure 29. Associations of PANSS variables with GRPs across all feature sizes in $Val_{Bari}$ using the primary assignment method and the sensitivity-analysis method.**

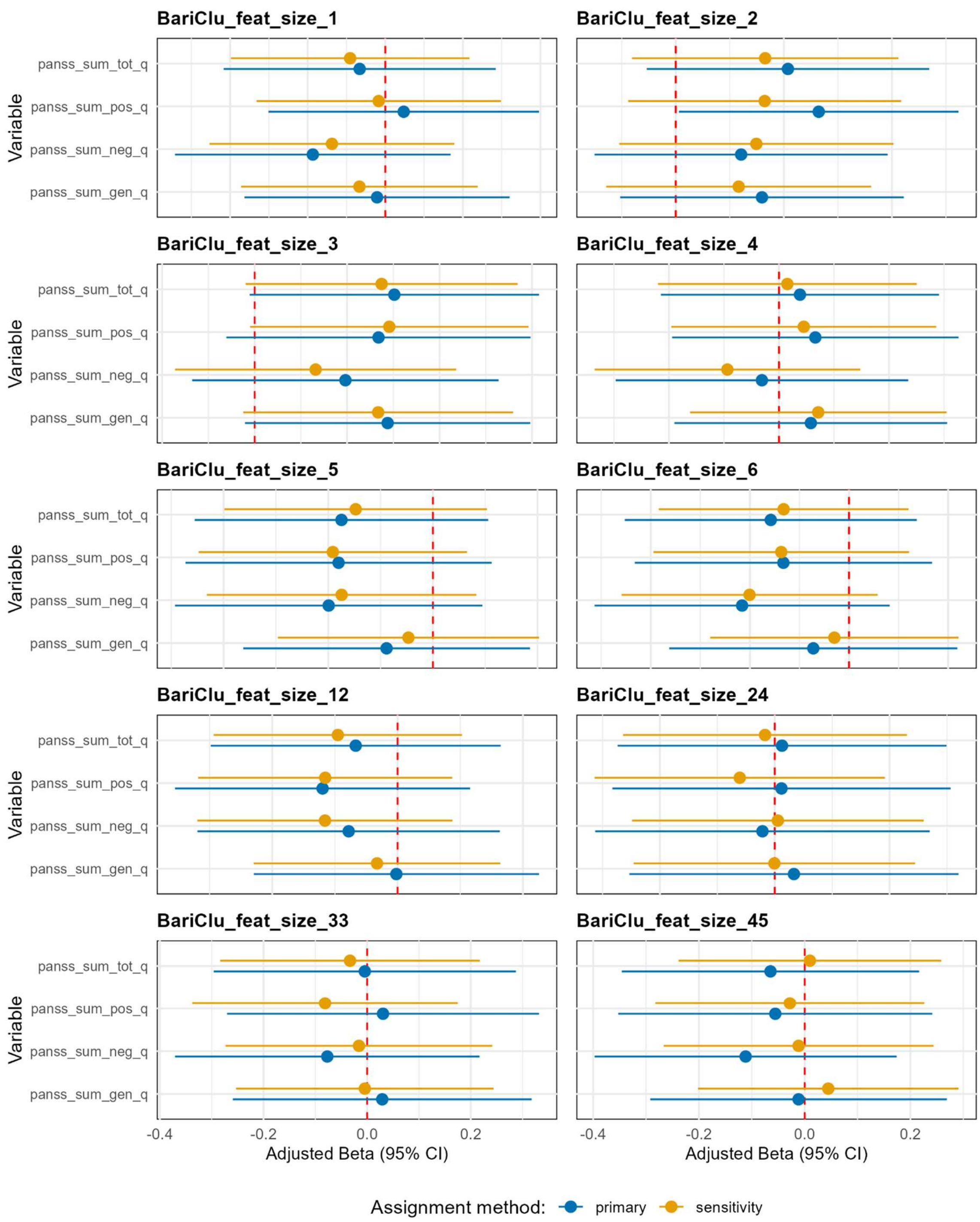


670

The error bars show 95 % confidence intervals. Orange indicates the method in which each SCZ patient was assigned to the GRP corresponding to the model that predicted case status with the higher probability. Blue indicates the method in which SCZ patients were assigned to the corresponding GRP only if they were correctly identified as cases by one of the two prediction models. GRP 2 was used as the reference group; therefore, Beta < 0 indicates higher scores in GRP 2, whereas Beta > 0 indicates higher higher scores in GRP 1. PsyCourse variable names can be found in Supplementary Table 30.

**Supplementary Figure 30. Cluster associations of selected phenotypes in Val$_{PsyCourse}$ across all feature sizes**

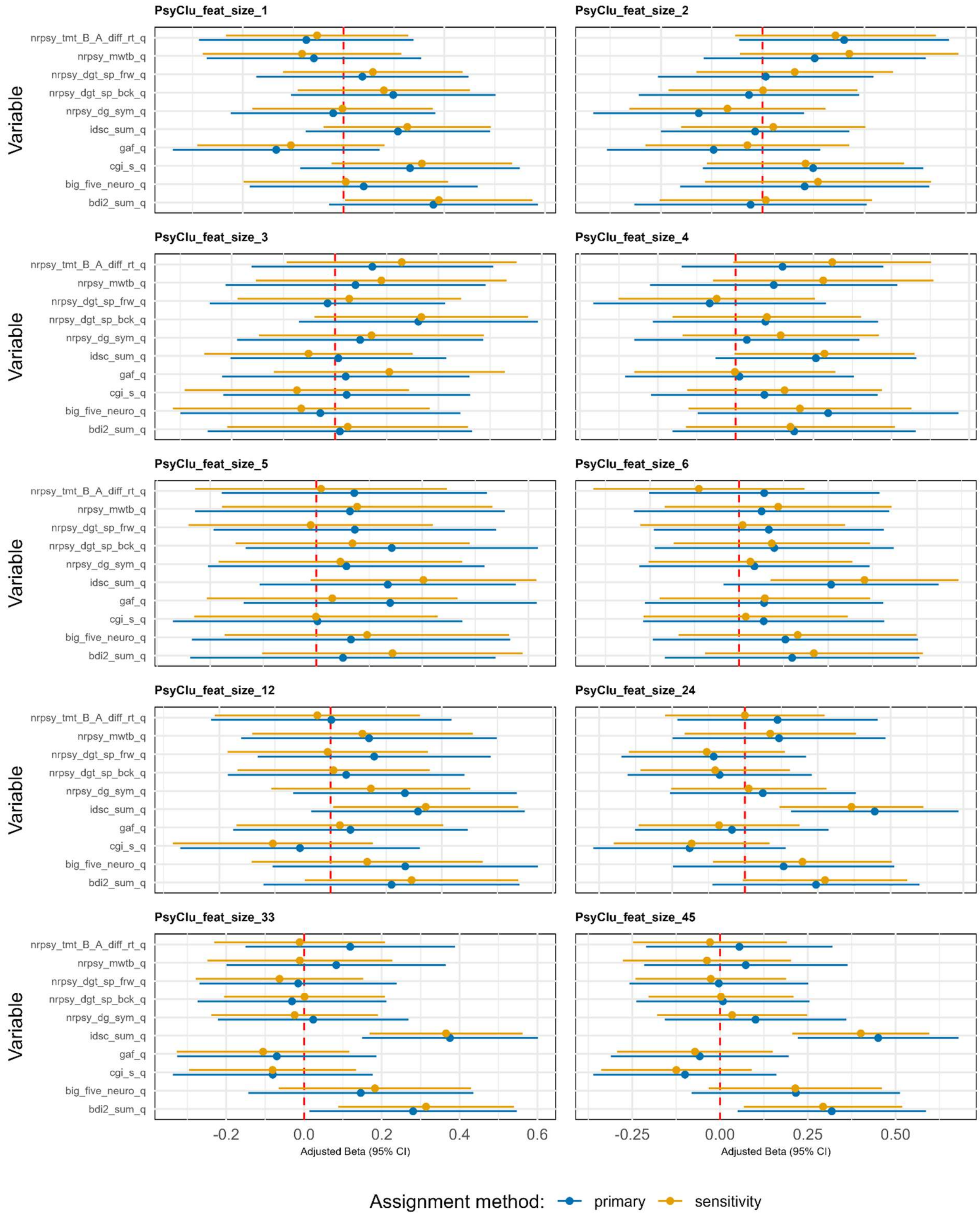


The error bars show 95 % confidence intervals. Orange indicates the method in which each SCZ patient was assigned to the GRP corresponding to the model that predicted case status with the higher probability. Blue indicates the method in which SCZ patients were assigned to the corresponding GRP only if they were correctly identified as cases by one of the two prediction models. GRP 2 was used as the reference group; therefore, Beta < 0 indicates higher scores in GRP 2, whereas Beta > 0 indicates higher higher scores in GRP 1. PsyCourse variable names can be found in Supplementary Table 30.

**Supplementary Figure 31. Feature size 1: Differences in comorbidity rates of mental disorders and disorder categories between the composite PGS groups of high vs low risk in Train$_{FinnGen}$.**

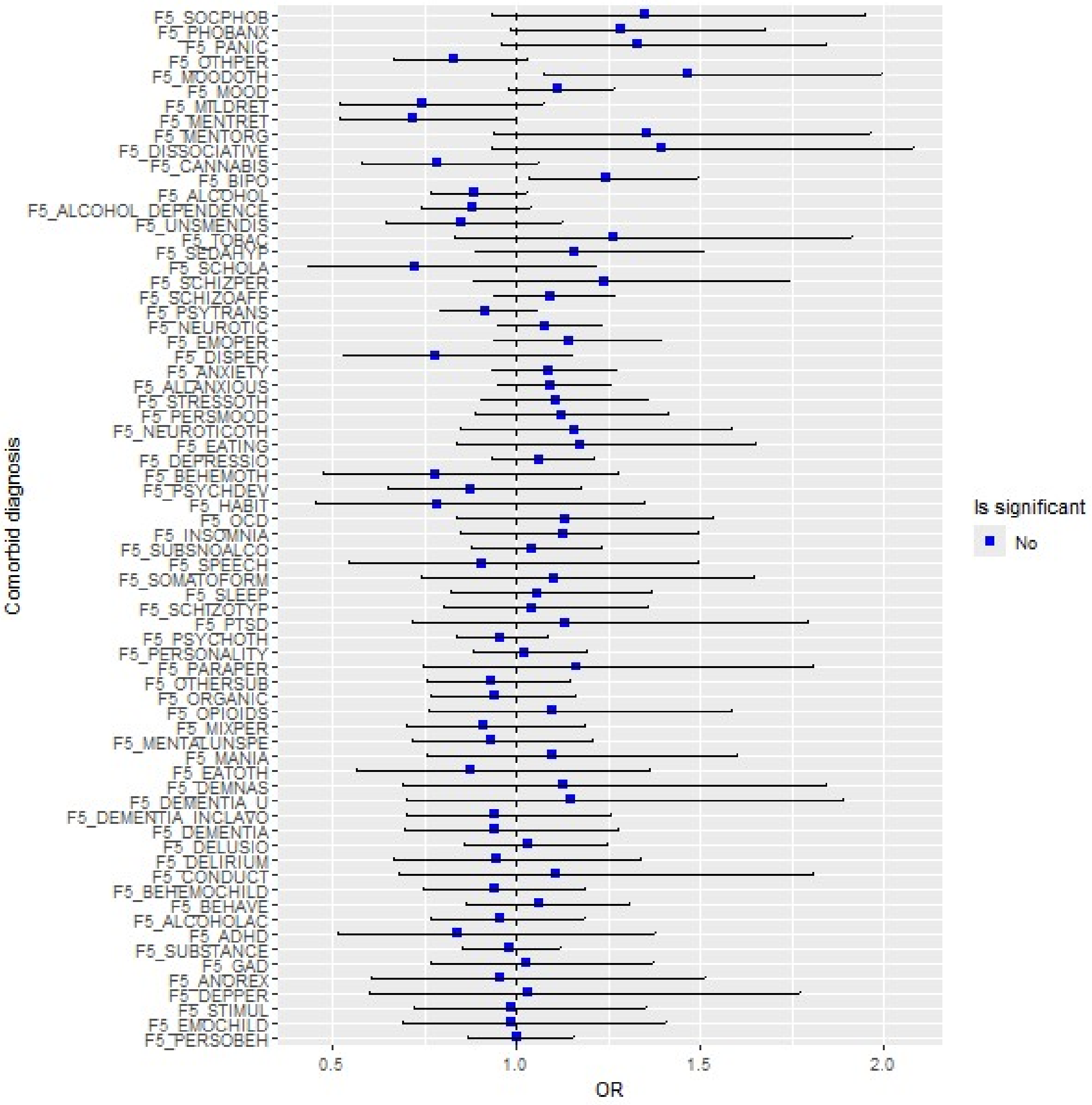


Red color indicates statistical significance at FDR of 0.05. The error bars show 95 % confidence intervals. Low-risk group was used as the reference group; therefore, ORs < 1 indicate higher rates in low-risk group, whereas ORs > 1 indicate higher rates in high-risk group. The definitions of the phenotype codes for comorbid diagnosis can be found in Supplementary Table 32.

**Supplementary Figure 32. Feature size 2: Differences in comorbidity rates of mental disorders and disorder categories between the composite PGS groups of high vs low risk in Train$_{FinnGen}$.**

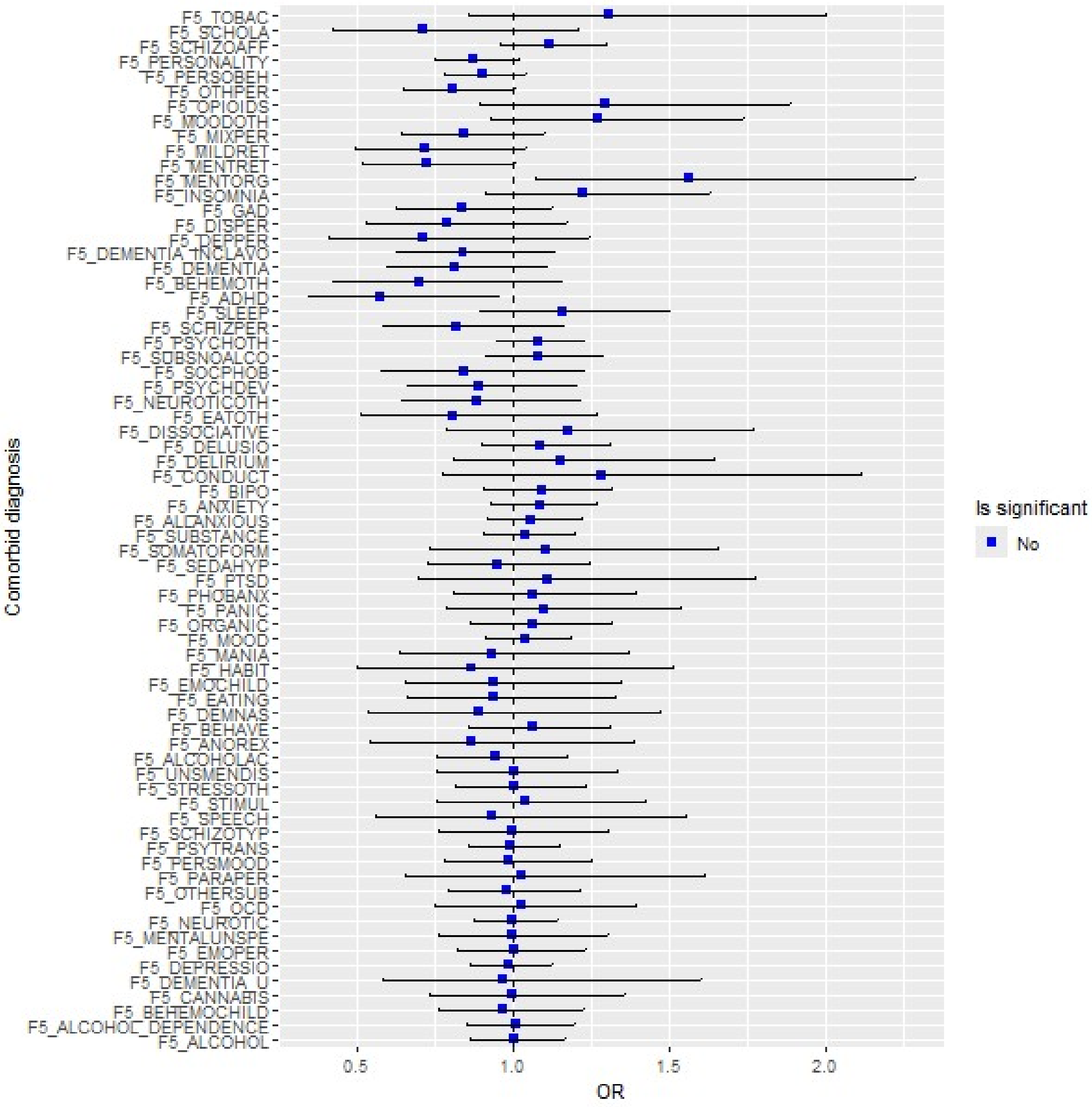


Red color indicates statistical significance at FDR of 0.05. The error bars show 95 % confidence intervals. Low-risk group was used as the reference group; therefore, ORs < 1 indicate higher rates in low-risk group, whereas ORs > 1 indicate higher rates in high-risk group. The definitions of the phenotype codes for comorbid diagnosis can be found in Supplementary Table 32.

**Supplementary Figure 33. Feature size 3: Differences in comorbidity rates of mental disorders and disorder categories between the composite PGS groups of high vs low risk in Train$_{FinnGen}$.**

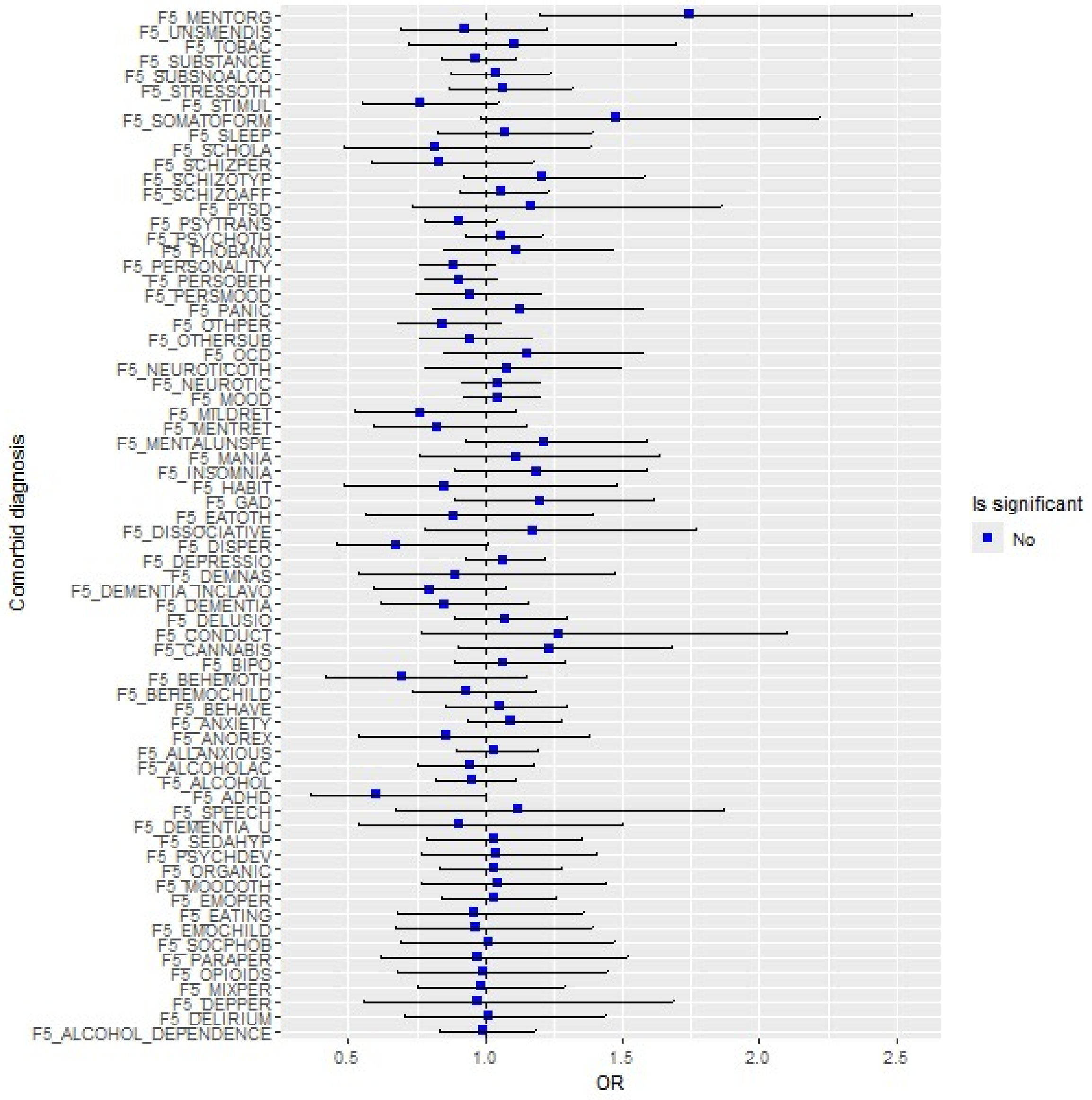


Red color indicates statistical significance at FDR of 0.05. The error bars show 95 % confidence intervals. Low-risk group was used as the reference group; therefore, ORs < 1 indicate higher rates in low-risk group, whereas ORs > 1 indicate higher rates in high-risk group. The definitions of the phenotype codes for comorbid diagnosis can be found in Supplementary Table 32.

**Supplementary Figure 34. Feature size 4: Differences in comorbidity rates of mental disorders and disorder categories between the composite PGS groups of high vs low risk in Train$_{FinnGen}$.**

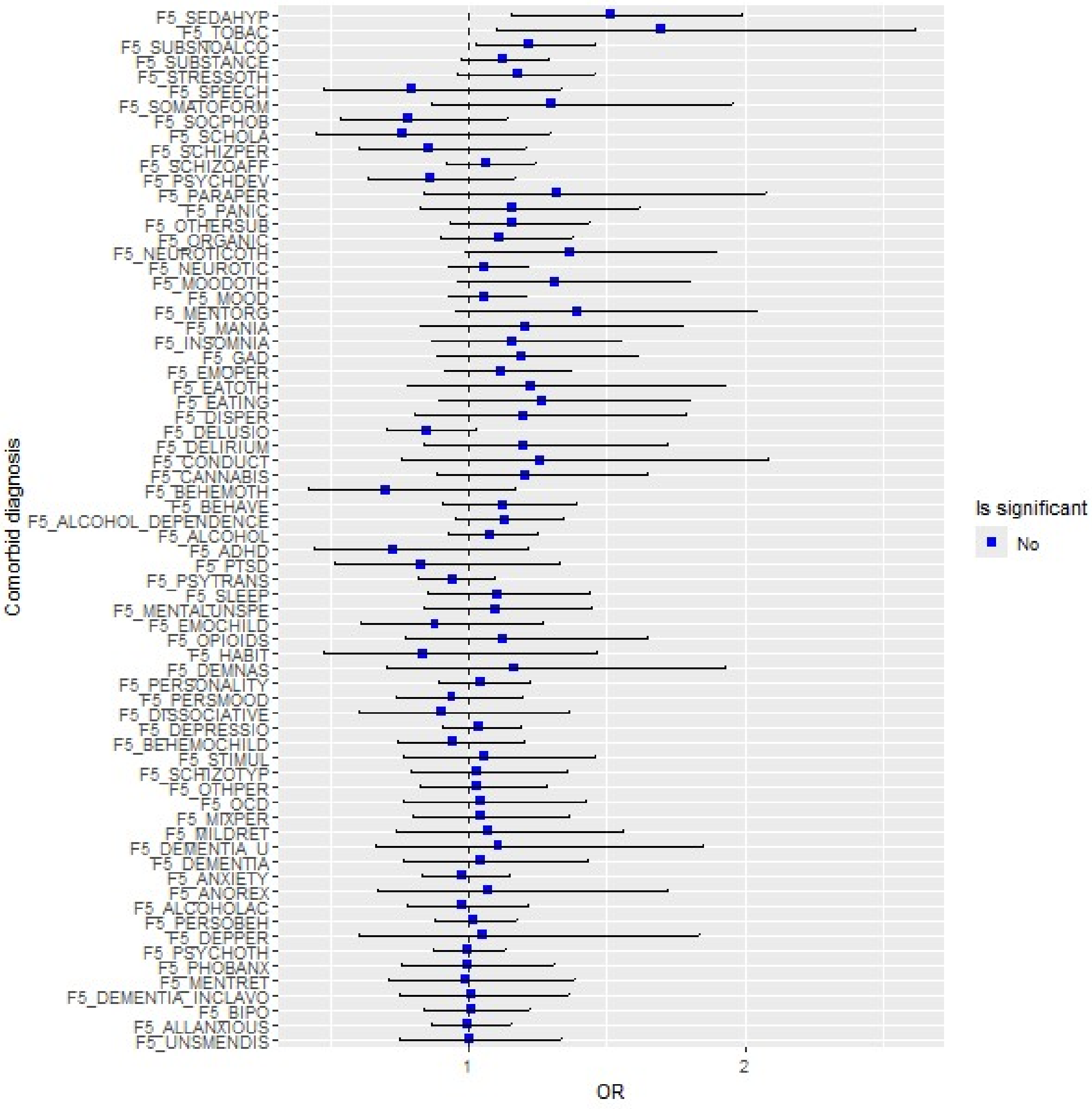


Red color indicates statistical significance at FDR of 0.05. The error bars show 95 % confidence intervals. Low-risk group was used as the reference group; therefore, ORs < 1 indicate higher rates in low-risk group, whereas ORs > 1 indicate higher rates in high-risk group. The definitions of the phenotype codes for comorbid diagnosis can be found in Supplementary Table 32.

**Supplementary Figure 35. Feature size 5: Differences in comorbidity rates of mental disorders and disorder categories between the composite PGS groups of high vs low risk in Train$_{FinnGen}$.**

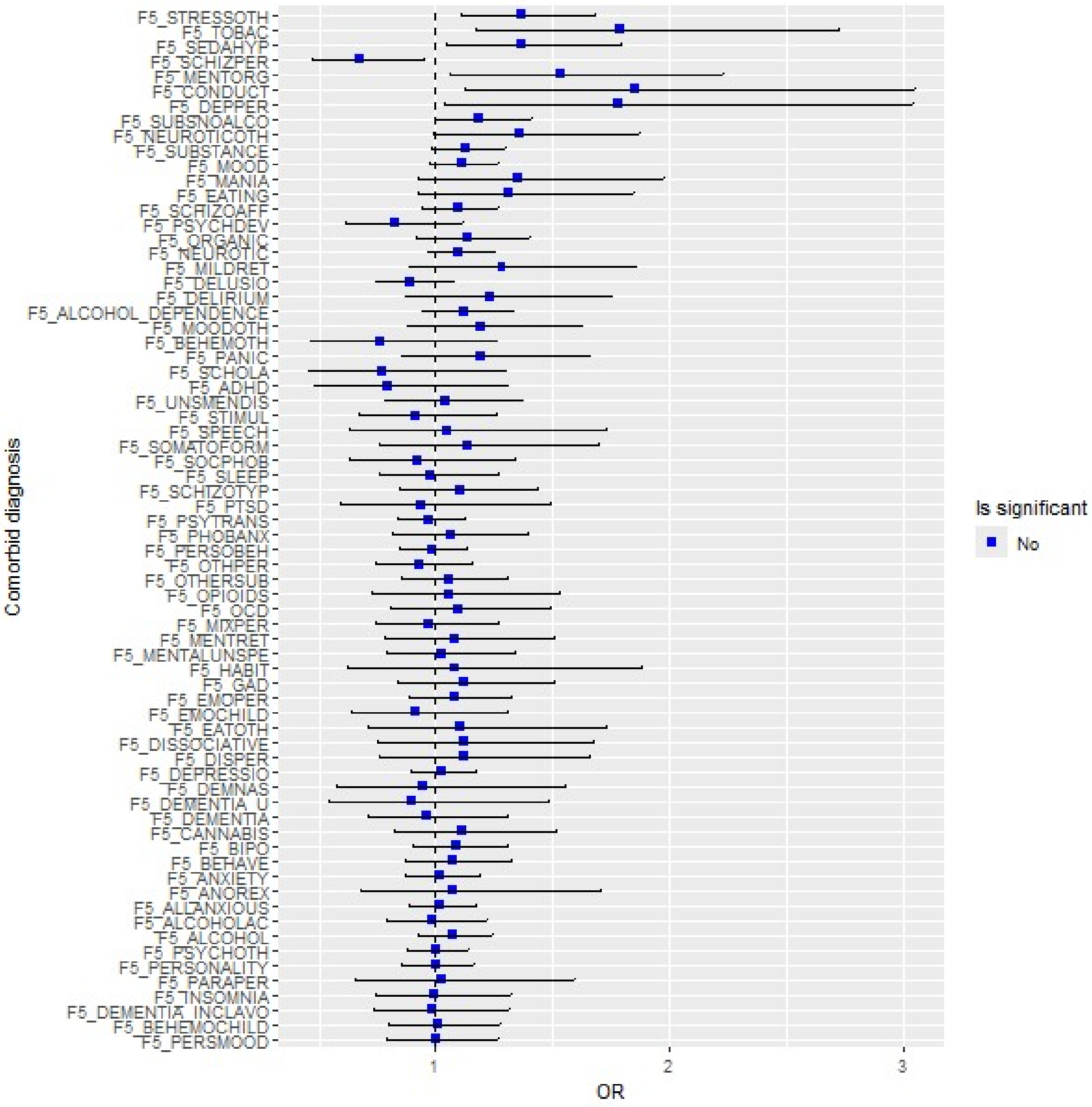


Red color indicates statistical significance at FDR of 0.05. The error bars show 95 % confidence intervals. Low-risk group was used as the reference group; therefore, ORs < 1 indicate higher rates in low-risk group, whereas ORs > 1 indicate higher rates in high-risk group. The definitions of the phenotype codes for comorbid diagnosis can be found in Supplementary Table 32.

**Supplementary Figure 36. Feature size 6: Differences in comorbidity rates of mental disorders and disorder categories between the composite PGS groups of high vs low risk in Train$_{FinnGen}$.**

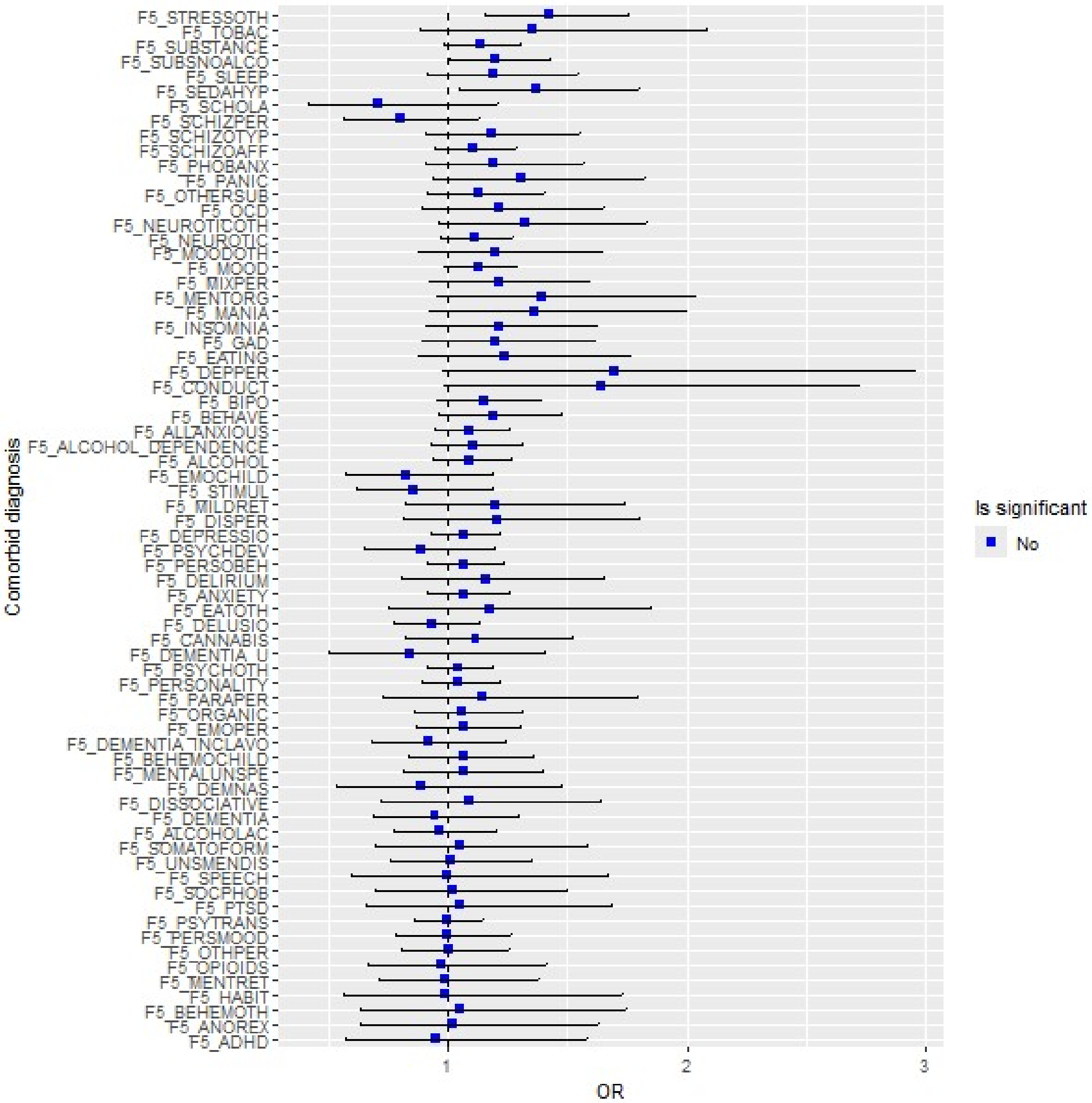


Red color indicates statistical significance at FDR of 0.05. The error bars show 95 % confidence intervals. Low-risk group was used as the reference group; therefore, ORs < 1 indicate higher rates in low-risk group, whereas ORs > 1 indicate higher rates in high-risk group. The definitions of the phenotype codes for comorbid diagnosis can be found in Supplementary Table 32.

**Supplementary Figure 37. Feature size 12: Differences in comorbidity rates of mental disorders and disorder categories between the composite PGS groups of high vs low risk in Train$_{FinnGen}$.**

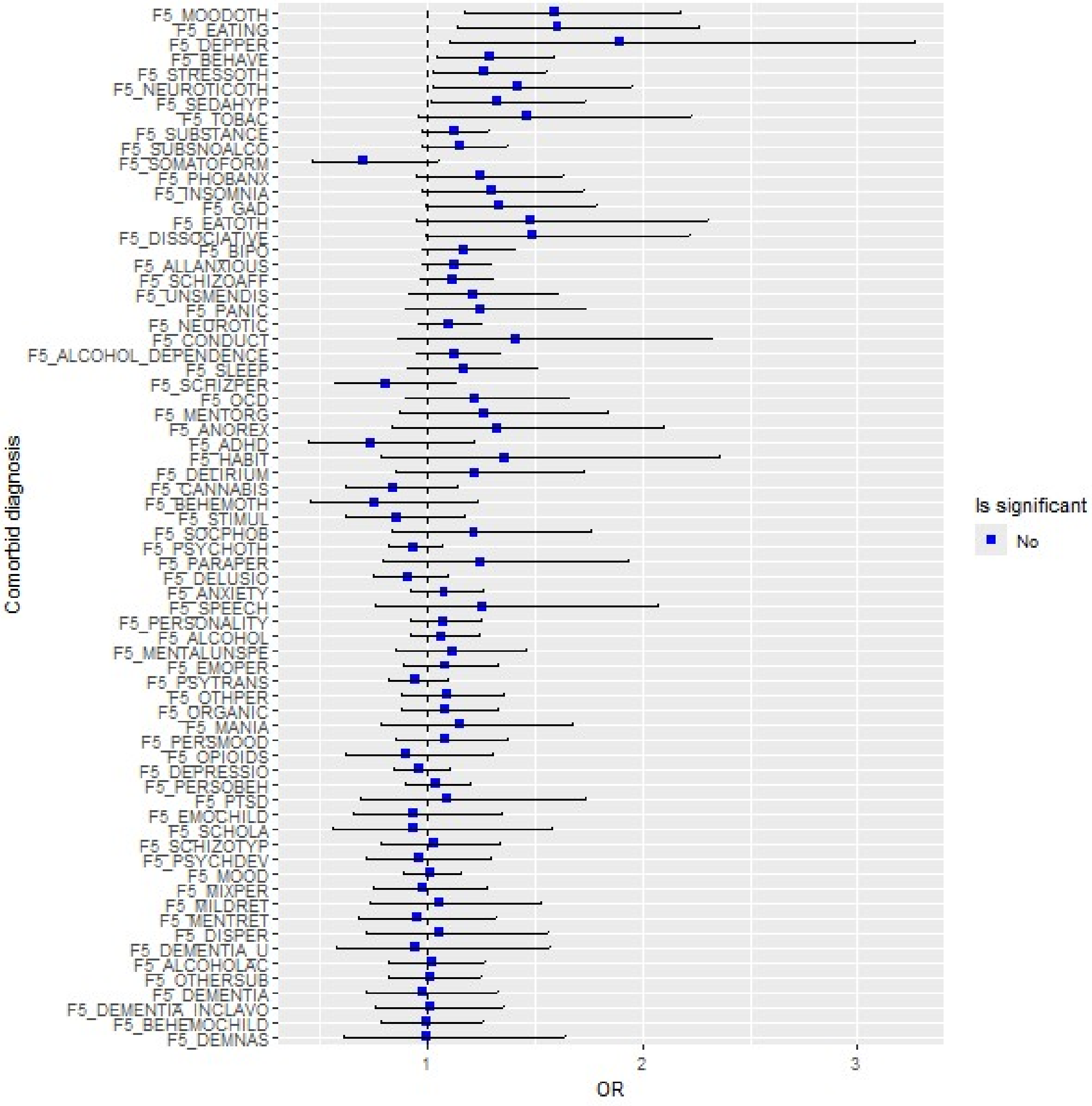


Red color indicates statistical significance at FDR of 0.05. The error bars show 95 % confidence intervals. Low-risk group was used as the reference group; therefore, ORs < 1 indicate higher rates in low-risk group, whereas ORs > 1 indicate higher rates in high-risk group. The definitions of the phenotype codes for comorbid diagnosis can be found in Supplementary Table 32.

**Supplementary Figure 38. Feature size 24: Differences in comorbidity rates of mental disorders and disorder categories between the composite PGS groups of high vs low risk in Train$_{FinnGen}$.**

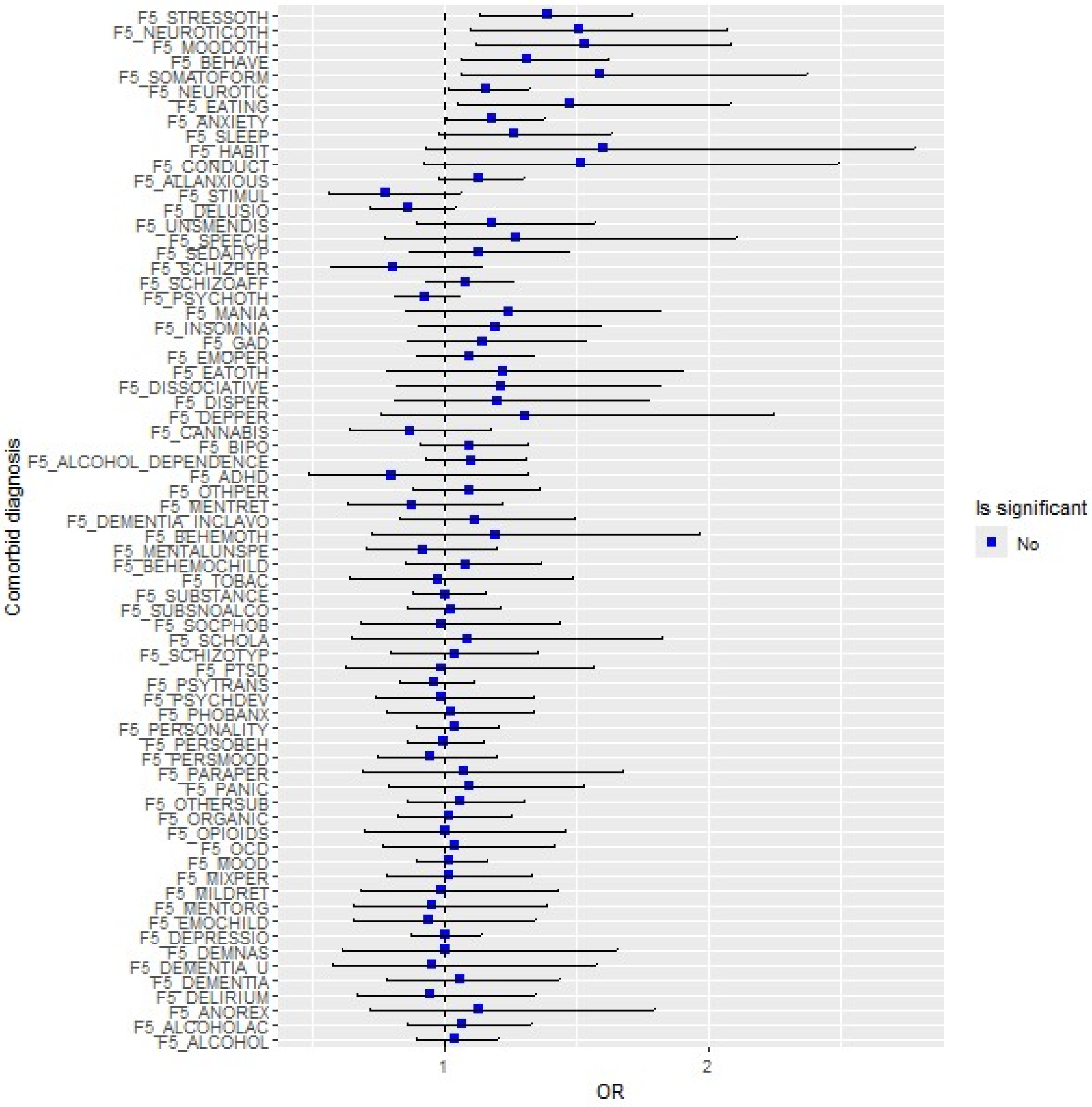


Red color indicates statistical significance at FDR of 0.05. The error bars show 95 % confidence intervals. Low-risk group was used as the reference group; therefore, ORs < 1 indicate higher rates in low-risk group, whereas ORs > 1 indicate higher rates in high-risk group. The definitions of the phenotype codes for comorbid diagnosis can be found in Supplementary Table 32.

**Supplementary Figure 39. Feature size 33: Differences in comorbidity rates of mental disorders and disorder categories between the composite PGS groups of high vs low risk in Train$_{FinnGen}$.**

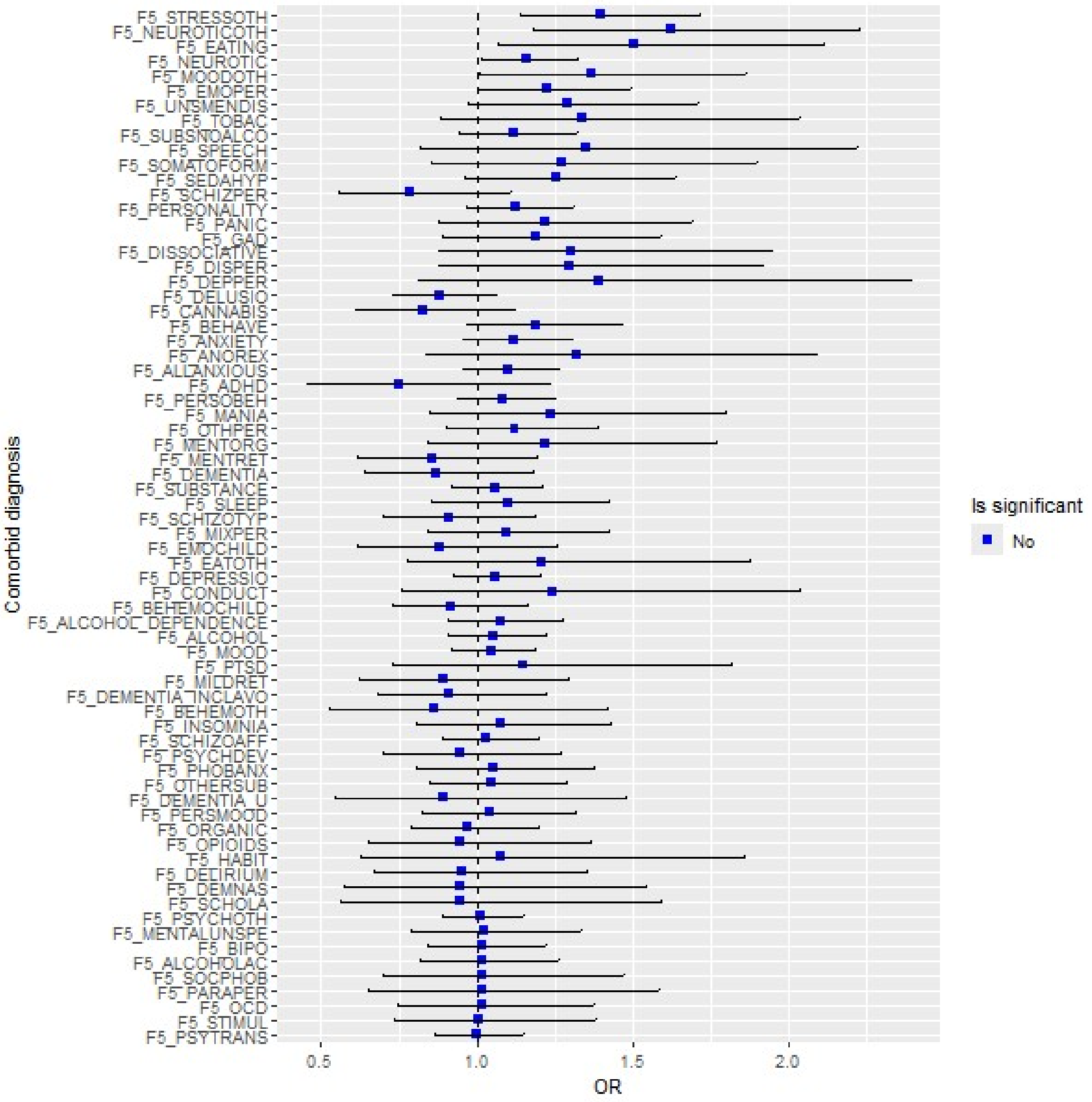


Red color indicates statistical significance at FDR of 0.05. The error bars show 95 % confidence intervals. Low-risk group was used as the reference group; therefore, ORs < 1 indicate higher rates in low-risk group, whereas ORs > 1 indicate higher rates in high-risk group. The definitions of the phenotype codes for comorbid diagnosis can be found in Supplementary Table 32.

**Supplementary Figure 40. Feature size 45: Differences in comorbidity rates of mental disorders and disorder categories between the composite PGS groups of high vs low risk in Train$_{FinnGen}$.**

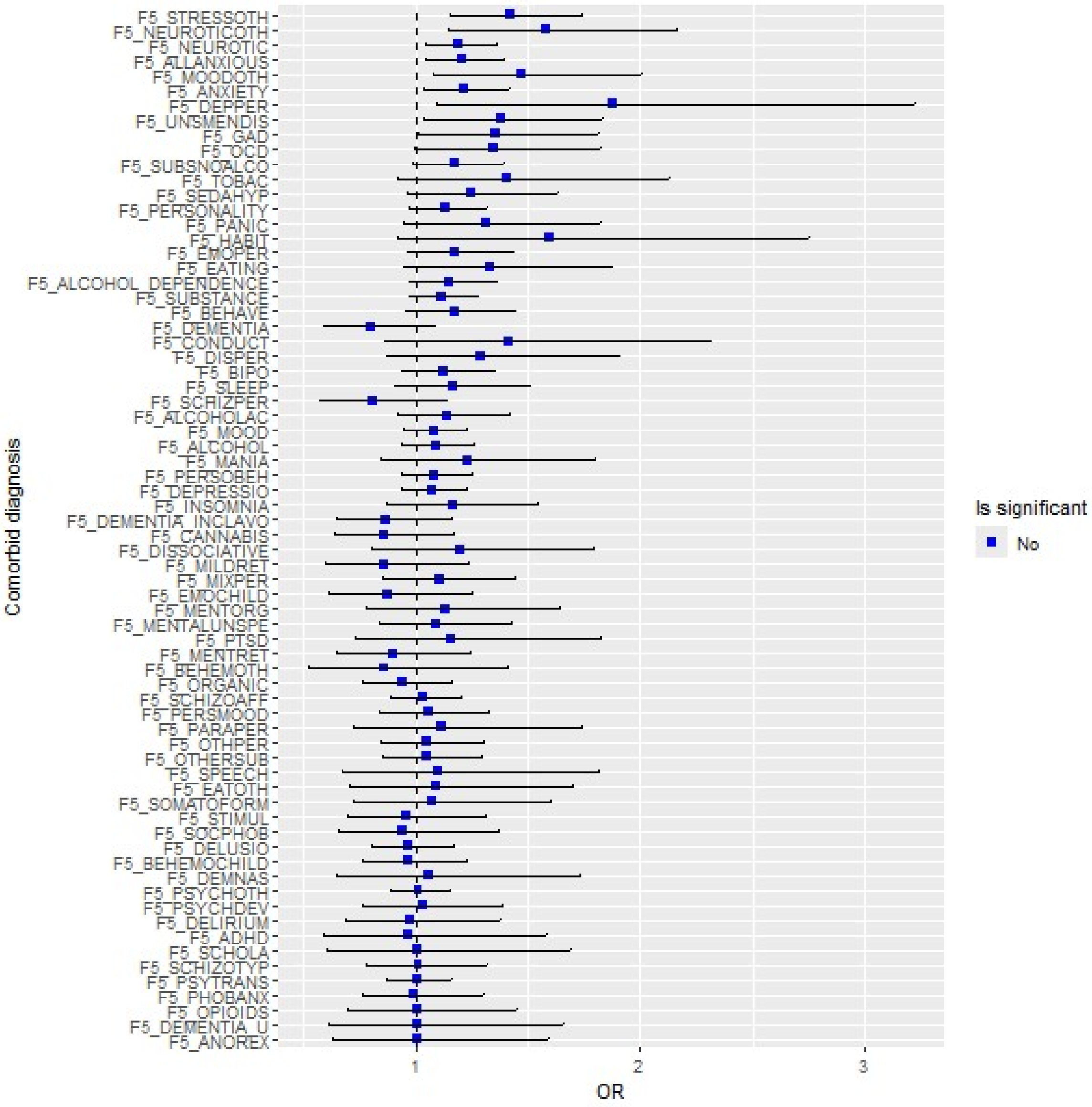


Red color indicates statistical significance at FDR of 0.05. The error bars show 95 % confidence intervals. Low-risk group was used as the reference group; therefore, ORs < 1 indicate higher rates in low-risk group, whereas ORs > 1 indicate higher rates in high-risk group. The definitions of the phenotype codes for comorbid diagnosis can be found in Supplementary Table 32.

**Supplementary Figure 41. Difference in Clozapine and antidepressants prescription rates between the composite PGS across feature sizes in Train$_{FinnGen}$.**

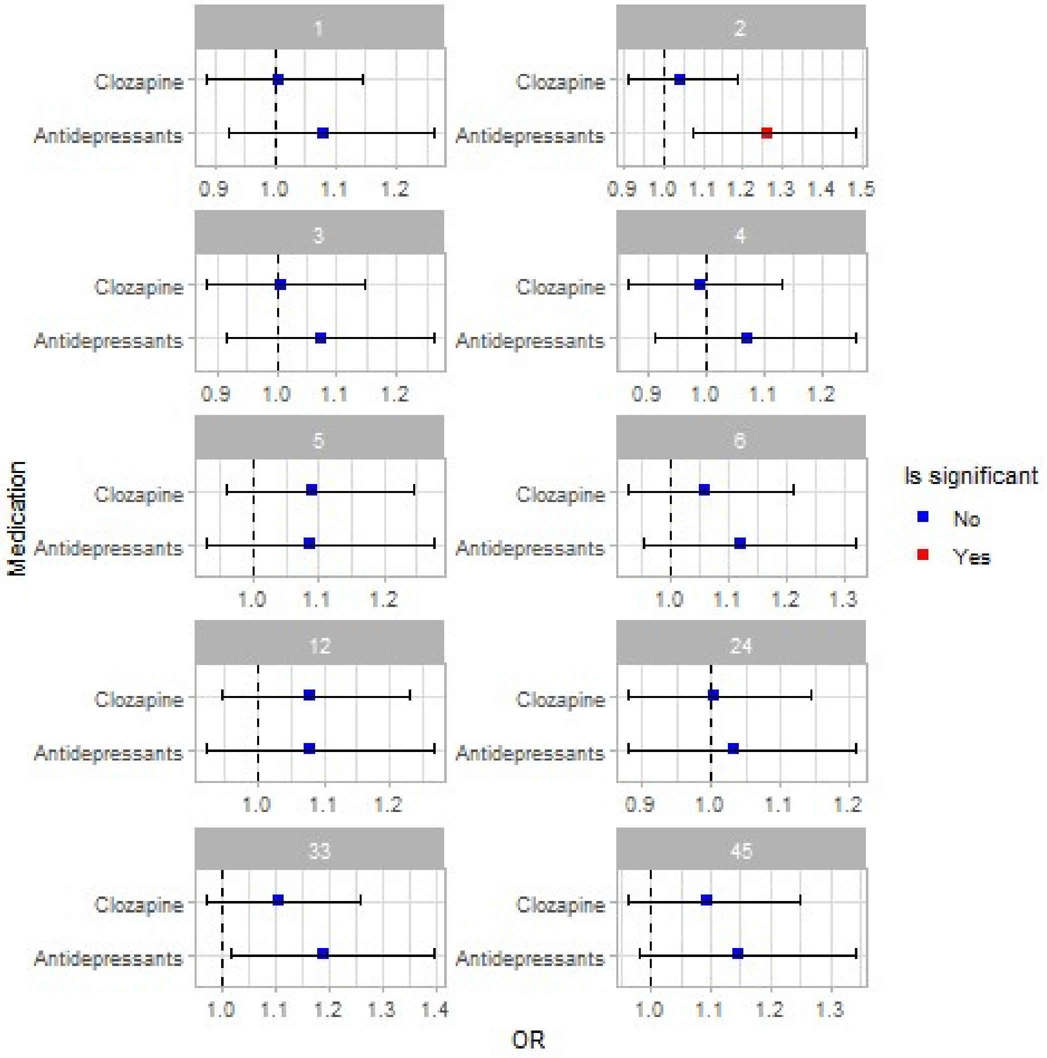


Red color indicates statistical significance at FDR of 0.05. The error bars show 95 % confidence intervals. Feature size is displayed above each panel. Low-risk groups were used as reference groups; therefore, ORs < 1 indicate higher prescription rates in low-risk groups, whereas ORs > 1 indicate higher prescription rates in high-risk groups.

**Supplementary Figure 42. Feature size 24: Differences in comorbidity rates of mental disorders and disorder categories between the GRPs in $\text{Train}_{\text{FinnGen}}$. Group assignment was based on the alternative GRP assignment method (assigning all SCZ cases, based on the highest probability) described in sensitivity analyses.**

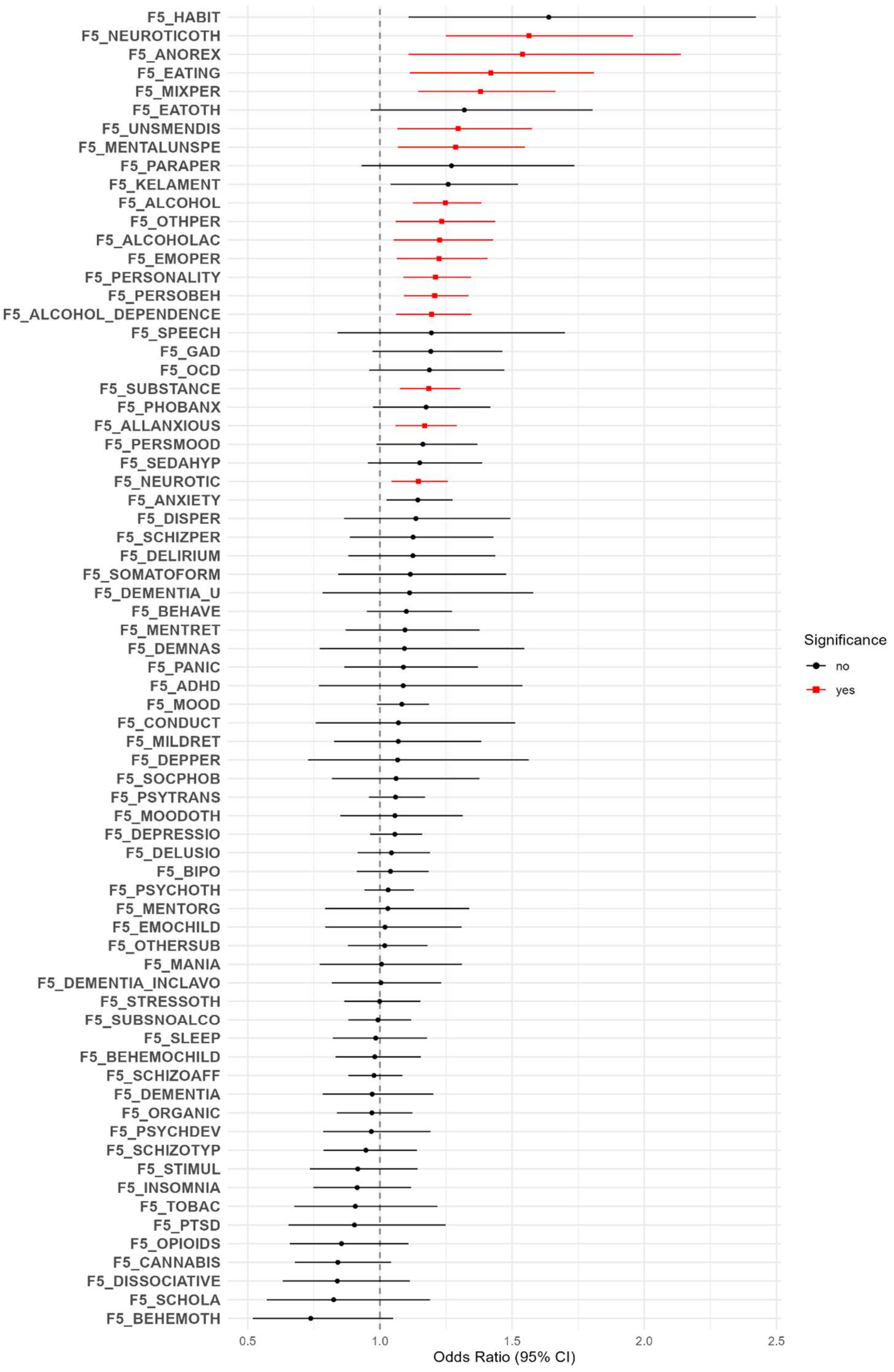

Red color indicates statistical significance at FDR of 0.05. The error bars show 95 % confidence intervals. GRP 2 was used as the reference group; therefore, ORs $< 1$ indicate higher rates in GRP 2, whereas ORs $> 1$ indicate higher rates in GRP 1. The definitions of the phenotype codes for comorbid diagnosis can be found in Supplementary Table 32.

**Supplementary Figure 43. Associations of clozapine and antidepressant prescriptions with GRPs at feature size 24 in $Train_{FinnGen}$ using the primary assignment method and the sensitivity-analysis method.**

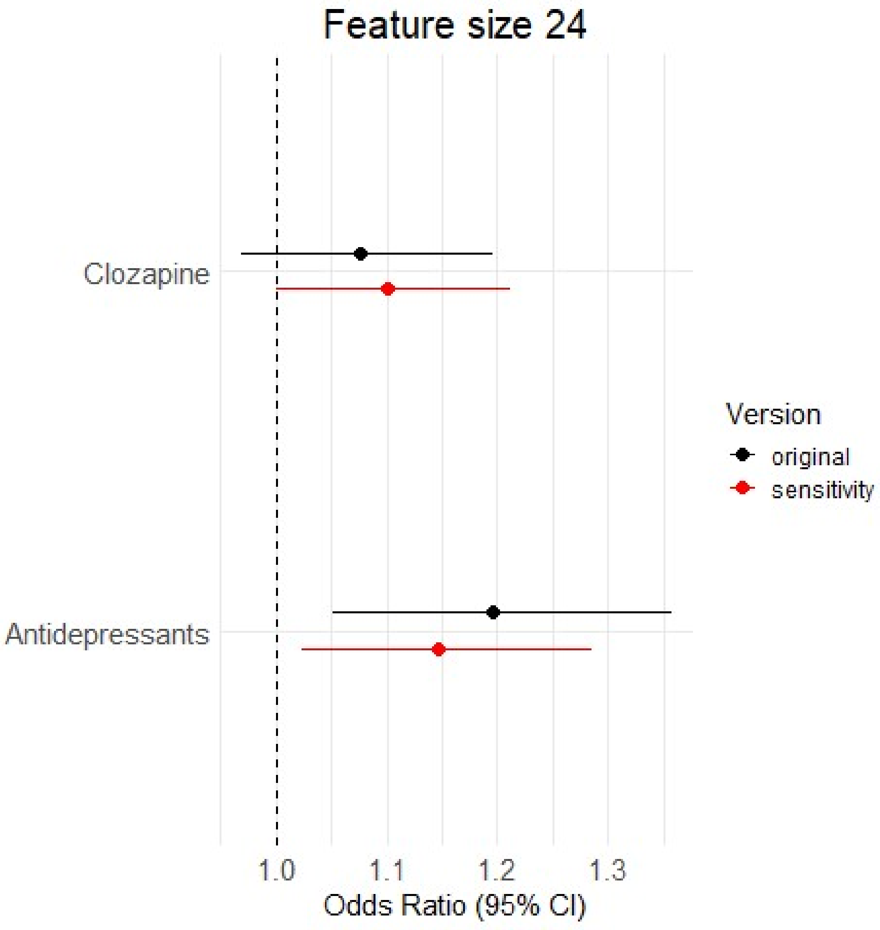


The error bars show 95 % confidence intervals. Red indicates the method in which each SCZ patient was assigned to the GRP corresponding to the model that predicted case status with the higher probability. Black indicates the method in which SCZ patients were assigned to the corresponding GRP only if they were correctly identified as cases by one of the two prediction models. GRP 2 was used as the reference group; therefore, ORs < 1 indicate higher prescription rates in GRP 2, whereas ORs > 1 indicate higher prescription rates in GRP 1.

**Supplementary Figure 44. Differential diagnostic associations of SCZ GRPs across feature sizes in $Train_{FinnGen}$**

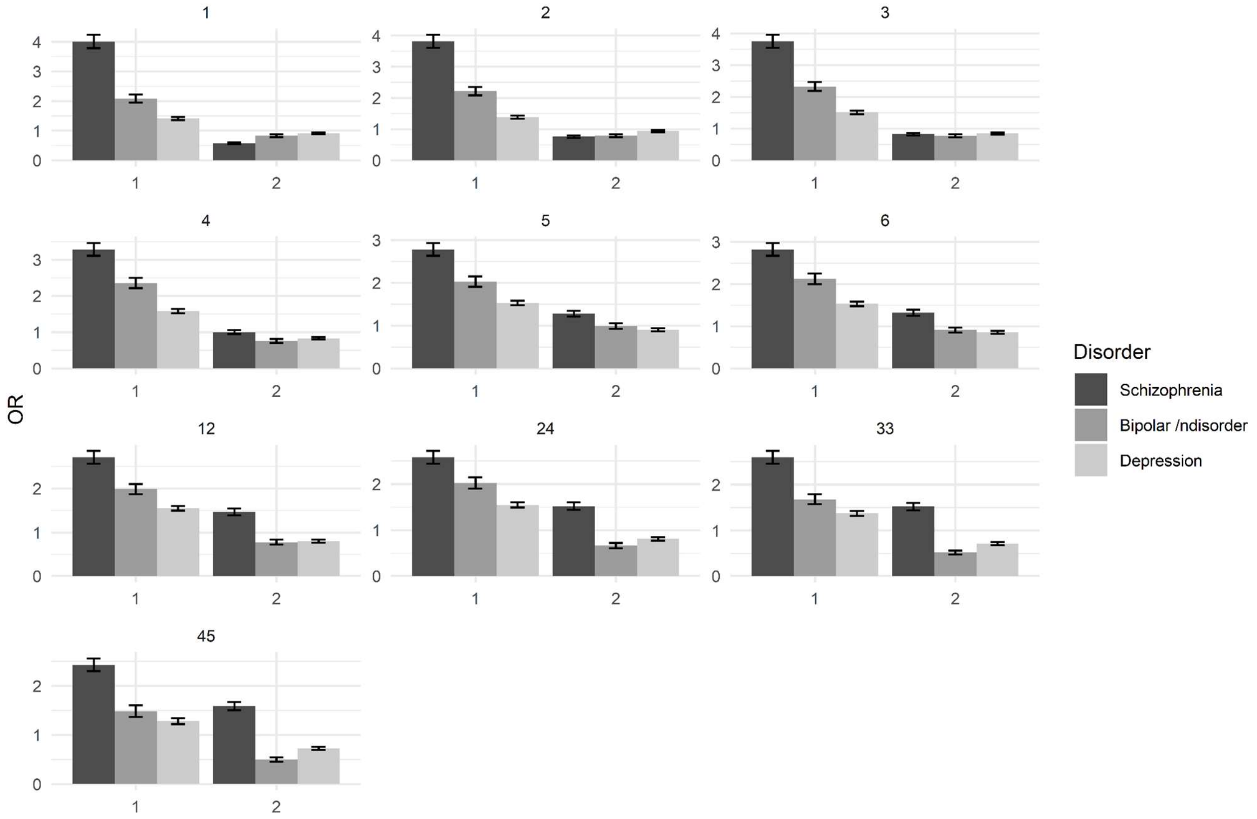


Feature sizes are displayed at the top of each panel, and the GRP on which a given model is based is presented on the x-axis.
